# Single-Cell Gene Expression Heritability Informed by Genotype × Cell-Context Interactions

**DOI:** 10.64898/2026.11.23.26368758

**Authors:** Lida Wang, Chaimaa Fadil, Lili Wang, Krisna Van Dyke, Alexander Gusev

## Abstract

Most signals from genome-wide association studies (GWAS) of complex traits implicate noncoding variants with putative gene regulatory effects. However, currently identified regulatory variants, particularly expression quantitative trait loci (eQTLs), explain only a small fraction of GWAS associations. Moreover, GWAS loci are enriched near evolutionarily constrained genes, whereas constrained genes are depleted of detectable eQTLs, suggesting that weak or context-specific regulatory mechanisms may contribute substantially to disease risk but remain poorly captured by existing approaches. Here, we introduce scHINT (single-cell gene expression Heritability estimation with genotype × context INTeraction), a computationally efficient framework for quantifying cell-level expression heritability and context-dependent genetic regulation from large-scale single-cell data. Applying scHINT to human brain single-nucleus RNA-seq data, we found that genotype-by-cell-state heritability was widespread across brain cell types and, on average, exceeded main genetic heritability, indicating that context-dependent genetic effects explain a substantial fraction of heritable expression variation. G×Cell-State heritability was enriched among low-frequency variants and constrained genes, and showed stronger enrichment for functional gene sets and disease heritability than main genetic effects. Using a novel G×Cell-State TWAS framework, we identified 68% more disease-risk genes and recapitulated rare-variant burden-test gene *TREM2*, which was missed by state-agnostic TWAS. In blood, we observed the same direction of enrichment for G×Cell-State heritability at constrained genes, but weaker than in brain, suggesting that cell-state-dependent regulation at constrained genes may be more prominent in brain. Finally, extending scHINT to perturbation screens revealed substantial perturbation-by-cell-state effects, highlighting broad context dependence in transcriptional responses.

## Introduction

Large-scale genome-wide association studies (GWAS) have identified many robust disease-associated loci, but most associated variants lie outside protein-coding regions^1^ and are thought to act through gene-regulatory mechanisms^2-4^. This has motivated extensive efforts to connect GWAS loci to molecular function through expression quantitative trait loci (eQTLs), using approaches such as colocalization^5-7^, transcriptome-wide association studies^8-13^, and Mendelian randomization^14-16^.

However, despite the success of resources such as GTEx^17^ in mapping genetic effects on gene expression across human tissues, many GWAS loci cannot be confidently linked to steady-state gene expression measured in bulk tissues^18,19^.

Disease-relevant regulatory effects that do not colocalize with steady state expression may be weak, context-dependent, or active only in specific cellular environments^20,21^. Bulk-tissue eQTL studies aggregate expression across heterogeneous mixtures of cells and may therefore miss genetic effects that are restricted to rare cell populations or transient cellular states. This limitation is particularly important for complex diseases, where genetic risk is often enriched in regulatory elements active in disease-relevant cell types^22^, and where the same variant may exert different effects depending on cellular identity^23,24^, stimulation^25-29^, development^30-32^, or pathological state^33^. GWAS and bulk eQTL studies also tend to implicate systematically different classes of genes^18^. GWAS loci are enriched near evolutionarily constrained genes^34,18,35^ and key functional annotations, whereas bulk cis-eQTLs are depleted near constrained genes and tend to cluster near transcription start sites^18^. Taken together, these patterns suggest that disease risk may often involve genes under strong functional constraint, where large steady-state regulatory perturbations are selected against and common disease-associated regulatory effects are weak or revealed only under specific cellular contexts^36^.

Single-cell RNA sequencing^37,38^ provides an opportunity to address these limitations by measuring gene expression within individual cells rather than averaged tissues. Population-scale single-cell eQTL studies have demonstrated that genetic effects can be highly cell-type-specific^39-44,22,45^, and methods have been developed to connect cell-type-specific regulation with GWAS through single-cell expression^46-53^. In addition to cell-types, single-cell data can reveal the presence of cellular “states” – transient and continuous gene activations such as cell cycle, hypoxia, or stress response^54^. However, most existing approaches rely on pseudo-bulk profiles, which aggregate cells from the same individual within a cell type^17,55-57^. Although pseudo-bulk analysis improves robustness and reduces technical noise^36,43^, it loses cell-level resolution and cannot directly capture genetic effects that vary across continuous cell states within cell types.

Although single-cell eQTL mapping has been highly informative for identifying regulatory variants, eQTL discovery remains strongly constrained by sample size, effect size and statistical power^58^. In contrast, heritability-based approaches do not require individual variant–gene associations to reach significance and can therefore quantify the aggregate contribution of genetic variation to molecular or complex traits. Heritability partitioning has been widely used in complex-trait genetics to characterize enrichment across functional annotations^2^, tissues^59^, and cell-type-specific regulatory elements^3^. Similarly, prior studies have quantified the heritability of gene expression and estimated the extent to which disease heritability is mediated through genetically regulated expression^55,60^, but these analyses have largely relied on bulk or pseudo-bulk level expression profiles. More recently, Xu et al. introduced scGeneHE^61^, a scalable mixed-model framework that extended SAIGEQTL^62^ for estimating gene expression heritability directly from single-cell count data without requiring pseudo-bulk aggregation. However, no studies have quantified the extent to which gene expression heritability may be explained by genotype-by-cell-state interactions at single-cell resolution.

In this work, we introduce **scHINT** (**s**ingle-**c**ell gene expression **H**eritability estimation with genotype-by-cell-context **INT**eractions), a framework for quantifying cell-level expression heritability. scHINT models genotype × cell-state interactions and partitions expression variance into main genetic, G×Cell-State, individual-level, and cell-level components, enabling estimation of both main genetic effects shared across cells and context-specific genetic effects that vary continuously with cell state. We applied scHINT to 3.8 million cells from six major brain cell types across 584 ROS/MAP individuals^40,63^. Through simulations and empirical analyses, we evaluated the accuracy and robustness of scHINT and generated a high-resolution map of context-dependent genetic regulation in the aging human brain. These analyses showed that cell-state-dependent genetic effects capture a distinct component of expression heritability enriched among low-frequency variants, constrained genes, activation dependent genes, and disease-relevant pathways. We extended scHINT to downstream disease interpretation using a novel G×Cell-State TWAS framework and show that it can link context-dependent regulatory effects to disease-risk genes missed by traditional TWAS. Finally, we demonstrate that scHINT can be generalized to additional settings, including different tissues, interaction with cell-type, disease status and perturbational screens, providing a flexible framework for dissecting context-dependent genetic regulation from single-cell data.

## Results

### Methods overview

We introduce **scHINT**, a scalable statistical framework for estimating gene expression heritability at single-cell resolution while explicitly modeling genotype-by–cell-state interaction effects (**Fig. 1**). In contrast to existing approaches that primarily capture marginal genetic effects within or between discrete cell types, scHINT enables decomposition of heritability into components attributable to shared genetic effects and their modulation across continuous cellular contexts. We assume that cell states are known a priori, for instance through the application of dimensionality reduction methods, and we investigate multiple such in the real data (**Methods**).

**Fig. 1.**
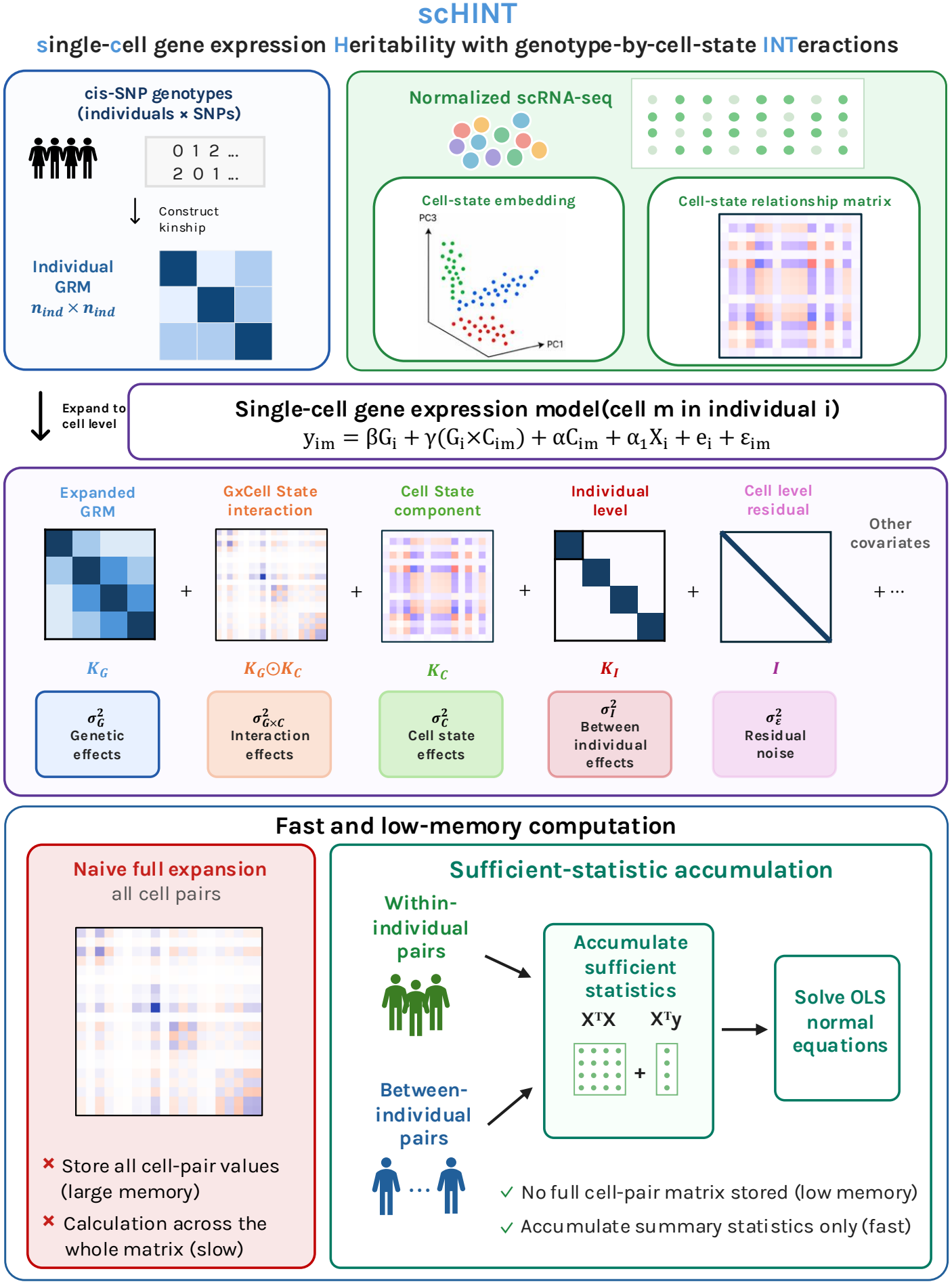
Overview of the scHINT framework. scHINT estimates cell-level gene expression heritability within a cell type by combining cis-SNP genotypes with normalized single-cell expression profiles. Individual genotypes define a genetic relatedness matrix (GRM), which is expanded to the cell level by mapping each cell to its donor; continuous cell-state features (for example, within-cell-type principal components) define a cell-state relationship matrix (CS-RM). Expression of each cell is modelled as the sum of a main genetic effect (G), gene × cell-state interaction effects (G×Cell-State; element-wise product of the expanded GRM and the CS-RM), marginal cell-state effects, effects of other sample- and cell-level covariates, an individual-level effect shared by all cells of a donor, and cell-level residual noise. Expression variance is thereby partitioned into genetic 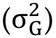, G×Cell-State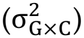, cell-state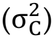, other-covariate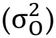, individual-level 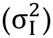 and residual 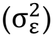components. To avoid constructing the full cell-by-cell matrices, scHINT accumulates sufficient statistics separately from within-individual and between-individual cell pairs and estimates the variance components by solving the resulting Haseman–Elston normal equations.

For a given gene, let *y*_*im*_ denote the normalized expression level measured in cell *m* from individual *i*. We model single-cell expression as

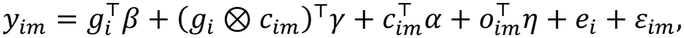

where *g*_*i*_ denotes the cis genotypes for individual *i, c*_*im*_ denotes a vector of cell-state features for cell *m, o*_*im*_ denotes a vector of additional covariates, *β* represents the vector of genetic effect size of cis-SNPs, *γ* represents the vector of G×Cell-State interaction effects, *α* and *η* capture the non-genetic cell-state effects and the effects of other covariates, respectively. The individual-level random effect *e*_*i*_ captures expression variation shared across cells from the same donor, and *ε*_*im*_ denotes cell-level residual noise.

scHINT quantifies these variance components by extending Haseman–Elston regression^64-66^, a moment-based approach for variance component estimation. Specifically, to account for shared genetic background across cells from the same individual, we construct an individual-level genetic relationship matrix (GRM) using cis-SNPs and expand it to the cell level by mapping cell to donor. We standardized each cell state to have a mean of zero and a variance of one. We then constructed the corresponding cell-state relationship matrix (CS-RM), which quantifies the similarity in cell-state activity between pairs of cells. The G×Cell-State interaction component is then modeled by the element-wise product of the expanded GRM and the CS-RM (**Fig. 1**). In our real data analyses, we inferred cell states from normalized single-cell RNA-sequencing profiles using principal component analysis (PCA)^67,68^ or other continuous embeddings^69,70^. In addition, we incorporate two context-level variance components (corresponding to cell and sample level covariates) to account for heteroskedasticity^46,71^, a donor-level variance component to account for variation (variance of *e*_*i*_) shared across cells from the same donor, and a diagonal component capturing residual cell-level noise (variance of *ε*_*im*_). The framework is flexible and can be extended to model other forms of interactions, including cell type or other contextual factors (**Supplementary Fig. 1, Supplementary Note 1**).

A direct HE-regression implementation would require constructing kernels across all cell pairs, leading to approximately *O*(*M*^2^*K*^2^) time complexity and *O*(*M*^2^) memory complexity, where M is the total number of cells and K is the number of cell-state PCs or covariates. This is computationally infeasible for datasets containing millions of cells. To enable scalable estimation, scHINT exploits the repeated-measurement structure of single-cell data and specifies the covariance induced by cells nested within individuals without explicitly forming full cell-by-cell matrices (**Methods and Fig. 1**). By aggregating cell-level quantities into individual-level sufficient statistics, scHINT reduces the dominant computational cost to approximately *O*(*N*^2^*K*^2^) time and *O*(*N*^2^) memory, where N is the number of individuals. Compared with scGeneHE for single-cell-level analysis, scHINT achieved substantially greater computational efficiency, particularly for large datasets, with a 36-fold speedup when analyzing one million cells (**Supplementary Fig. 2**). For uncertainty quantification, scHINT uses a jackknife based on donor-level sufficient summary statistics, allowing standard errors to be estimated without additional computational burden. In contrast, scGeneHE relies on 10-round bootstrap inference, which requires repeated model fitting and approximately 370-fold greater computational cost (**Methods, Supplementary Fig. 2**).

**Fig. 2.**
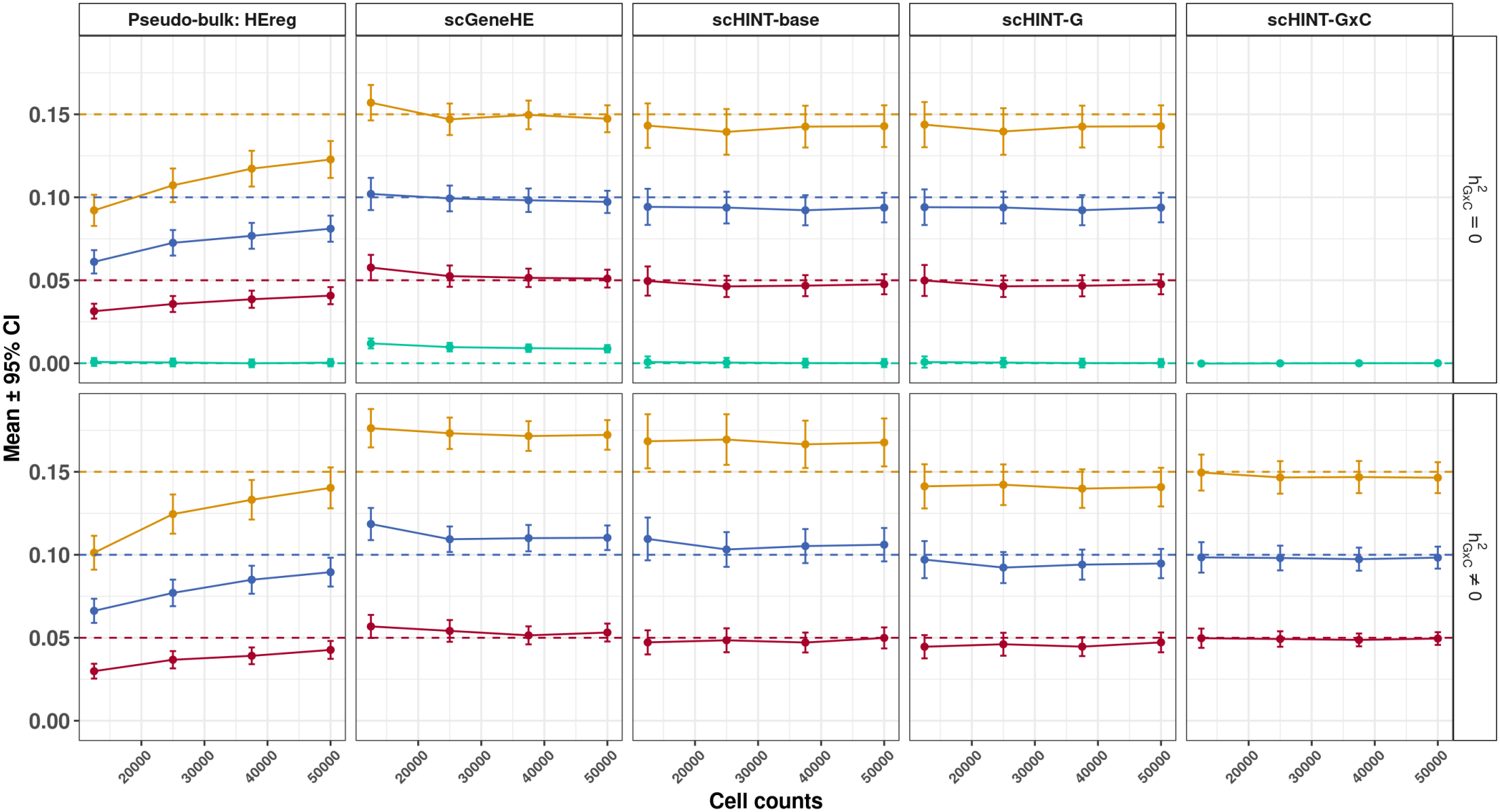
Simulations comparing scHINT with existing estimators. Mean estimates and 95% confidence intervals across 500 simulation replicates for Haseman–Elston regression applied to pseudo-bulk data (Pseudo-bulk: HEreg) and for scGeneHE, scHINT-base (no interaction term), scHINT-G and scHINT-G×C applied to cell-level data. Columns show the methods and rows show simulations with no G×C heritability (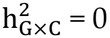; top) or non-zero G×Cell-State heritability (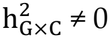 ≠ 0; bottom). The x axis shows the total number of cells in each simulation (500 individuals × 25–100 cells). The y axis shows the estimated main genetic heritability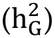, or 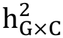 for scHINT-G×C. Coloured points and lines show estimated heritability, and dashed horizontal lines of the same colour mark the simulated true values (gold, 0.15; blue, 0.10; red, 0.05; green, 0). Here the cell-state and individual-level variance components were set to 0 and the mean read count per cell was μ_c_ = 10. Pseudo-bulk HEreg is biased downwards at small cell numbers, scGeneHE and scHINT-base are inflated when G×C effects are present, and scHINT-G and scHINT-G×C are unbiased. See Supplementary Figs. 3–4 for scenarios with non-zero cell-state and covariate variance and a higher read count.

### scHINT shows unbiased estimation of genetic and interaction heritability in simulation

We conducted extensive simulations to evaluate the performance of scHINT across a range of scenarios using real genotype data and realistic expression statistics. For each individual, single-cell gene expression was generated by combining a shared genetic component, a genotype-by–cell state interaction component, and additional individual- and cell-level effects. We systematically varied the proportion of variance explained by the main genetic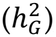, G*×*Cell-State 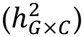, cell-state 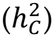 and other-covariate 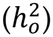 components, as well as the number of cells per individual and the mean read count per cell (**Methods**).

Heritability estimated from single-cell expression is not directly comparable to pseudo-bulk heritability, because the total phenotypic variance at the single-cell level includes both between-individual and between-cell variation. To enable a fair comparison between scHINT and pseudo-bulk analyses, we defined normalized main and interaction heritability as follows:

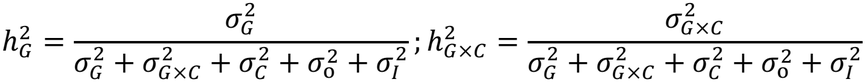

Specifically, the denominator includes variance components corresponding to genetic variation, G×Cell-State interactions, cell state, other covariates and individual-level noise. We excluded cell-level variation from the denominator because these components are not represented in pseudo-bulk expression data and would otherwise make the comparison inconsistent. The resulting estimate can be interpreted as the variance that could be explained in a population where each cell was measured without stochastic error (while still allowing for individual-level environmental variance).

We compared the following four approaches:

1. Haseman-Elston regression^64^ (HEreg) applied to pseudo-bulk expression data.
2. scGeneHE^61^ applied directly to cell-level expression data.
3. scHINT-base, a reduced scHINT model without the genotype-by-cell-state interaction term, applied to cell-level expression data.
4. scHINT, our full model, applied to cell-level expression data and used to estimate both the main genetic component, scHINT-G, and the genotype-by-cell-state interaction component, scHINT-G×C.

Under the null in which the variance of all sources was set to zero, HEreg, scHINT-base, scHINT-G, and scHINT-G×C all produced unbiased estimates centered at zero. In contrast, scGeneHE produced inflated heritability estimates under the null because its estimates are constrained to be non-negative, causing the mean estimate across replications to be significantly greater than zero (**Fig. 2**).

Under the alternative with non-zero variance terms, scHINT consistently produced unbiased estimates of both the main genetic heritability and the G×Cell-State heritability across parameter settings. Each of the other methods produced biased estimates in certain scenarios. First, HEreg applied to pseudo-bulk data showed downward bias when the number of cells per individual was small and approached the true value as the number of cells per individual increased. This attenuation was more pronounced when the mean read count per cell was larger, corresponding to increased cell-level variation that pseudo-bulk analysis cannot explicitly model (**Supplementary Fig. 3-4, Supplementary Table 1**). We provide a theoretical derivation of this phenomenon in the **Supplementary Note 2**. Second, both scHINT-base and scGeneHE operate directly on cell-level data but do not model G×C interactions. As a result, they became inflated when 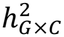 *h*^2^ was non-zero, reflecting the misspecification induced by omitting the interaction variance component. Third, scGeneHE was sensitive to the inclusion of covariates, including cell states and pseudo-bulk gene expression PCs (**Supplementary Fig. 3-4**). This difference is partly due to the distinct definition of heritability used by scGeneHE. Unlike scHINT, scGeneHE estimates two variance components: a genetic component based on the genetic relatedness matrix, *τ*_1_, and a cell-level relatedness component based on the cell relatedness matrix, *τ*_2_. Heritability is then defined as 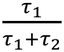 and does not account for all sources of phenotypic variation. As a result, when sample-level covariates explain substantial variation, the remaining variance decomposition can overstate the relative contribution of genetic effects, leading to inflated heritability estimates (relative to the total expression variance). We carried out a wide range of additional simulations demonstrating that scHINT is robust to variation in cell-type proportions^72,73^, expression standardization procedures^74,75^ (**Supplementary Note 3, Supplementary Fig. 5-7, Supplementary Tables 2-3**) as well as unbiasedness for G×Cell-Type heritability estimation (**Supplementary Note 4, Supplementary Fig. 8-9, Supplementary Table 4**).

**Fig. 3.**
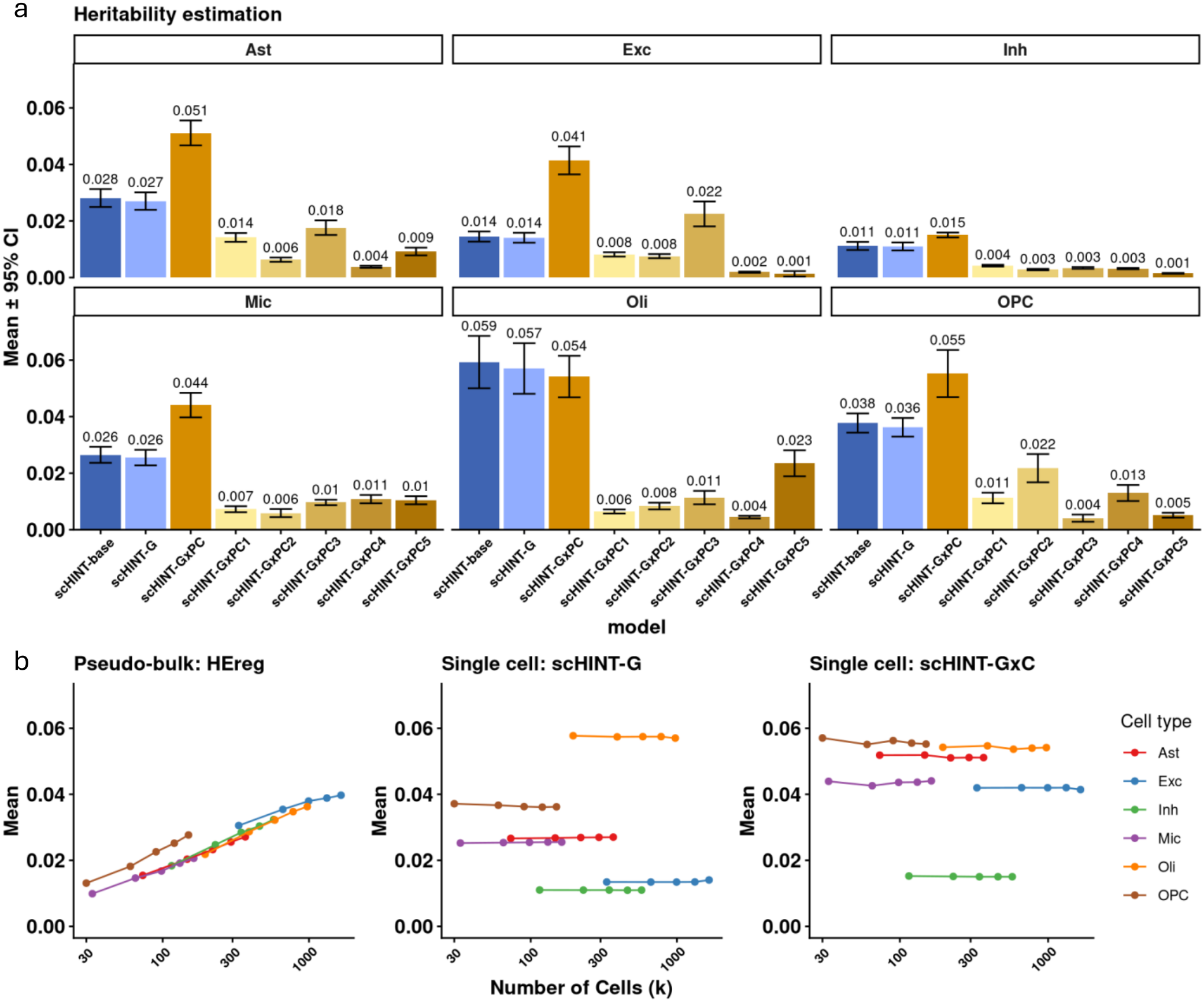
Application of scHINT to ROS/MAP brain single-nucleus RNA-seq data. **a**. Mean heritability estimates and 95% confidence intervals across genes for each major brain cell type, comparing scHINT-base, scHINT-G and scHINT-G× Cell-State (the latter shown as the combined contribution of G×PC1–PC5). b. Mean heritability estimates as a function of the number of cells included in the analysis for pseudo-bulk HE regression, scHINT-G and scHINT-G×Cell-State, shown for each cell type. Pseudo-bulk estimates fall as fewer cells are aggregated, whereas scHINT-G/GxCell-State estimates are stable.

### Single-cell data reveals substantial G×Cell-State interaction heritability

We applied scHINT to the combination of two large brain snRNA-seq datasets from Mathys et al.^63^ and Fujita et al.^40^, both generated from ROS/MAP samples^76,77^. After processing the snRNA-seq data (**Methods**), we focused on six major cell types: excitatory neurons (Exc), inhibitory neurons (Inh), astrocytes (Ast), oligodendrocytes (Oli), oligodendrocyte precursor cells (OPC), and microglia (Mic), which comprise 3.8 million nuclei from 584 individuals. As in the simulation analyses, we benchmarked scHINT against three alternative approaches, HEreg^64^ which was applied to cell-type-specific pseudo-bulk expression, scGeneHE^61^ and scHINT-base, which were applied directly to cell-level expression. Specifically, we used within-cell-type PCA^67^ to infer the cell states and estimate the G×PC heritability from each of the top five PCs jointly (**Supplementary Fig. 10**).

We found that G×C heritability was pervasive across major brain cell types (**Fig. 3a**), indicating that cell-state-dependent genetic effects contribute broadly to single-cell expression variation in the human brain. Across most cell types, the aggregate heritability from the five G×PC components were comparable to, and in several cases exceeded, the main genetic heritability, supporting the importance of context-dependent regulation beyond average genetic effects. For example, excitatory neurons, astrocytes, microglia, and OPCs showed a substantially larger combined G×PC heritability than the main genetic component (192.8%, 88.9%, 69.2% and 52.8% higher respectively). The leading cell-state axis did not necessarily explain the largest interaction heritability. Although PC1 contributed appreciably in some settings (0.0086 on average), other PCs often showed equal or stronger effects (0.0087 on average), such as PC3 in excitatory neurons (0.022) and astrocytes (0.018), PC2 in OPCs (0.022) and PC5 in oligodendrocytes (0.023). Taken together, these results suggest that genetic regulation is modulated by multiple, distinct cell-state axes.

The main genetic and G×Cell-State component heritability estimates were largely independent, with correlation coefficients ranging from −0.022 to 0.031 (**Supplementary Fig. 11**). Consistent with this interpretation, the scHINT-base model produced a slightly higher average estimate than the corresponding scHINT-G (0.0295 versus 0.0284). Moreover, scHINT G×Cell-State heritability was broadly distributed across all bins of scHINT-G heritability (**Supplementary Fig. 12**). Together, these results indicate that direct genetic regulation can be weak even when context-dependent genetic effects remain appreciable (and vice versa). Genes exhibiting this pattern may therefore represent particularly informative candidates for future G×Cell-State eQTL analyses.

We carried out multiple robustness and sensitivity analyses. First, we performed a down sampling analysis to evaluate sensitivity to cell sampling depth. Pseudo-bulk HE regression estimates decreased as fewer cells were used to construct pseudo-bulk expression profiles, consistent with our simulations. In contrast, scHINT estimates remained broadly stable across cell numbers, for both the main genetic (scHINT-G) and G×Cell-State (scHINT-G×C) components within each cell type (**Fig. 3b**). Second, we compared to estimates of main effect heritability from scGeneHE. Because of the computational burden, we first performed a meta-cell analysis and compared the resulting scHINT and scGeneHE estimates. We found that scGeneHE estimates were highly sensitive to covariate adjustment: including 25 pseudo-bulk and five cell-level PCs roughly doubled the mean estimate (0.044 to 0.088), whereas unadjusted scGeneHE estimates were closer to scHINT-G (mean 0.035). This is consistent with our simulations, in which scGeneHE is inflated when covariates explain substantial variance, and is detailed in the **Supplementary Note 5 (Supplementary Fig. 13)**. Thirdly, we demonstrated the robustness of scHINT to alternative cell-state derivation methods, the number of PCs included in the model, the sparsity of read counts, the inclusion of genes to generate cell states, and the interaction with diseases status in **Supplementary Note 6 (Supplementary Fig. 14-21)**.

Finally, complementary to the G×Cell-State analysis, we applied the scHINT-G×Cell-Type model to the same datasets after pseudo-bulk aggregation. To evaluate cell-type-dependent genetic effects, we performed both pairwise analyses between cell types and a joint analysis modeling genetic heterogeneity across all six major cell types (**Supplementary Note 7 and Supplementary Fig. 22-26**). In the joint analysis, G×Cell-Type heritability was on average 0.92 times the main genetic heritability, lower than the corresponding G×Cell-State ratio of 1.72. Thus, cell states modulate genetic effects at least as strongly as discrete cell types.

### G×Cell-State heritability is enriched at constrained genes and low frequency variants

Having established that G×Cell-State heritability is pervasive across brain cell types, we next sought to characterize the genetic architecture and functional relevance of these context-dependent effects.

First, we asked whether genes with higher G×Cell-State heritability exhibit unique patterns of evolutionary constraint (**Fig. 4a, Supplementary Table 5**). Across all six cell types, main genetic heritability showed negative correlations with multiple constraint annotations (with the lowest cor = - 0.15, p-value = 1.50 *×*10^-54^), implying that genes with more/larger main genetic effects were less likely to be under selection. These results are consistent with prior findings from analyses of bulk and pseudo-bulk eQTLs^78-80^. In contrast, many G×Cell-State components showed positive correlations with the same constraint metrics (with the highest cor = 0.13, p-value= 1.12*×*10^-36^). We further examined the relationship between evolutionary constraint and the proportion of total heritability attributable to G and G×Cell-State. This produced consistent but stronger correlations (with the highest cor = 0.17, p-value= 1.49*×*10^-68^) (**Supplementary Fig. 27, Supplementary Table 6**). A larger absolute G×Cell-State heritability at more constrained genes could be consistent with a larger mutational target size for interactions. These opposing correlation directions suggest that context-dependent genetic effects preferentially concentrate in genes under stronger selective constraint, whereas main genetic effects are relatively depleted among constrained genes. We also examined model with additional G×PC terms (10, 15 and 20 PCs). Overall, the model with 5 G×PC terms shows the highest correlation with constraint scores, although the optimal number depends on cell type and read depth. Interestingly, excitatory and inhibitory neurons still retained a positive correlation with more PCs or after down-sampling (**Supplementary Table 7, Supplementary Note 6, Supplementary Fig. 18**).

**Fig. 4.**
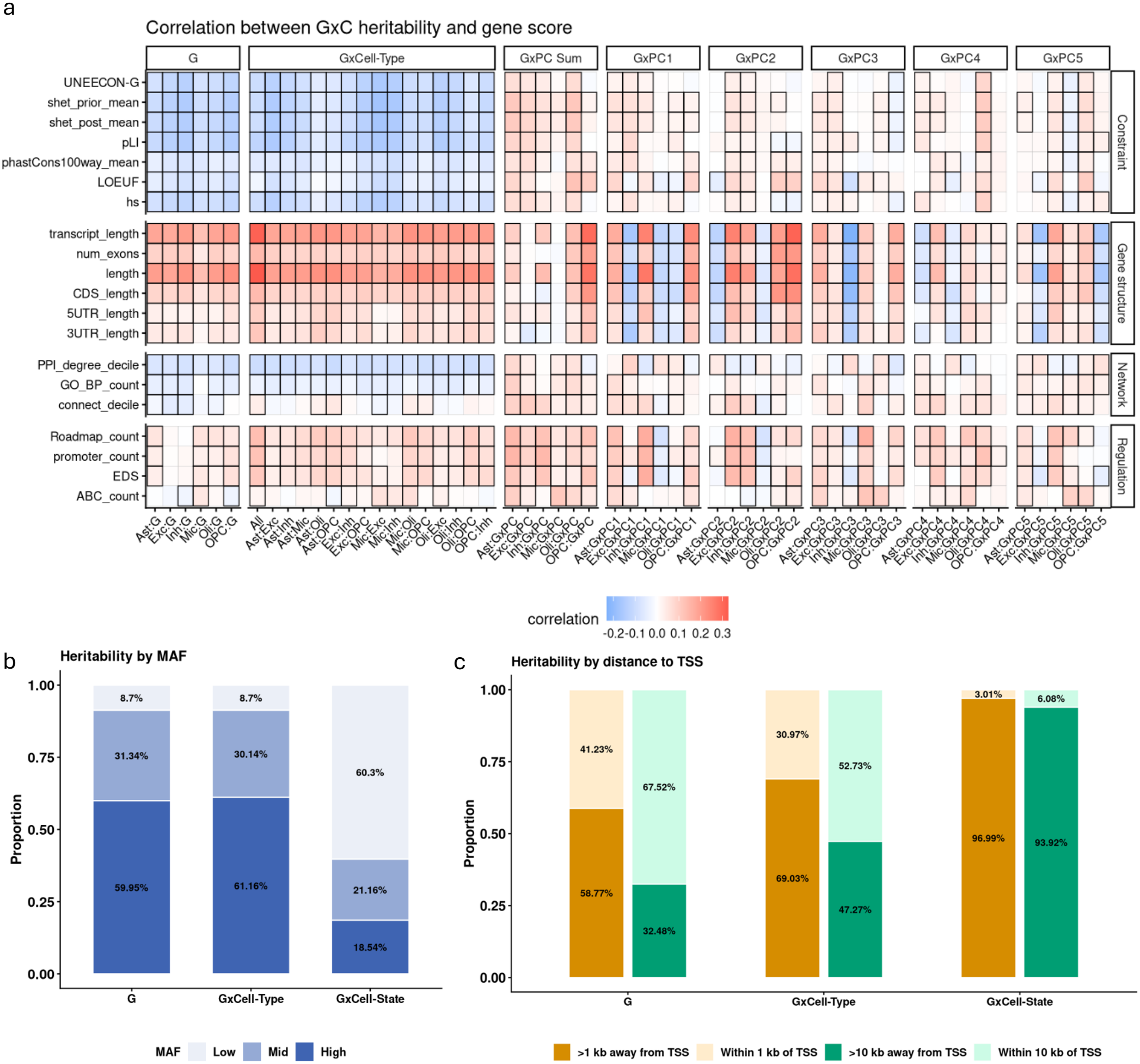
G×Cell-State heritability is enriched at constrained genes and low-frequency variants. **a**, Correlation between gene-level heritability estimates from scHINT models and functional, regulatory, evolutionary-constraint and network-based gene scores across the major brain cell types. Rows are gene scores grouped by category; columns are heritability components for each cell type and cell-state PC. Colour indicates the standardized regression coefficient, and black borders mark significant correlations (FDR < 0.05). **b**, Proportion of heritability explained by variants in each minor allele frequency (MAF) bin for the main genetic (G), G×Cell-Type and G×Cell-State components. **c**, Proportion of heritability explained by variants within or outside windows around the transcription start site (TSS) for the same components.

We also examined associations between heritability components and regulatory, network, and gene-structure annotations. All three components, G, G×Cell-Type, and G×Cell-State were positively correlated with regulatory features such as promoter counts, suggesting greater heritability for genes with more complex regulatory architecture. For network annotations, G and G×Cell-Type were negatively correlated, whereas G×Cell-State was positively correlated, potentially reflecting stronger constraint on stable genetic effects but greater flexibility for state-dependent regulation. Gene-structure features, including gene length, were positively correlated with G and G×Cell-Type, while their associations with G×Cell-State varied in direction, indicating a more context-specific relationship.

Motivated by the gene level correlation results, we next examined how heritability is partitioned by minor allele frequency (MAF), where components under stronger directional or stabilizing selection are expected to have a larger contribution from lower frequency variants. Strikingly, low-frequency variants (MAF between 1% and 5%) accounted for 60% of G×Cell-State heritability, while high-frequency variants (MAF >20%) explained only 19% on average. In contrast, main genetic heritability was primarily driven by common variants (**Fig. 4b**): low-frequency variants accounted for only 9% of main genetic heritability, while high-frequency variants explained 60% on average. A similar pattern was observed for G×Cell-Type heritability, where low-frequency variants explained 9% and high-frequency variants explained 61%. Similar frequency partitions were observed at highly and lowly constrained genes (bottom and top 20% of pLI), although highly constrained genes exhibited a higher proportion of G×Cell-State heritability overall (**Supplementary Fig. 28**). Together, these results reveal a striking shift in allele-frequency architecture, with genetic effects that vary across continuous cell states disproportionately driven by low-frequency variants, particularly among constrained genes, in contrast to the common-variant-dominated architecture of average genetic effects and cell-type-level interactions.

Finally, we quantified the distribution of heritability with respect to physical distance from each gene. It is well known that eQTLs tend to be tightly clustered near transcription start sites (TSSs)^81,82^. Interestingly, G×Cell-State heritability exhibited distinct enrichment patterns with respect to gene distance. Promoter regions, defined as a 2-kb window around the TSS, and promoter-proxy regions, defined as a 20-kb window around the TSS, explained 41% and 68% of main genetic heritability, and 31% and 53% of G×Cell-Type heritability, respectively, but accounted for only 3% and 6% of G×Cell-State heritability (**Fig. 4c**). Because GWAS-associated variants tend to lie farther from the nearest TSS than conventional eQTLs, this pronounced depletion of promoter-proximal effects suggests that G×Cell-State regulation may follow a more distal, GWAS-like regulatory architecture.

### G×Cell-State heritability shows more significant enrichment for functional gene sets

Differences in the selective constraint and regulatory landscapes of genes likely reflect the functional roles of G and G×Cell-State. Individual G×PC components differed in their correlation with constraint, from positive to slightly negative. To gain a better understanding of the heritability estimation and to explore functional disparities between G and G×Cell-Context, we next evaluated whether G×Cell-State/Type heritability was enriched for a broad definition of functional gene sets (**Methods**). Compared with the main genetic heritability, G×Cell-State heritability was significantly associated with 7.12-fold more unique functional gene sets (**Fig. 5a, Supplementary Table 8)**. Across all major brain cell types, G×Cell-State components consistently identified many more significant gene-set enrichments than the corresponding main genetic component. For example, in excitatory neurons, G×PC2 and G×PC4 were associated with 572 and 385 significant gene sets, respectively, whereas the main genetic component was associated with only 43. Similar patterns were observed in other cell types. We likewise observed more significant G×Cell-Type heritability enrichments than main heritability. These differences indicate that genes with higher G×Cell-Context heritability are more strongly connected to annotated biological pathways and functional gene programs.

**Fig. 5.**
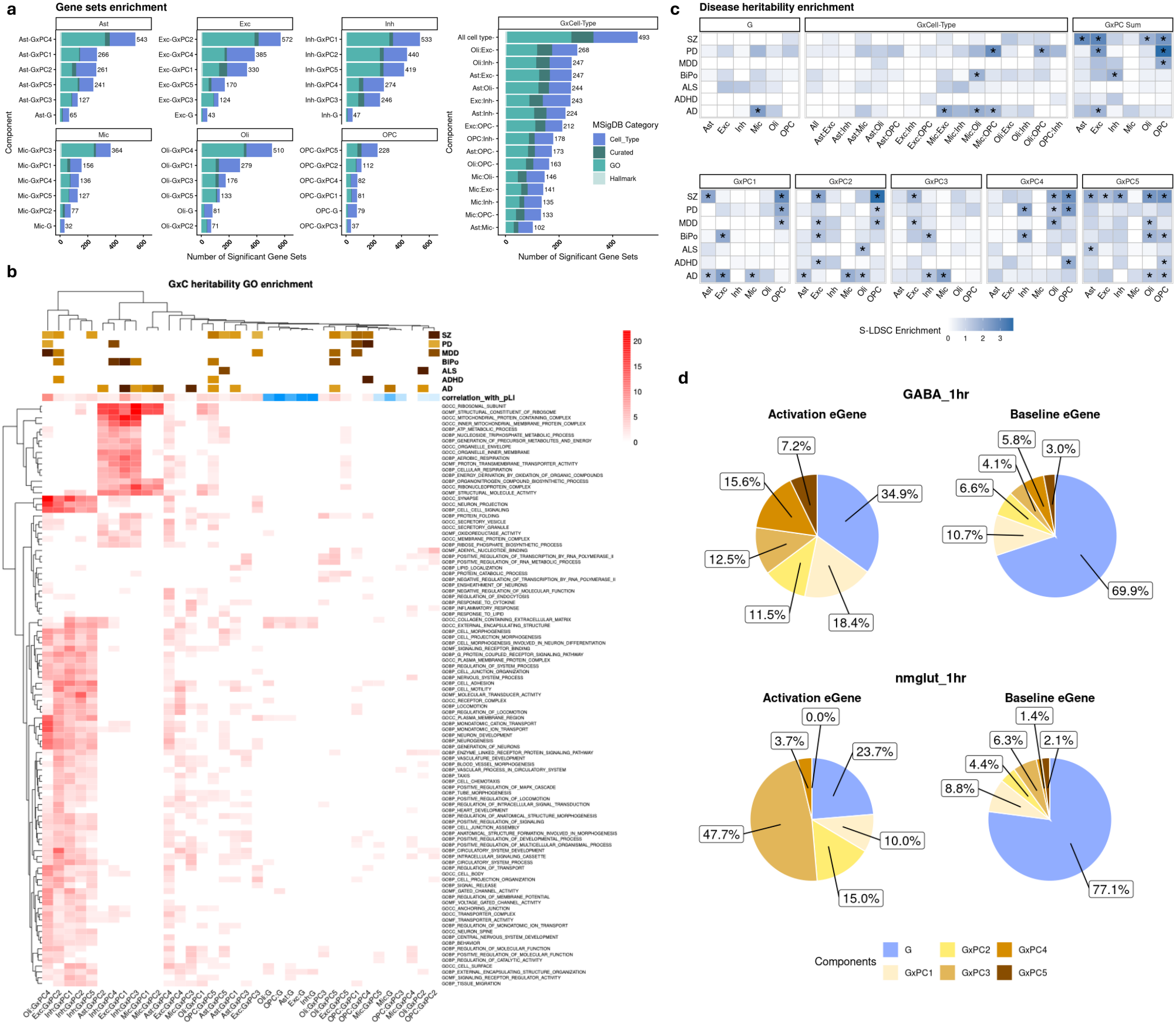
Functional and disease enrichment of G×Cell-State heritability. **a**, Gene-set enrichment of scHINT heritability estimates across MSigDB collections. Bars show the number of significant gene sets (FDR < 0.05) for each cell type and heritability component, stratified by gene-set collection. **b**, Bi-clustered heatmap of Gene Ontology (GO) enrichment across G×PC components. Rows are GO terms and columns are cell-type-specific G×PC components; color indicates enrichment significance (-log10 P), and rows and columns were hierarchically clustered. Top annotations show disease heritability enrichment and the correlation of each component with pLI. **c**, Stratified LD score regression of disease heritability enrichment for the top 300 genes ranked by each heritability estimate (SNPs within 100 kb of the TSS). Rows are brain-related diseases (ADHD, bipolar disorder, MDD, schizophrenia, ALS, Alzheimer’s disease, Parkinson’s disease) and columns are component-specific gene annotations; color indicates enrichment magnitude and asterisks mark significant enrichment (FDR p-value < 0.05). **d.** Relative contribution of heritability components (main genetic, G, and G×PC1–PC5) estimated by scHINT in ROS/MAP neurons for baseline eGenes and stimulation-specific eGenes identified in iPSC-derived neurons by Liang et al. after 1 h of KCl stimulation (top. Inh, bottom. Exc).

To identify shared and unique biological patterns across G×Cell-State components, we bi-clustered heritability components and Gene Ontology terms according to enrichment significance (**Fig. 5b**). We then annotated each heritability component by its correlation with pLI, linking functional enrichment patterns to evolutionary constraint. The resulting heatmap revealed several coherent patterns suggestive of shared regulatory modules. Module 1 comprised Oli: G×PC4, Exc: G×PC2, and Inh: G×PC1/2/5. These components showed broad enrichment across GO-term clusters related to neuronal development and differentiation, such as neurogenesis, generation of neurons, and nervous-system processes; ion-channel activity, including gated and voltage-gated channels; and receptor-mediated signaling, including signaling-receptor binding and G-protein-coupled receptor pathways. Module 2 comprised Ast: G×PC2, Inh: G×PC3/4, Exc: G×PC1, and Mic: G×PC1/2. It showed a largely complementary GO-term enrichment pattern including ribosome-related functions, such as ribosomal subunits and structural constituents of the ribosome, and energy-metabolism pathways, including ATP metabolism and energy derivation through oxidation. The remaining G×Cell-State components generally showed weaker or slightly negative correlations with pLI, fewer significant GO enrichments, and less clearly module-specific patterns. Nevertheless, some captured distinct biological processes not represented in the two major modules. For example, cytokine- and lipid-response pathways were not prominent in either major module but were enriched for Ast: G×PC4 and Mic: G×PC4. In contrast, the main genetic heritability components showed substantially weaker GO enrichment overall, with many terms displaying near-null signals. Together, these results indicate that G×Cell-State heritability captures heterogeneous but coherent biological programs that are less apparent from main genetic heritability alone.

### Genes with G×Cell-State heritability are enriched for disease heritability

We next evaluated whether context-dependent expression heritability captures disease-specific genetic signals. We used stratified linkage disequilibrium score regression (S-LDSC^2^) to evaluate disease heritability enrichment of SNPs near the top 300 most heritable G or G×C genes (**Methods**) and seven neuropsychiatric and neurodegenerative traits: attention deficit hyperactivity disorder (ADHD)^83^, bipolar disorder (BiPo)^84^, major depressive disorder (MDD)^85^, Schizophrenia (SZ)^86^, and neurodegenerative disease, i.e., amyotrophic lateral sclerosis (ALS)^87^, Alzheimer’s disease (AD)^88^, and Parkinson’s disease (PD)^89^ (**Supplementary Table 9**).

In total, 53 G×PC components showed significant disease-specific enrichment (with FDR p value < 0.05, **Fig. 5c, Supplementary Table 10**). Notably, Exc G×PC2, Inh G×PC5, and Oli G×PC4, all assigned to Module 1, showed significant enrichment for schizophrenia (SZ) heritability (Fig. 5b). Module 1 was enriched for GO terms related to neuronal development and differentiation, consistent with biological processes previously implicated by genetic studies of SZ. Large-scale schizophrenia GWAS and rare-variant studies have particularly implicated excitatory and inhibitory neurons^86^, postsynaptic signaling, neurotransmitter receptors, synaptic complexes^90^ and voltage-gated calcium-channel genes^91^.

Alzheimer’s disease showed prominent enrichment in microglia (GxPC1/2/3), consistent with extensive genetic and functional evidence implicating immune and microglial mechanisms in AD^92^. Mic:G×PC3 was uniquely enriched for lipid-response and inflammatory pathways, both of which are highly relevant to AD because microglia regulate lipid handling, plaque-associated immune responses, and phagocytic clearance in the diseased brain^93,94^. Interestingly, AD enrichment was also observed more broadly across other cell types; for example, the Module 2 components Ast: G×PC2, Exc: G×PC1, Inh: G×PC3, and Mic: G×PC1 were all significantly enriched for AD heritability (**Fig. 5b**). This module was characterized by pathways related to energy metabolism, secretion and vesicle biology, consistent with established roles for mitochondrial dysfunction, synaptic degeneration and vesicular trafficking in AD pathogenesis^95^. These results suggest that AD-associated genetic effects are distributed across both microglial immune–lipid programs and broader neuronal and glial processes.

In contrast, functional annotations defined from top main effect heritable genes identified only one significant disease enrichment, involving AD in microglia. G×Cell-Type interaction heritability captured broader disease-relevant signal, with six significant enrichments across disease traits and cell-type pairs (**Fig. 5c**), including multiple AD enrichments involving microglia × excitatory neurons, oligodendrocytes, and OPCs, as well as bipolar disorder enrichments involving oligodendrocyte and microglia interactions However, G×Cell-Type enrichment remained substantially more limited than G×Cell-State enrichment, with 6 versus 53 significant associations. These results suggest that cell-type interactions capture disease-relevant effects beyond main genetic regulation while continuous cell states reveal a broader layer of context-dependent genetic regulation.

### Activation-dependent eQTLs reproduce G×Cell-State heritability

A challenge for inference from data-derived cellular states is their lack of direct biological interpretability. To externally replicate our G×Cell-State heritability estimates and provide them interpretable context, we turned to the recent study of neuronal activation eQTLs by Liang et al^27^. In that study, excitatory (Glut+) and inhibitory (GABA+) neurons differentiated from 100 human induced pluripotent stem cell lines were exposed to membrane-depolarizing potassium chloride, and eGenes were identified at baseline and after 1 or 6 hours of stimulation. We thus compared the relative contributions of heritability components estimated in neurons in our data to the baseline eGenes and stimulation-specific eGenes independently identified in Liang et al (**Fig. 5d and Supplementary Fig. 29**). The proportion of heritability attributable to G×C effects differed substantially between the two groups. In excitatory neurons characterized by a low-neurofilament program (nmGlut), G×Cell-State effects explained 76.31% of the total heritability for eGenes specific to the 1-hour stimulation condition, compared with only 22.88% for baseline eGenes. This difference was driven primarily by PC3, which explained 47.7% of the total heritability among stimulation-specific eGenes but only 6.3% among baseline eGenes (difference P=1.97×10^−7^). Indeed, Exc:G×PC3 was enriched for pathways related to synaptic signaling, neurotransmitter release, vesicle-mediated transport, and synaptic plasticity, indicating that this component captures a stimulation-dependent neuronal regulatory program. These results support the interpretation that scHINT captures PC3-dependent genetic regulation that becomes more prominent during neuronal activation. In inhibitory (GABA+) neurons, all five G×PC components explained significantly more heritability for genes specific to the 1/6-hour stimulation condition than for baseline eGenes. All five components were also assigned to modules 1 and 2, which were broadly enriched for diverse GO terms, suggesting that these cell-state PCs capture biologically important regulatory programs in inhibitory neurons.

### G×C-TWAS identifies novel risk genes for neurodegenerative diseases

Transcriptome-wide association studies (TWAS) provide a complementary strategy for connecting GWAS loci to their target genes. However, conventional TWAS models are typically trained on bulk or pseudo-bulk expression and therefore capture main genetic effects while ignoring genetic effects that vary across cellular states. Given the strong disease relevance of G×Cell-State heritability, we reasoned that extending TWAS to model context-dependent genetic regulation could provide a powerful framework for linking cell-state-dependent regulatory effects to GWAS signals. We therefore evaluated whether incorporating G×Cell-State effects can improve cell-level expression prediction and identify disease-risk genes that are missed by conventional main-effect TWAS.

To reduce the computational burden, we aggregated cells into approximately 10,000 metacells per cell type^96^. For each gene within each cell type, we trained two classes of cell-level expression prediction models (**Methods**): a Baseline model using elastic net regression to predict cell-level expression directly from genotype alone and a G×Cell-State model using a two-step interaction to generate two sets of weights that represent genetic components and G×Cell-State components. We additionally performed conventional pseudo-bulk TWAS within each cell type and G×Cell-Type TWAS across the six cell types (**Supplementary Note 8**). For each target cell type in G×Cell-Type TWAS, the main-effect and cell-type-specific interaction weights were summed to obtain prediction weights.

Importantly, cell-level and sample-level models were trained and evaluated under different data settings. Cell-level prediction is inherently more challenging because individual-cell expression contains substantially greater residual variation as well as donor-level error. We therefore assessed cell-level prediction performance using mixed-effect models that account for donor-level random effects across cells. A prediction model was considered significant if the corresponding prediction coefficient was positive and nominally significant, defined as coefficient > 0 and P < 0.05. Prediction performance for pseudo-bulk models was assessed using Pearson correlation, with models considered predictable at r > 0.1 and P < 0.05 (**Methods and Supplementary Note 8**).

At the cell level, the Baseline model, G×Cell-State main-effect weights, and G×Cell-State interaction weights yielded 12,781, 13,839, and 12,748 significant prediction models, respectively, across the six cell types. Combining the main and interaction components increased the number of significant G×Cell-State prediction models to 15,854, indicating that incorporating cell-state-dependent genetic effects improves cell-level gene expression prediction (**Supplementary Fig. 30**). At the sample level, conventional pseudo-bulk TWAS and G×Cell-Type TWAS produced 20,114 and 23,528 significant prediction models, respectively.

We then evaluated whether scHINT heritability estimates can forecast the empirical predictive model accuracy (as has previously been observed for heritability estimates and polygenic scores^97^). As expected, genes with higher scHINT-estimated main genetic heritability were more likely to yield significant base-model predictions (**Fig. 6a**). However, G×Cell-State interaction heritability estimates identified additional predictive components beyond the main genetic effect. For example, when genes were binned by scHINT G×PC1 heritability, the proportion of significant G×PC1 prediction weights and mean prediction accuracy increased with G×PC1 heritability (**Fig. 6b** and **Supplementary Fig. 31-32**). In contrast, prediction components for main effect or other G×PC showed lower significance rates, suggesting that the interaction signal is concentrated in specific cell-state axes rather than uniformly distributed with main effect or other PCs.

**Fig. 6.**
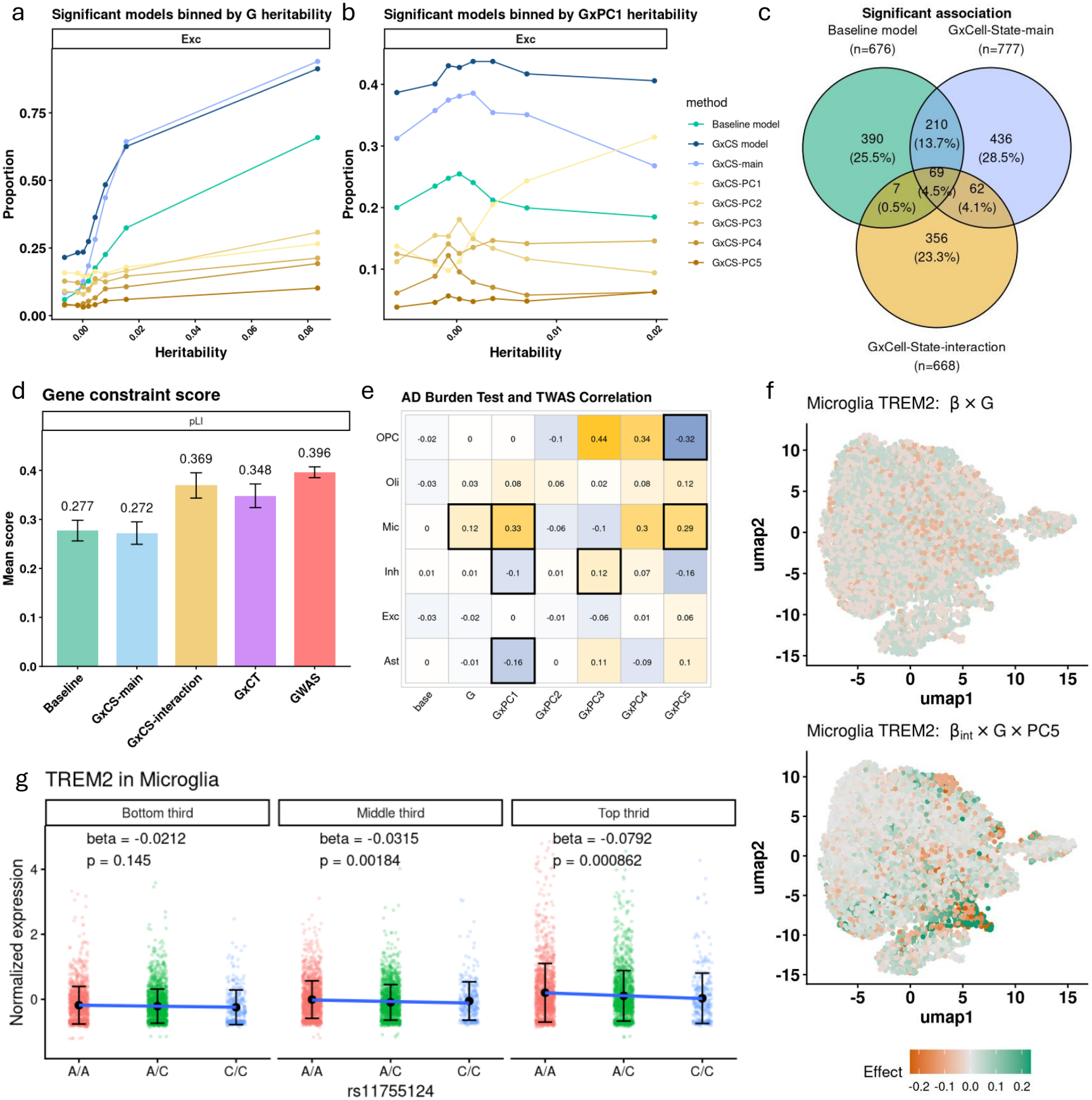
G×Cell-State TWAS identifies context-dependent disease genes. **a**,**b**, Proportion of significant prediction models in excitatory neurons across genes binned by main genetic heritability (**a**) or G×PC1 heritability (**b**), comparing the baseline model with the two-step G×Cell-State models (main genetic weights, G×PC1–5 weights and combined weights). **c**, Overlap of significant gene–trait associations identified by the baseline model, the G×Cell-State main genetic weights and the G×Cell-State interaction weights; n is the number of significant gene–trait–model pairs and overlaps are shared gene–trait–cell-type pairs. **d**, Mean pLI constraint score (higher, more constrained) of genes identified by the baseline model, the main genetic weights, the G×Cell-State interaction weights, the G×Cell-Type model and the GWAS-nearest genes; error bars show 95% confidence intervals. **e**, Correlation between Alzheimer’s disease gene-burden test z scores and TWAS z scores across cell types and model components; black borders mark significant correlations (P < 0.05). **f**, UMAP of microglia coloured by the *TREM2* genetic effect of rs11755124 from the main genetic model (β × G) and from the G×PC5 interaction model (β_G*×*PC5_ × G × PC5). **g**, *TREM2* expression by rs11755124 genotype within PC5-defined tertiles of microglia; effect sizes and P values are from mixed-effects eQTL models fitted within each tertile with donor as a random effect.

Having established that G×Cell-State interaction models replicated in held-out samples and tracked with heritability, we tested them for association with GWAS data from 7 neuropsychiatric disorders (**Fig. 6c**). Using the predictable models, a total of 676 significant associations were detected by the baseline model, 777 by the G×Cell-State model main weights, and 668 by the G×Cell-State model interaction weights. A substantial fraction (357) was unique to the G×Cell-State model interaction weights, indicating that interaction-based TWAS captures regulatory effects missed by models based only on average expression. Traditional pseudo-bulk TWAS and G×Cell-Type TWAS models identified 679 and 917 significant associations, with 447 unique to the G×Cell-Type model (**Supplementary Fig. 33 and Supplementary Tables 11-12**). Together, these results show that both discrete cell-type interactions and continuous cell-state interactions can reveal disease-relevant genetic regulation beyond conventional average-expression models.

Genes identified by G×Cell-State and G×Cell-Type TWAS models also showed stronger evolutionary constraint than other models, with mean pLI scores (higher, more constrained) of 0.28 for baseline models, 0.27 for G×Cell-state main weights, 0.35 for G×Cell-Type models, 0.37 for G×Cell-State models, and 0.40 for GWAS nearest genes (**Fig. 6d**). This pattern suggests that integration of state-dependent and cell-type dependent genetic regulation into GWAS can highlight disease-relevant genes under stronger selective constraint.

### G×C-TWAS implicates rare-variant burden test genes

Although traditional TWAS nominates genes that are enriched for likely causal genes^98^, they are still often discordant with genes identified by rare-variant tests^99,100^. One possible explanation is that conventional expression prediction models miss disease-relevant regulatory mechanisms that act only in specific cellular contexts (or nominate broadly active regulatory mechanisms that are correlated but not causal for disease^101^). We therefore evaluated whether G×Cell-State TWAS better aligns with rare-variant burden-test results than conventional TWAS models. Across cell types and model components, G×Cell-State TWAS components showed stronger correlations with AD gene-burden test results than either the base model or main-effect TWAS components. For example, in microglia, the G×PC1 and G×PC5 TWAS z-scores showed nominally significant correlations of 0.33 (p-value =0.014) and 0.29 (p-value =0.029) respectively, with AD gene-burden test z-scores. In contrast, the corresponding correlation was 0 (p-value =1) and 0.12 (p=0.236) for the baseline TWAS model and main-effect TWAS component (**Fig. 6e**). These patterns suggest that G×Cell-State TWAS may capture disease-relevant regulatory effects that conventional TWAS models miss, particularly for constrained genes whose genetic effects emerge only in specific cellular states.

We highlight examples where G×Cell-State-TWAS uniquely implicated genes that were also significant rare burden genes. *TREM2* provides an illustrative example. *TREM2* plays a central role in microglial biology and AD risk^102,103^, and rare-variant burden tests have repeatedly identified *TREM2* as an AD risk gene^88,104^. However, the gene expression of *TREM2* was not predictable by either the cell-level base-TWAS model (prediction p-value = 0.42) or the traditional pseudo-bulk TWAS model (prediction p-value = 0.15). In contrast, the G×PC5 component significantly predicted *TREM2* expression in microglia (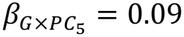 and p-value = 8.1 *×*10^-11^) and showed a significant AD gene–trait TWAS association (p-value = 1.8 *×*10^-4^) mediated by the G×PC5 weights.

While TWAS implicates genes, it does not identify the putative causal variant. We thus performed SuSiE fine-mapping^105^ at the *TREM2* locus for AD GWAS. We identified rs11755124 as a fine-mapped variant with highest PIP (0.96) and genome-wide significant AD association (p-value = 2.38 *×*10^-11^). In a mixed-effect G×C eQTL analysis, rs11755124 showed a nominal main eQTL effect on *TREM2* expression (*β* = −0.043 and p-value = 0.0077) and highly significant GxPC5 interaction effect (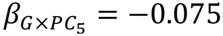 and p-value = 5.13 *×*10^-13^). Mapping these effects onto the microglial UMAP showed that the main genetic effect (*β × G*) had little spatial structure, whereas the G×PC5 interaction 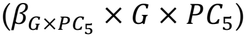 localized the genetic effect of rs11755124 to a distinct region of the microglial state space (**Fig. 6f**). In addition, we stratified microglia into PC5 tertiles. The rs11755124 genotype effect on *TREM2* expression was not significant in the first PC5 tertile (*β* = −0.021 and p-value = 0.14), but became more significant in the middle PC5 tertile (*β* = −0.032 and p-value = 0.0018) and reached its strongest effect in the top PC5 tertile (*β* = −0.079 and p-value = 0.00086, **Fig. 6g**). Together, these analyses show that G×C-TWAS can identify state-dependent genetic regulation that main-effect TWAS does not capture and that are enriched for known rare-variant burden genes.

### Blood cell types show weaker but independently validated G×Cell-State heritability

Having observed substantial G×Cell-State heritability across major brain cell types, we next asked whether context-dependent genetic effects were present in blood. We applied scHINT to the OneK1K single-cell RNA-seq dataset, focusing on 15 cell types spanning five major immune lineages: CD4 T cells, CD8 T cells, B cells, natural killer (NK) cells and monocytes.

Overall, G×Cell-State heritability was lower in blood than in brain, although its magnitude varied considerably across immune cell types (**Supplementary Fig. 34**). Larger G×Cell-State contributions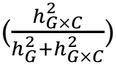 were observed in more differentiated or dynamically regulated populations, including CD8 effector memory T cells (20.82%), intermediate B cells (43.47%) and proliferating NK cells (73.32%). By contrast, naive populations showed particularly modest contributions from state-dependent genetic effects, with G×Cell-State accounting for 4.97%, 8.59% and 13.14% of heritability in naive CD4 T cells, naive CD8 T cells and naive B cells, respectively. These differences suggest that state-dependent genetic regulation is relatively limited in transcriptionally quiescent naive populations but becomes more prominent in differentiated or activated immune states.

We next examined the relationship between genetic regulation and gene constraint (**Supplementary Fig. 35-36, Supplementary Table 6**). In OneK1K, main genetic heritability and the proportion attributable to main genetic effects were negatively associated with gene constraint (mean coefficients = −0.061 and −0.031, respectively), consistent in direction but weaker than the corresponding associations in ROS/MAP brain cells (−0.100 and −0.101). The relationship between absolute G×Cell-State heritability and constraint was considerably weaker and heterogeneous across blood cell types (mean coefficient = 0.003 and 60.0% positive), in contrast to the more consistently positive association observed in brain (mean coefficient 0.054 and 90.5% positive). The proportion of genetic heritability attributable to G×Cell-State was consistently positively associated with constraint across blood cell types (mean coefficient = 0.031 and 99.0% positive) and brain cell types (mean coefficient = 0.101 and 100% positive). Thus, although state-dependent genetic effects are weaker overall in blood, they remain relatively enriched among constrained genes when considered relative to total genetic regulation.

To test whether the G×Cell-State component captured biologically meaningful immune-state-dependent regulation, we analyzed an independent CD4 T-cell activation eQTL dataset^29^. Activation-dependent eGenes consistently showed a larger contribution from G×Cell-State effects, particularly in memory T-cell populations (**Supplementary Fig. 37)**. At day 5 of activation, the G×Cell-State fraction increased from 1.7% among baseline eGenes to 2.9% among activation-dependent eGenes in naive CD4 T cells, but from 4.9% to 20.2% in CD4 central memory T cells and from 4.8% to 20.4% in CD4 effector memory T cells. The stronger enrichment in memory populations and at later activation time points supports the interpretation that the cell-state PCs capture transcriptional programs along which genetic effects are modulated during immune activation.

Several biological and technical factors could contribute to the lower G×Cell-State heritability observed in blood. OneK1K expression profiles are substantially sparser than ROS/MAP brain profiles (∼90% versus ∼60% zero measurements). However, downsampling ROS/MAP by restricting library size or artificially increasing sparsity produced little change in G×Cell-State estimates (**Supplementary Note 6, Supplementary Fig. 20**), arguing against sparsity alone as an explanation. Genotype resolution also differs between datasets: ROS/MAP uses whole-genome sequencing, whereas OneK1K relies on SNP-array genotypes followed by TOPMed imputation^106,107^ and contains limited information for low-frequency variants, which contribute disproportionately to G×Cell-State heritability in brain. Differences between single-cell RNA-seq and single-nucleus RNA-seq may additionally affect the genes and regulatory programs that can be detected. Consistent with this possibility, genetic heritability showed opposite relationships with gene length in the two datasets (**Supplementary Fig. 35a**), and snRNA-seq is known to preferentially capture longer transcripts.

Nonetheless, the persistence of these differences across robustness analyses, together with the activation-eQTL results, suggests that they are unlikely to be entirely technical. Instead, the weaker G×Cell-State component in blood may also reflect genuine differences in the architecture of context-dependent genetic regulation between immune and brain cell populations.

### scHINT generalizes to perturbation single-cell data

Finally, to experimentally validate our findings of broad cell state dependence, we extended scHINT to data from perturbational screens. In this setting, we define perturbation “heritability” as the proportion of expression variance explained by perturbation effects in perturbed cells. This extension allows scHINT to quantify both the average transcriptional response to a perturbation and the extent to which this response changes across continuous cell states or discrete cell types.

We first applied this framework to a large genome-wide perturb-seq data set from K562 cells^108^, which contains 2.06 million cells and 7,189 gene perturbations. Across response genes, the summed perturbation-by-cell-state component explained about 4-fold expression variance than the main perturbation component, indicating that perturbation effects often vary across transcriptional states rather than acting uniformly across cells (**Fig. 7a**). To prevent the PCs from capturing the perturbation itself, we also derived them from non-targeted control cells and projected this PC space onto the targeted cells. Heritability estimates obtained with control-derived PCs were highly concordant with those obtained with PCs derived from targeted cells (Pearson r = 0.992 for the main perturbation effect and r = 0.964 for the summed perturbation × cell-state component; **Supplementary Fig. 38**), indicating that PCs derived from control and from targeted cells capture consistent cellular programs and that the choice between them has little influence on the results.

**Fig. 7.**
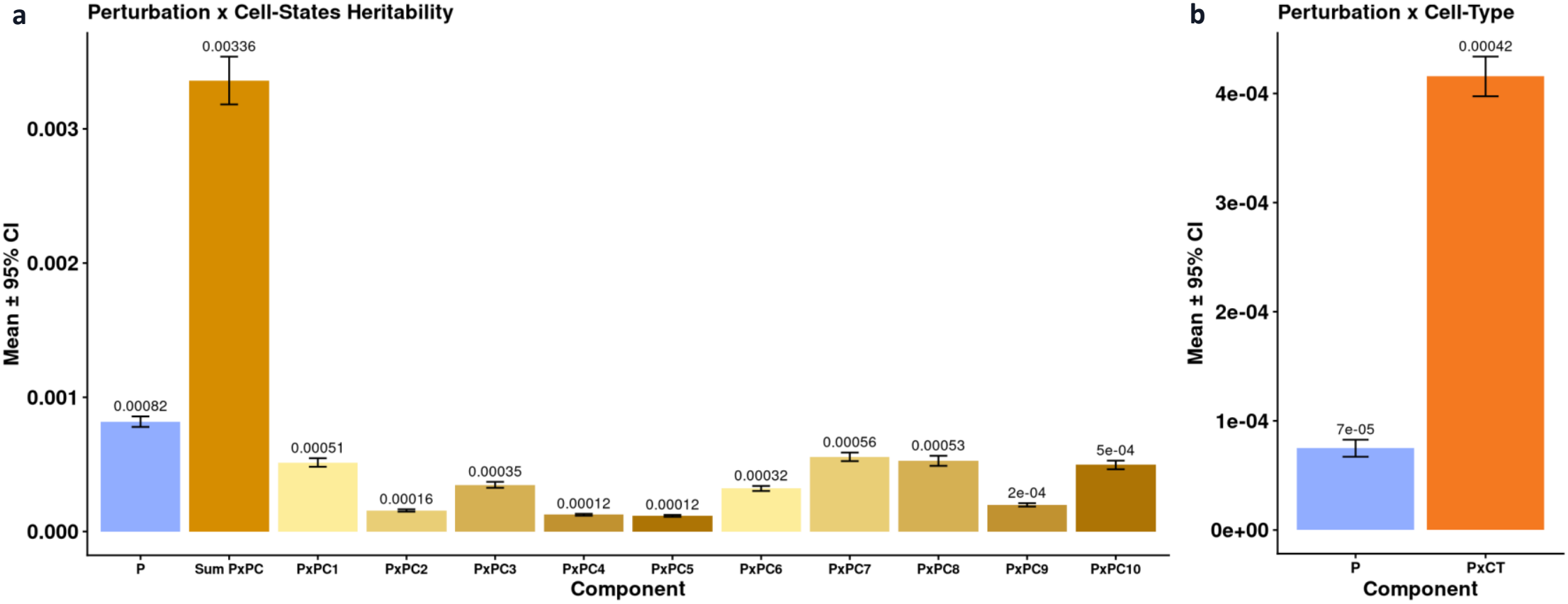
Extension of scHINT to perturbation single-cell data. **a**, Mean perturbation heritability and 95% confidence intervals for the main perturbation effect, the summed perturbation × cell-state interaction component and the individual perturbation × PC components in a genome-wide Perturb-seq screen in K562 cells (2.06 million cells, 7,189 gene perturbations). **b**, Mean perturbation heritability and 95% confidence intervals for the main perturbation effect and the perturbation × cell-type interaction effect in a multiome Perturb-seq dataset (137,604 cells, 19 transcription-factor knockouts, 10 cell types). Perturbation heritability is defined as the proportion of expression variance explained by perturbation effects in perturbed cells.

Next, we applied scHINT to a multi-ome perturb-seq dataset containing 137,604 cells, 19 transcription factor knockouts and 10 cell types^109^. In this multi-cell-type perturbation setting, perturbation-by-cell-type interaction effects explained 4.5-fold more expression variance than main perturbation effects alone (**Fig. 7b**). Together, these analyses show that scHINT can be applied beyond germline genetic regulation to quantify context-dependent transcriptional responses in experimental data. The large perturbation-by-cell-state and perturbation-by-cell-type components indicate that perturbation effects vary substantially across cellular contexts, mirroring our findings in population-scale single-cell QTL data.

## Discussion

A central unresolved question in human genetics is why many GWAS loci often cannot be explained by regulatory effects measured in bulk tissues or by conventional eQTL studies. Our method, scHINT, suggests that a substantial fraction of this missing regulatory signal arises because genetic effects vary across continuous cellular states rather than acting uniformly across cells. Across major brain cell types, the G×Cell-State heritability was often greater than the main genetic heritability, and the two components were only weakly correlated. Thus, genes with little detectable average genetic regulation can still show substantial state-dependent genetic effects.

These context-dependent effects also showed a markedly different genetic architecture from conventional eQTL effects and even from cell-type specific eQTL effects. Main genetic and G×Cell-Type heritability was driven primarily by common variants located near transcription start sites. In contrast, G×Cell-State heritability was disproportionately explained by low-frequency variants and variants farther from promoters. This more closely resembles the architecture of GWAS signals and suggests that cell-state-dependent regulation may help bridge the long-standing gap between disease-associated variants and steady-state eQTLs.

A related question is why GWAS loci frequently implicate evolutionarily constrained genes, whereas standard eQTLs are depleted near such genes. We found that main genetic heritability was negatively associated with gene constraint, while aggregated G×Cell-State heritability was positively associated with constraint. State-dependent regulation may affect functionally important genes without producing large constitutive changes in expression that would be strongly selected against^110^.

To better understand and interpret G×Cell-State heritability, we evaluated its biological and disease relevance using several complementary analyses. G×Cell-State effects showed extensive pathway and disease enrichments that were largely absent for main genetic heritability. An external activation-eQTL dataset further provided orthogonal validation that the inferred cell-state axes capture genuine biological contexts, demonstrating that states derived from observational single-cell data can recover regulatory programs activated by experimental neuronal stimulation. Incorporating G×C effects into the TWAS framework also improved cell-level expression prediction and identified additional disease-risk genes with stronger concordance with rare-variant burden-test results. Together, these complementary lines of evidence support the robustness and functional importance of state-dependent genetic regulation.

Our findings complement recent work by Chen et al.^55^, who developed CIGMA to quantify cell-type-shared and cell-type-specific genetic regulation across discrete blood cell types. Methodologically, CIGMA can be viewed as conceptually similar to our scHINT-G×Cell-Type model, which is applied to sample-level expression, with cell type serving as the interaction context. In contrast, scHINT-G×Cell-State model operates directly on individual cells and models genotype interactions with continuous cell-state variation within a cell type. This distinction is important because substantial regulatory heterogeneity exists among cells assigned to the same cell type and cannot be captured by discrete cell-type labels or sample-level aggregation. Resolving such cell-level context therefore requires methods such as scHINT that can directly model single-cell observations while remaining computationally scalable to millions of cells. Interestingly, both approaches implicate selectively constrained genes, but reveal different patterns of context dependence. Chen et al. found that constrained genes have lower absolute shared and cell-type-specific genetic variance but greater relative eQTL cell-type specificity in blood. In contrast, we find that constrained genes exhibit consistently greater absolute G×Cell-State heritability in brain. In blood, the direction of individual G×PC heritability effects varies across cell-state axes, but the relative contribution of G×Cell-State heritability is consistently greater in constrained genes, in line with the increased relative context specificity reported by Chen et al^55^. These results suggest that the brain may harbor particularly strong state-dependent genetic regulation at constrained genes, while selected blood cell states show similar features. Despite these tissue-specific differences, both analyses support a common pattern in which a greater fraction of genetic regulation at constrained genes emerges in specific cellular contexts. Together, these findings underscore the importance of studying both cell-type-specific and cell-state-specific effects to more fully characterize how genetic regulation varies across biological contexts.

Several limitations and opportunities for future development remain. First, although scHINT can prioritize genes with strong cell-state-dependent effects and reveals that G×Cell-State heritability is disproportionately contributed by low-frequency variants, it does not identify the causal variants underlying these interactions. Resolving such variants will require dedicated G×Cell-State QTL mapping at single-cell resolution. Our results support the feasibility and potential value of this direction, particularly for genes with weak main genetic heritability but substantial G×Cell-State heritability, whose regulatory effects may be obscured in conventional bulk or pseudobulk eQTL analyses.

Second, scHINT models normalized cell-level expression within a Gaussian framework and therefore does not explicitly account for the sparsity of single-cell count data. This sparsity reflects technical factors, including sequencing depth, batch effects and molecular sampling, as well as biological processes such as transcriptional bursting. Poisson or negative-binomial models may better capture count distributions, but their computational cost remains substantial for datasets containing millions of cells. As population-scale single-cell datasets continue to grow, developing scalable cell-level models that retain appropriate count-based assumptions will be increasingly important^111^.

Third, the definitions of both cell type and cell state remain dependent on analytical choices. Our analyses focused on variation within predefined cell types and therefore rely on the accuracy and consistency of cell-type annotation. Marker-based classifications may obscure transitional or difficult-to-classify populations, while annotation strategies often differ across cohorts and resolutions.

Continuous cell-type assignments may provide a more robust alternative, particularly for differentiating or ambiguous populations. Similarly, the cell states analyzed here were inferred from scRNA-seq embeddings. Incorporating directly measured single- or multimodal contexts, including chromatin accessibility, protein abundance, spatial features and epigenomic profiles, could improve the interpretation of interaction effects.

Fourth, to reduce computational burden, G×C-TWAS models were trained using approximately 10,000 metacells and restricted to HapMap3 variants. This approximation likely reduces cellular heterogeneity and may miss signals driven by lower-frequency variants. More computationally efficient prediction models could improve both resolution and power. Extending mediation frameworks such as MESC to explicitly model G×C-regulated expression could further quantify the proportion of disease heritability mediated through cell-state-dependent genetic effects.

Finally, our application of scHINT to perturbation datasets identified substantial perturbation-by-cell-type and perturbation-by-cell-state interaction effects. This context specificity presents an important challenge for the development of virtual cell models: accurate prediction of perturbation responses will require models that capture how genetic, environmental and experimental perturbations interact with both cell identity and cell state.

In summary, scHINT reveals that cell-state-dependent genetic effects represent a substantial and distinct component of gene-expression regulation that is largely missed by bulk and conventional eQTL analyses. By linking these effects to constrained genes, disease-relevant pathways, TWAS associations and perturbation responses, scHINT provides a scalable framework for characterizing how genetic effects vary across continuous cellular contexts.

## Methods

### The scHINT model

For each gene, let *y*_*im*_ denote the normalized expression level measured in cell *m* from individual *i*, where *i* = 1, …, *n* and *m* = 1, …, *M*_*i*_. Let *g*_*i*_ ∈ ℝ^*p*^ denote the vector of standardized cis-genotypes for individual *i*, defined using variants within ±500 kb of the gene, and let *c*_*im*_ ∈ ℝ^*K*^ denote a vector of cell-state features for cell *m*, such as the leading within-cell-type principal components. We model single-cell expression as

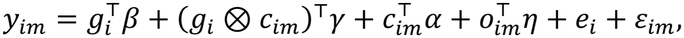

where *β* ∈ ℝ^*p*^ represents the main cis-genetic effect, *γ* ∈ ℝ^*pK*^ represents G×C interaction effects, *α* ∈ ℝ^*K*^ captures non-genetic cell-state effects, and *o*_*im*_ denotes additional covariates. The individual-level random effect *e*_*i*_ captures expression variation shared across cells from the same donor, and *ε*_*im*_ denotes cell-level residual noise. We assume

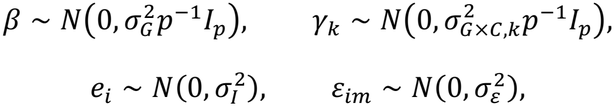

with mutually independent random components. Under this model, *σ*^2^ quantifies the contribution of main cis-genetic effects, whereas *σ*^2^ quantifies the contribution of genetic effects that vary along cell-state axis *k*.

scHINT estimates these components by extending Haseman–Elston regression.

Let

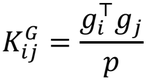

denote the local cis-genetic relatedness between individuals *i* and *j*. For a pair of cells (*i, m*) and (*j, n*), the expected expression product is

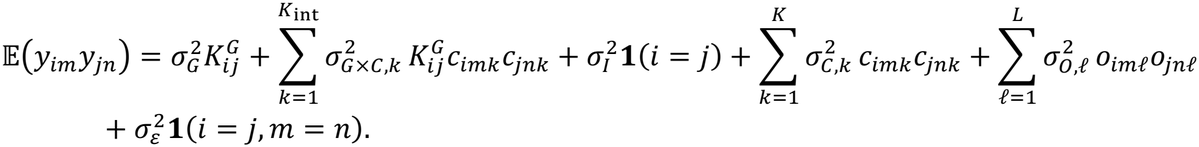

Because the estimator is based on cross-products of distinct cells, the residual term is not used as a fitted HE component for off-diagonal pairs. The key terms are therefore the genetic kernel 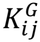, the individual-level kernel ***1***(*i* = *j*), the cell-state kernels *c*_*imk*_*c*_*jnk*_, and the interaction kernels 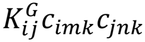. This formulation converts the high-dimensional random-effects model into a low-dimensional regression of expression cross-products on covariance kernels.

In practice, scHINT fits two nested Haseman-Elston regressions. The baseline model is

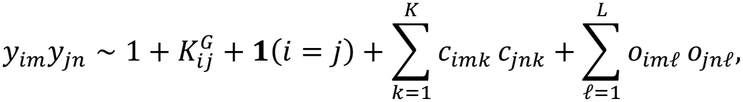

which estimates the main genetic component while adjusting for individual-level variation, cell-state similarity and additional cell-level covariates. The interaction model extends this regression to

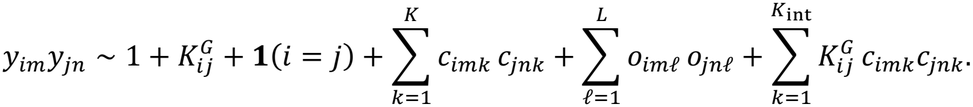

The coefficients of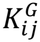 and 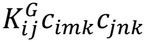estimate the main genetic and genotype-by-cell-state variance components, respectively.

Through the nested Haseman-Elston regressions, the baseline model can estimate the variance components of (*G, I*, PC_1_, …, PC_*K*_, *O*_1_, …, *O*_*L*_), and interaction model adds 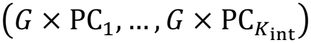. The aggregate context-dependent component is then defined as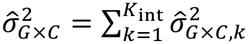.

### Scalable estimation

A direct implementation of HE regression regresses the cross-product *y*_*im*_*y*_*jn*_ of every pair of distinct cells on the kernels of the baseline model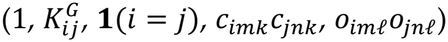 and of the interaction model, which adds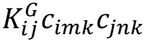. With *M* cells in total this requires *M*(*M* − 1)*/*2 pair observations, so building the regression takes approximately *O*(*M*^2^*K*^2^) time and *O*(*M*^2^) memory, where *K* is the number of cell-state PCs and covariates. This is computationally infeasible for datasets containing millions of cells.

scHINT avoids forming cell pairs by exploiting the structure of the kernels. Every kernel is the product of an individual-level term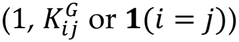 and a cell-level term (*c*_*imk*_*c*_*jnk*_ or *o*_*imP*_*o*_*jnP*_), so sums over cell pairs factor into products of per-individual sums. For each gene we therefore compute, a set of sufficient statistics: the number of cells, the sums of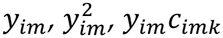 and *y*_*im*_*o*_*imP*_ (*X*^T^*y*), and the cross-product matrices of the cell-state features and covariates ∑_*m*_ *c*_*imk*_ *c*_*imk*>_, ∑_*m*_ *c*_*imk*_ *o*_*imP*_ and ∑_*m*_ *o*_*imP*_ *o*_*imP*>_ (*X*^T^*X*). The entries of *X*^T^*X* and *X*^T^*y* are then assembled by looping over pairs of individuals, weighting each pair by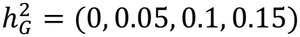 and its powers, and the variance components are obtained by solving the ordinary least-squares normal equations,

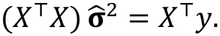

Computing the donor-level statistics requires a single pass over the cells, and assembling *X*^T^*X* and *X*^T^*y* reduces the dominant computational cost to approximately *O*(*N*^2^*K*^2^) time and *O*(*N*^2^) memory (for the individual-level GRM), where *N* is the number of individuals (*N* ≪ *M*). Because the statistics are additive across donors, standard errors are obtained from a donor-level jackknife that removes one individual’s contributions from the sums, without refitting from cell-level data, so uncertainty is estimated at little additional cost.

### Simulating cell-level gene expression data

We performed simulations to evaluate scHINT under controlled single-cell expression architectures with known contributions from main cis-genetic effects, genotype-by-cell-state effects and non-genetic sources of cell-level variation. Simulations were based on real genotype data to preserve local allele-frequency and linkage-disequilibrium structure. For each simulated gene, we selected cis-variants located within 500 kb of the gene body and standardized the resulting genotype matrix. The same individual-level genotype vector was then assigned to all cells from the corresponding donor.

For each gene, expression was simulated for 500 individuals with *M* cells per individual. Simulation settings were defined by

- the number of cells per individual, *M* ∈ (25, 50, 75, 100),
- the number of causal variants, *n*_causal_ = 4,
- the main genetic variance parameter, 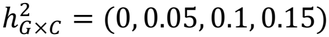,
- the genotype-by-cell-state variance parameter, 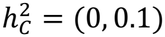,
- the cell state variance parameter, 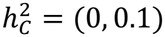,
- the variance parameter from other covariates (sex, pseudo-bulk PCs, etc.), 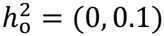,
- the cell-level count mean *μ*_*c*_ = (10, 25)

For individual *i* and cell *m*, let *g*_*i*_ ∈ ℝ^*p*^ denote the standardized cis-genotype vector, and let *c*_*im*_ denote a standardized cell-state variable. For each gene, *n*_causal_ variants were sampled as causal, with main genetic effects and genotype-by-cell-state effects generated independently. Effect sizes for the main genetic and G×C components were drawn from normal distributions with mean zero and variance equal to the corresponding heritability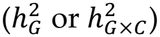 divided by n_causal, using the same set of causal variants; non-causal SNPs were assigned zero effect for both components. The cell-state variable was drawn from a standard normal distribution. Other sample-level covariates were represented by pseudo-bulk expression principal components only.

The latent expression mean was generated as

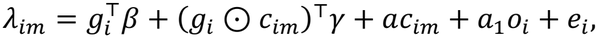

where 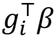 denotes the main cis-genetic effect, (*g*_*i*_ *⊙ c*_*im*_)^T^*γ* denotes the genotype-by-cell-state interaction effect, *ac*_*im*_ is a non-genetic cell-state effect, *a*_1_*o*_*i*_ is the effect of other covariates and *e*_*i*_ is an individual-level random effect shared across all cells from donor *i*. Specifically, we do not include cell-level noise in the heritability and normalize the heritability estimate by

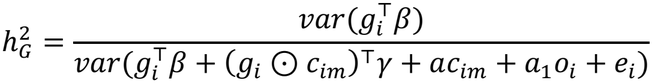

The sum of effect-size vectors is scaled to have variance 1 and each component has the variance corresponding to the variance parameter, 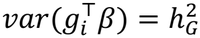, etc.

Observed cell-level expression was generated from a Poisson count model,

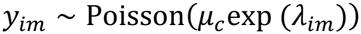

We then used normalized log(1 + counts) values as the input for scHINT. For the pseudo-bulk analysis, gene expression was generated by aggregating cell-level expression within each individual, followed by log-normalization.

For each simulation setting, we evaluated all methods across 500 independent replications. For each method and estimand, we summarized empirical performance using the mean heritability estimate and its standard error. We then constructed 95% confidence intervals for the mean estimate and compared these intervals with the corresponding ground-truth heritability values specified in the simulation.

### Harmonization and processing of brain snRNA-seq datasets

To maximize statistical power, we integrated two postmortem human brain single-nucleus RNA-seq datasets from Fujita et al.^40^ and Mathys et al.^63^. Both datasets were generated from ROS/MAP cohorts and included partially overlapping donors. Single-nucleus RNA-seq quality control and preprocessing were performed following the standard pipeline described in Fujita et al.

After harmonizing and combining the single-cell expression profiles across the two datasets, 584 samples with both snRNA-seq and whole-genome sequencing (WGS) data were retained for analysis. All downstream analyses were then conducted separately within six major brain cell types: excitatory neurons (Exc, 1648653 cells), inhibitory neurons (Inh, 570964 cells), astrocytes (Ast, 361103 cells), oligodendrocytes (Oli, 965066 cells), oligodendrocyte precursor cells (OPC, 148826 cells), and microglia (Mic, 160849 cells). We then normalized the single-cell read counts using a transcript-per-10,000 transformation. For comparison, we also generated pseudobulk expression profiles for each cell type by aggregating unique molecular identifier (UMI) counts across all nuclei assigned to a given cell type within each individual. All heritability analyses adjusted for sample-level covariates, including sex, age, study cohort, 5 genotype principal components and 25 pseudobulk expression PCs. Cell-level analyses additionally adjusted for cell-level covariates, including log-transformed UMI counts, batch, mitochondrial percentage, and five cell-level expression PCs. As in the simulation analyses, we benchmarked scHINT against three alternative approaches, HEreg^64^ which was applied to cell-type-specific pseudo-bulk expression, scGeneHE^61^ and scHINT-base, which were applied directly to cell-level expression.

### Processing of OneK1K scRNAseq data

We analyzed the OneK1K scRNA-seq, which includes 1,248,980 cells from 981 donors and provides raw UMI counts for cells that passed the original quality control, together with Azimuth level-2 annotations. We grouped cells into 15 fine cell types: CD4 Naive, TCM, TEM, and CTL; CD8 Naive, TCM, and TEM; B naive, intermediate, and memory; NK, NK CD56bright, and NK proliferating; and CD14 and CD16 monocytes. These cell types contained 1,660–272,553 cells from 661–928 donors. We retained genes detected in at least 1% of cells within each cell type, resulting in 9,161–10,065 genes.

We normalized UMI counts as log1p(TP10K) using each cell’s total count across the retained genes. Within each cell type, we regressed normalized expression for each gene on sex, age, percentage of mitochondrial reads, sequencing pool, genotype PCs 1–5, and log library size using ordinary least squares. We then computed cell state principal components from the residual expression of highly expressed genes within each cell type.

### Cell-State estimation

To define continuous cell-state axes for scHINT, we evaluated multiple approaches, including within-cell-type principal component analysis (PCA^67^), across-cell-type PCA, residual PCA^69^, and consensus non-negative matrix factorization (cNMF^70^). For PCA-based cell-state construction, we used the residualized meta-cell expression profiles as input.

For within-cell-type PCA, this procedure was performed separately within each major brain cell type, allowing cell-state axes to capture transcriptional heterogeneity specific to each cell type. For across-cell-type PCA, PCA was performed using meta-cells from all major cell types together, and the resulting PCs were then used as shared cell-state axes across cell types. We also applied the residPCA framework, a recently proposed approach for identifying disease-associated or context-dependent cellular states from single-cell RNA-seq data. In parallel, we applied cNMF to identify non-negative gene expression programs and used the resulting program usage scores as alternative cell-state representations.

We compared these different cell-state definitions in downstream scHINT analyses and used them to evaluate the robustness of genotype-by-cell-state heritability estimates across alternative representations of cellular heterogeneity.

### Correlation with gene constraint scores

We tested whether main and interaction heritability estimates were associated with the continuous gene scores (**Supplementary Table 5**). For each gene score, associations were estimated using linear regression with the gene score as the outcome and heritability estimates as predictors. We used Spearman-scale analyses, in which both gene scores and heritability estimates were first rank-transformed and then standardized before regression. The reported effect size corresponds to the standardized regression coefficient, and p values were obtained from the corresponding regression model. For interaction heritability components, we used joint models to estimate the association of context-dependent heritability after adjusting for the corresponding main genetic heritability.

- One-cell-type main genetic heritability: score ∼ 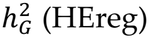 (HEreg).
- Two-cell-type interaction heritability: score ∼ 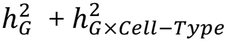 (scHINT). Adjust for the main heritability of two cell types.
- >All-cell-type interaction heritability: score ∼ 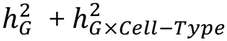 (scHINT). Adjust for the main heritability of all cell types.
- >Cell-state model main genetic heritability: score ∼ 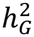 (scHINT-baseline)
- G x PC_k_ heritability: score ∼ 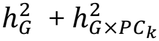 (scHINT). Adjust for the main heritability.
- Sum of G x cell-state heritability: score ∼ 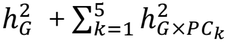 (scHINT). Adjust for the main heritability.
- Proportion of heritability: score 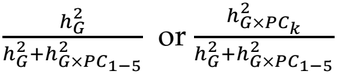.

### Partitioning heritability by minor allele frequency and distance to the TSS

To partition heritability by variant class, we extended scHINT to include multiple genetic relationship matrices (GRMs), each calculated from a subset of the cis-SNPs, and calculated a corresponding G×PC term for each GRM. For the minor allele frequency (MAF) partition, SNPs were grouped into low-frequency (1–5%), intermediate (5–20%) and high-frequency (>20%) classes. For the partition by distance to the transcription start site (TSS), SNPs were grouped into those inside and outside a window around the TSS, using a ±1-kb promoter window and a ±10-kb promoter-proxy window.

The proportion of G (or G×Cell-State) heritability attributable to each class was calculated as the variance component of that class divided by the sum across classes. The same partition was applied to the G×Cell-Type model.

### Gene set enrichment analysis

We obtained gene sets from MSigDB v2024.1 human-symbol GMT files, including Hallmark, Reactome, KEGG legacy, Gene Ontology biological process, cellular component, molecular function, and cell-type signature collections. We retained gene sets containing 10–500 genes to avoid unstable estimates from very small sets and nonspecific signals from overly broad annotations.

For each tested gene set, we coded each gene with a binary membership indicator. We evaluated enrichment using logistic regression, with gene-set membership as the outcome and rank-normalized heritability estimates as predictors. To reduce confounding by gene expression level, we included mean expression level as a covariate.

- One-cell-type main genetic heritability: gene-set membership ∼ 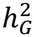 (HEreg) + mean expression.
- Two-cell-type interaction heritability: gene-set membership ∼ 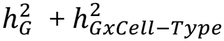 (scHINT) + mean expression.
- All-cell-type interaction heritability: gene-set membership ∼ 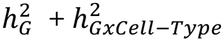 (scHINT) + mean expression.
- Cell-state model main genetic heritability: gene-set membership ∼ *h*^2^ (scHINT-baseline) + mean expression
- G x PC_k_ heritability: gene-set membership ∼ 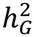 (scHINT) + mean expression.

Adjust for the main heritability.

### Bi-clustering of Gene Ontology enrichment

To identify shared biological patterns across G×PC components, we assembled the Gene Ontology (GO) enrichment results into a matrix with GO terms as rows and cell-type-specific G×PC components as columns, with entries given by enrichment significance (−log10 P). Rows and columns were bi-clustered by hierarchical clustering using Euclidean distance and visualized with the pheatmap package^112^. Columns were annotated with the correlation of each heritability component with pLI and with disease heritability enrichment from S-LDSC.

### Stratified LDSC analysis of disease heritability enrichment

To evaluate whether genes with high expression heritability were enriched for disease-associated genetic risk, we performed stratified linkage disequilibrium score regression (S-LDSC) using GWAS summary statistics from seven brain-related diseases and traits. For each heritability model, genes were ranked by their estimated heritability, and the top 300 genes were selected to define a gene set. We performed this procedure separately for each heritability component, including main genetic heritability and interaction heritability from the one-cell-type, two-cell-type, all-cell-type, and cell-state scHINT models.

For each selected gene set, we constructed a binary genomic annotation by including variants located within a 100-kb window around the transcription start site of each gene. These annotations were then tested using S-LDSC to estimate whether SNP heritability for each brain-related disease was enriched near genes with high expression heritability. Disease heritability enrichment was quantified as the proportion of SNP heritability explained by the annotation divided by the proportion of SNPs included in the annotation. Enrichment p values were obtained from the S-LDSC regression coefficient for each annotation.

### Meta-cell analysis

To reduce sparsity and computational burden in estimating heritability using scGeneHE and building TWAS gene expression prediction model, we constructed donor-specific meta-cells. Meta-cell construction was performed separately for each dataset and each major brain cell type.

Within each donor and cell type, raw counts were first normalized to transcripts per 10,000 reads before microclustering. Microclustering was performed using the “applyMicroClustering” function from the VISION R package^96^, which groups transcriptionally similar cells prior to aggregation. The target number of cells per microcluster was determined adaptively based on the total number of cells, with the goal of retaining approximately 50,000 meta-cells per cell type for scGeneHE and 10,000 meta-cells for TWAS. Donors with too few cells were assigned to a single fallback meta-cell containing all available cells from that donor and cell type.

After microclustering, gene expression was aggregated on the raw-count scale. For meta-cell (k), the count for gene (g) was defined as the sum of raw UMI counts across all cells assigned to that microcluster,

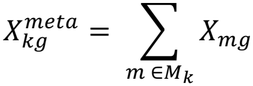

where (*M*_*k*_) denotes the set of cells in meta-cell (k). Thus, the resulting meta-cell expression matrix represents summed UMI counts and remains suitable for downstream count-based normalization and modeling.

We also summarized covariates at the meta-cell level. Library size was computed by summing total UMI counts across cells within each microcluster. Mitochondrial percentage was summarized as the total cell-level mitochondrial fraction across all cells in the microcluster. Donor-level variables, including study, sex, age and genetic principal components, were assigned uniformly to all meta-cells derived from the same donor.

### G×Cell-State TWAS model

For each gene and cell type, we trained cis-genetic prediction models using metacell-level logTP10K expression residuals and donor-matched genotypes. To reduce the computational burden, we also used microclustering to obtain approximately 10,000 meta-cells per cell type. For a target gene *g*, we selected SNPs within ±500 kb of the gene body. Before fitting genetic prediction models, we adjusted expression for the cell-state PCs and other covariates.

We compared a base cis-expression model with a two-step G×Cell-State model. All weights were estimated using elastic-net regression with *α* = 0.5. The penalty parameter was selected by 5-fold cross-validation within the training set. SNPs with nonzero elastic-net coefficients were retained as prediction weights. Let *G*_*ij*_ denote the standardized dosage of SNP *j* for donor *i*, and let *y*_*icg*_ denote the normalized expression of gene *g* in metacell *c* from donor *i*.

The base model used donor genotypes to predict the cell-level expression phenotype directly:

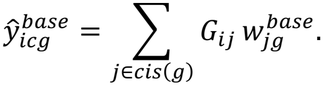

This model captures the main cis-genetic component of expression but does not allow the genetic effect to vary across cell states.

For the G×Cell-State model, we used a two-step decomposition. First, we estimated a donor-level main genetic model from the donor mean of the cell-level phenotype:

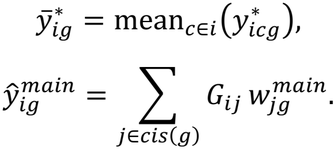

Second, we removed the donor-level mean phenotype from each metacell to define a within-donor residual phenotype:

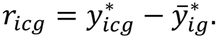

We then modeled this within-donor residual using genotype-by-cell-state interaction features. For each SNP *j* and each of the first five cell-state PCs, we constructed

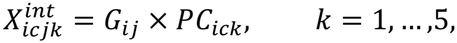

and fit an elastic-net model

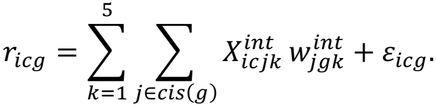

The final G×cell-state prediction score combined the donor-level main genetic component and the cell-state-dependent interaction component:

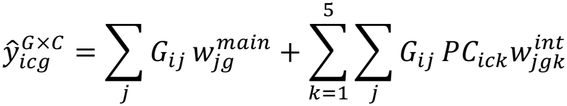

### G×Cell-State TWAS cross-validation and prediction evaluation

We evaluated prediction performance using 5-fold donor-level cross-validation. Donors, rather than cells, were assigned to folds, ensuring that all cells from the same donor appeared either in the training set or in the test set. For each fold, we trained the base model, the donor-level main model, and the G×Cell-State interaction model using the training donors only. We then predicted expression in held-out donors and tested whether the predicted expression scores explained observed cell-level expression.

Because multiple cells came from the same donor, we evaluated held-out prediction using a linear mixed model with a donor random intercept. For the base model, we fit

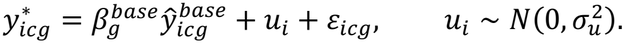

To evaluate the contribution of the main and PC-specific interaction components jointly, we fit

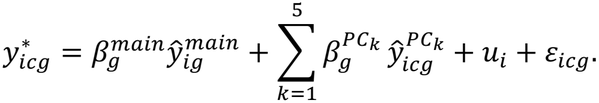

The fixed-effect coefficient and P value for each prediction score were extracted from the mixed model in each cross-validation fold.

For each gene, cell type, and prediction component, we combined evidence across the five held-out folds. The cross-validation effect estimate was summarized as the mean fixed-effect estimate across folds:

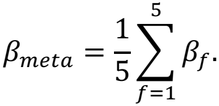

P values were combined using a signed Stouffer Z-score. For fold *f*, we converted the two-sided P value *p*_*f*_ and effect estimate *β*_*f*_ to a signed Z-score:

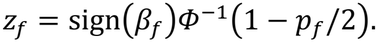

The fold-level statistics were then combined as

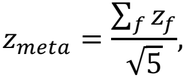

with two-sided P value

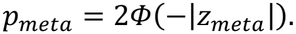

We defined the significant prediction components if *β*_*meta*_ > 0 and *p*_*meta*_ *<* 0.05. After cross-validation, we refit the base, main, and G×Cell-State interaction models using all donors.

The final weights were then used in the second-stage TWAS analysis with FUSION^9^ to estimate summary-statistics-based gene–trait associations.

### G×Cell-State eQTL analysis

To test for cell-state-dependent effects of individual variants, we fitted for each SNP–gene pair in a given cell type a linear mixed-effects model of cell-level expression with donor as a random effect. For cell c from donor i,

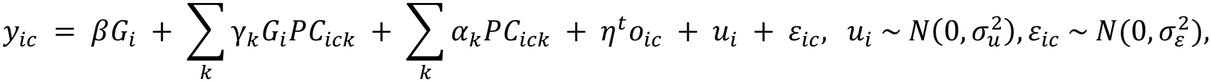

where *G*_*i*_ is the dosage of the SNP, *PC*_*ick*_ is the score of cell-state PC k, *o*_*ic*_ denotes the sample- and cell-level covariates used in the heritability analyses, *u*_*i*_is the donor random intercept and *ε*_*ic*_ is residual noise. The main eQTL effect (*β*) and each G×PC interaction effect (*γ*_*k*_) were tested. For the stratified analysis, microglia were divided into tertiles of PC5, and the genotype effect on expression was estimated within each tertile using the same model without interaction terms, again with donor as a random effect.

### Perturbation heritability

We estimated G×Cell-State heritability in perturb-seq datasets by treating perturbations as the genotype/G term. For each gene, let *y*_*m*_ denote the normalized expression level in cell *m*, let *p*_*m*_ denote its assigned perturbation, and let *c*_*m*_ be a vector of K standardized cell-state principal components. Expression is centered and scaled to unit variance. We model

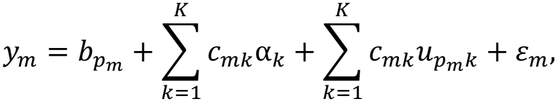

where 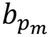 is the expression shift shared by cells receiving perturbation *p*_*m*_, α_*k*_ capture the cell-state k effects, 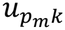 is the perturbation-specific change in expression along PC k, and *ε*_*m*_, is cell-level residual variation. The random effects are mutually independent across perturbations and PC axes, with

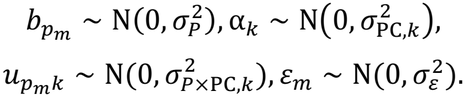

Under this model, 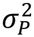 quantifies perturbation-associated variation that is shared across cell states, 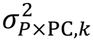 quantifies variation along cell-state k, whereas 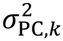 quantifies variation in the perturbation response along PC axis k.

We also extended Haseman–Elston regression to estimate the variance components. For two cells m and n *m* ≠ *n*,

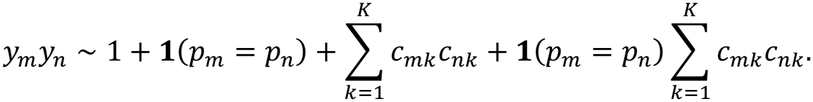

The coefficients of these three terms represent perturbation variance components, cell-state variance components and interaction variance components.

To scale estimation across genes, we use the fact that genes measured in the same cells share perturbation assignments and cell-state PCs. The resulting pairwise predictor cross-products are therefore calculated once and reused; only the expression-dependent cross-products change by gene. We process cells in blocks and accumulate compact, perturbation-level summaries of expression and PC-weighted expression. Combining these summaries recovers the required pairwise quantities without constructing or storing cell-pair observations, reducing repeated computation and peak memory use.

## Acknowledgment

The authors thank members of the Gusev Lab at Dana-Farber Cancer Institute and Harvard Medical School for fruitful discussions regarding this manuscript.

## Data availability

Our scHINT G×Cell-State/Type and perturbation heritability estimation are available at https://zenodo.org/records/24254462. MSigDB gene sets are available at https://www.gsea-msigdb.org/gsea/msigdb. OneK1K individual level data is available at https://onek1k.org/. ROS/MAP single-nucleus RNA-seq data from Mathys et al is be available here: https://www.synapse.org/#!Synapse:syn52293417. ROS/MAP single-nucleus RNA-seq from Fujita et al. is available here: https://www.synapse.org/#!Synapse:syn31512863.

## Code availability

scHINT software is available at https://lidawangpsu.github.io/scHINT/.

Our analysis codes are available at https://github.com/LidaWangPSU/scHINT.

LDSC: https://github.com/bulik/ldsc.

## Reference

1. Maurano, M.T. et al. Systematic localization of common disease-associated variation in regulatory DNA. Science 337, 1190–5 (2012).

2. Finucane, H.K. et al. Partitioning heritability by functional annotation using genome-wide association summary statistics. Nat Genet 47, 1228–35 (2015).

3. Gusev, A. et al. Partitioning heritability of regulatory and cell-type-specific variants across 11 common diseases. Am J Hum Genet 95, 535–52 (2014).

4. Roadmap Epigenomics, C. et al. Integrative analysis of 111 reference human epigenomes. Nature 518, 317–30 (2015).

5. Giambartolomei, C. et al. Bayesian test for colocalisation between pairs of genetic association studies using summary statistics. PLoS Genet 10, e1004383 (2014).

6. Hormozdiari, F. et al. Colocalization of GWAS and eQTL Signals Detects Target Genes. Am J Hum Genet 99, 1245–1260 (2016).

7. Wallace, C. A more accurate method for colocalisation analysis allowing for multiple causal variants. PLoS Genet 17, e1009440 (2021).

8. Gamazon, E.R. et al. A gene-based association method for mapping traits using reference transcriptome data. Nat Genet 47, 1091–8 (2015).

9. Gusev, A. et al. Integrative approaches for large-scale transcriptome-wide association studies. Nat Genet 48, 245–52 (2016).

10. Hu, Y. et al. A statistical framework for cross-tissue transcriptome-wide association analysis. Nat Genet 51, 568–576 (2019).

11. Khunsriraksakul, C. et al. Integrating 3D genomic and epigenomic data to enhance target gene discovery and drug repurposing in transcriptome-wide association studies. Nat Commun 13, 3258 (2022).

12. Mancuso, N. et al. Probabilistic fine-mapping of transcriptome-wide association studies. Nat Genet 51, 675–682 (2019).

13. Zhou, D. et al. A unified framework for joint-tissue transcriptome-wide association and Mendelian randomization analysis. Nat Genet 52, 1239–1246 (2020).

14. Bowden, J., Davey Smith, G. & Burgess, S. Mendelian randomization with invalid instruments: effect estimation and bias detection through Egger regression. Int J Epidemiol 44, 512–25 (2015).

15. Bowden, J., Davey Smith, G., Haycock, P.C. & Burgess, S. Consistent Estimation in Mendelian Randomization with Some Invalid Instruments Using a Weighted Median Estimator. Genet Epidemiol 40, 304–14 (2016).

16. Zhu, Z. et al. Integration of summary data from GWAS and eQTL studies predicts complex trait gene targets. Nat Genet 48, 481–7 (2016).

17. Carithers, L.J. & Moore, H.M. The Genotype-Tissue Expression (GTEx) Project. Biopreserv Biobank 13, 307–8 (2015).

18. Mostafavi, H., Spence, J.P., Naqvi, S. & Pritchard, J.K. Systematic differences in discovery of genetic effects on gene expression and complex traits. Nat Genet 55, 1866–1875 (2023).

19. Zhang, J. & Zhao, H. eQTL studies: from bulk tissues to single cells. J Genet Genomics 50, 925–933 (2023).

20. Montgomery, S.B. Regulatory genomics at biobank scales. Nat Rev Genet 26, 657–658 (2025).

21. See, Y.X., Stuart, T. & Shin, J.W. Cis-regulatory elements at cellular resolution. Nat Rev Genet 26, 653–654 (2025).

22. Yazar, S. et al. Single-cell eQTL mapping identifies cell type-specific genetic control of autoimmune disease. Science 376, eabf3041 (2022).

23. Mu, Z. et al. The impact of cell type and context-dependent regulatory variants on human immune traits. Genome Biol 22, 122 (2021).

24. Zhernakova, D.V. et al. Identification of context-dependent expression quantitative trait loci in whole blood. Nat Genet 49, 139–145 (2017).

25. Calderon, D. et al. Landscape of stimulation-responsive chromatin across diverse human immune cells. Nat Genet 51, 1494–1505 (2019).

26. Fairfax, B.P. et al. Innate immune activity conditions the effect of regulatory variants upon monocyte gene expression. Science 343, 1246949 (2014).

27. Liang, L. et al. Single-cell multiomics of neuron activation reveals context-specific genetics of brain disorders. Science 392, eadw3949 (2026).

28. Ota, M. et al. Dynamic landscape of immune cell-specific gene regulation in immune-mediated diseases. Cell 184, 3006–3021 e17 (2021).

29. Soskic, B. et al. Immune disease risk variants regulate gene expression dynamics during CD4(+) T cell activation. Nat Genet 54, 817–826 (2022).

30. Strober, B.J. et al. Dynamic genetic regulation of gene expression during cellular differentiation. Science 364, 1287–1290 (2019).

31. Umans, B.D., Battle, A. & Gilad, Y. Where Are the Disease-Associated eQTLs? Trends Genet 37, 109–124 (2021).

32. Walker, R.L. et al. Genetic Control of Expression and Splicing in Developing Human Brain Informs Disease Mechanisms. Cell 181, 745 (2020).

33. Marshall, L., Raychaudhuri, S. & Viatte, S. Understanding rheumatic disease through continuous cell state analysis. Nat Rev Rheumatol 21, 323–335 (2025).

34. Lek, M. et al. Analysis of protein-coding genetic variation in 60,706 humans. Nature 536, 285–91 (2016).

35. Siewert-Rocks, K.M., Kim, S.S., Yao, D.W., Shi, H. & Price, A.L. Leveraging gene co-regulation to identify gene sets enriched for disease heritability. Am J Hum Genet 109, 393–404 (2022).

36. Nathan, A. et al. Single-cell eQTL models reveal dynamic T cell state dependence of disease loci. Nature 606, 120–128 (2022).

37. Macosko, E.Z. et al. Highly Parallel Genome-wide Expression Profiling of Individual Cells Using Nanoliter Droplets. Cell 161, 1202–1214 (2015).

38. Tang, F. et al. mRNA-Seq whole-transcriptome analysis of a single cell. Nat Methods 6, 377–82 (2009).

39. Bryois, J. et al. Cell-type-specific cis-eQTLs in eight human brain cell types identify novel risk genes for psychiatric and neurological disorders. Nat Neurosci 25, 1104–1112 (2022).

40. Fujita, M. et al. Cell subtype-specific effects of genetic variation in the Alzheimer’s disease brain. Nat Genet 56, 605–614 (2024).

41. Jang, B. et al. A meta-analysis of single-nucleus expression quantitative trait loci linking genetic risk to brain disorders. Nat Genet 58, 737–747 (2026).

42. Lopes, K.P. et al. Genetic analysis of the human microglial transcriptome across brain regions, aging and disease pathologies. Nat Genet 54, 4–17 (2022).

43. van der Wijst, M.G.P. et al. Single-cell RNA sequencing identifies celltype-specific cis-eQTLs and co-expression QTLs. Nat Genet 50, 493–497 (2018).

44. Wang, L. et al. An atlas of single-cell eQTLs dissects autoimmune disease genes and identifies novel drug classes for treatment. Cell Genom 5, 100820 (2025).

45. Zhang, Z.E., Kim, A., Suboc, N., Mancuso, N. & Gazal, S. Efficient count-based models improve power and robustness for large-scale single-cell eQTL mapping. Nat Genet (2026).

46. Chen, M. & Dahl, A. A robust model for cell type-specific interindividual variation in single-cell RNA sequencing data. Nat Commun 15, 5229 (2024).

47. Krockenberger, L. et al. FastGxC: Fast and powerful context-specific eQTL mapping in bulk and single-cell data. Cell Genom 6, 101250 (2026).

48. Lin, Z. & Su, C. scTWAS: a powerful statistical framework for single-cell transcriptome-wide association studies. Nat Commun 17(2026).

49. Ray, A. et al. Single-cell transcriptome-wide Mendelian randomization and colocalization analyses uncover cell-specific mechanisms in atherosclerotic cardiovascular disease. Am J Hum Genet 112, 1597–1609 (2025).

50. Venkatesh, S. et al. Single-nucleus transcriptome-wide association study of human brain disorders. Nature 657, 1016–1026 (2026).

51. Wang, L. et al. Integrating axis quantitative trait loci looks beyond cell types and offers insights into brain-related traits. Nat Commun 16, 10606 (2025).

52. Wang, L. et al. Integrating single cell expression quantitative trait loci summary statistics to understand complex trait risk genes. Nat Commun 15, 4260 (2024).

53. Zhang, M.J. et al. Polygenic enrichment distinguishes disease associations of individual cells in single-cell RNA-seq data. Nat Genet 54, 1572–1580 (2022).

54. Wagner, A., Regev, A. & Yosef, N. Revealing the vectors of cellular identity with single-cell genomics. Nat Biotechnol 34, 1145–1160 (2016).

55. Chen, M. et al. Cell-type-specific eQTLs underlie the genetic architecture of complex traits. Nature 659, 451–458 (2026).

56. de Klein, N. et al. Brain expression quantitative trait locus and network analyses reveal downstream effects and putative drivers for brain-related diseases. Nat Genet 55, 377–388 (2023).

57. Vosa, U. et al. Large-scale cis- and trans-eQTL analyses identify thousands of genetic loci and polygenic scores that regulate blood gene expression. Nat Genet 53, 1300–1310 (2021).

58. Huang, Q.Q., Ritchie, S.C., Brozynska, M. & Inouye, M. Power, false discovery rate and Winner’s Curse in eQTL studies. Nucleic Acids Res 46, e133 (2018).

59. Finucane, H.K. et al. Heritability enrichment of specifically expressed genes identifies disease-relevant tissues and cell types. Nat Genet 50, 621–629 (2018).

60. Yao, D.W., O’Connor, L.J., Price, A.L. & Gusev, A. Quantifying genetic effects on disease mediated by assayed gene expression levels. Nat Genet 52, 626–633 (2020).

61. Xu, Z. et al. Estimating the cis -heritability of gene expression using single cell expression profiles controls false positive rate of eGene detection. bioRxiv (2025).

62. Zhou, W. et al. Efficient and accurate mixed model association tool for single-cell eQTL analysis. medRxiv (2024).

63. Mathys, H. et al. Single-cell atlas reveals correlates of high cognitive function, dementia, and resilience to Alzheimer’s disease pathology. Cell 186, 4365–4385 e27 (2023).

64. Haseman, J.K. & Elston, R.C. The investigation of linkage between a quantitative trait and a marker locus. Behav Genet 2, 3–19 (1972).

65. Pazokitoroudi, A. et al. Efficient variance components analysis across millions of genomes. Nat Commun 11, 4020 (2020).

66. Wu, Y. & Sankararaman, S. A scalable estimator of SNP heritability for biobank-scale data. Bioinformatics 34, i187–i194 (2018).

67. Jolliffe, I.T. Principal component analysis, xxix, 487 p. (Springer, New York, 2002).

68. Jolliffe, I.T. & Cadima, J. Principal component analysis: a review and recent developments. Philos Trans A Math Phys Eng Sci 374, 20150202 (2016).

69. Carver, S., Taraszka, K., Groha, S. & Gusev, A. Discovery of disease-associated cellular states using ResidPCA in single-cell RNA and ATAC sequencing data. HGG Adv 7, 100538 (2026).

70. Kotliar, D. et al. Identifying gene expression programs of cell-type identity and cellular activity with single-cell RNA-Seq. Elife 8(2019).

71. Pazokitoroudi, A. et al. A scalable and robust variance components method reveals insights into the architecture of gene-environment interactions underlying complex traits. Am J Hum Genet 111, 1462–1480 (2024).

72. Liu, W. et al. A statistical framework to identify cell types whose genetically regulated proportions are associated with complex diseases. PLoS Genet 19, e1010825 (2023).

73. Rumker, L. et al. Identifying genetic variants that inluence the abundance of cell states in single-cell data. Nat Genet 56, 2068–2077 (2024).

74. Evans, C., Hardin, J. & Stoebel, D.M. Selecting between-sample RNA-Seq normalization methods from the perspective of their assumptions. Brief Bioinform 19, 776–792 (2018).

75. Ko, B.S., Lee, S.B. & Kim, T.K. A brief guide to analyzing expression quantitative trait loci. Mol Cells 47, 100139 (2024).

76. Bennett, D.A. et al. Religious Orders Study and Rush Memory and Aging Project. J Alzheimers Dis 64, S161–S189 (2018).

77. Perez-Gonzalez, A.P. et al. The ROSMAP project: aging and neurodegenerative diseases through omic sciences. Front Neuroinform 18, 1443865 (2024).

78. Chen, M. et al. Cell-type-specific eQTLs underlie the genetic architecture of complex traits. Nature (2026).

79. Glassberg, E.C., Gao, Z., Harpak, A., Lan, X. & Pritchard, J.K. Evidence for Weak Selective Constraint on Human Gene Expression. Genetics 211, 757–772 (2019).

80. Taylor, D.J. et al. Sources of gene expression variation in a globally diverse human cohort. Nature 632, 122–130 (2024).

81. Brown, C.D., Mangravite, L.M. & Engelhardt, B.E. Integrative modeling of eQTLs and cis-regulatory elements suggests mechanisms underlying cell type specificity of eQTLs. PLoS Genet 9, e1003649 (2013).

82. Dimas, A.S. et al. Common regulatory variation impacts gene expression in a cell type-dependent manner. Science 325, 1246–50 (2009).

83. Demontis, D. et al. Genome-wide analyses of ADHD identify 27 risk loci, refine the genetic architecture and implicate several cognitive domains. Nat Genet 55, 198–208 (2023).

84. Mullins, N. et al. Genome-wide association study of more than 40,000 bipolar disorder cases provides new insights into the underlying biology. Nat Genet 53, 817–829 (2021).

85. Howard, D.M. et al. Genome-wide meta-analysis of depression identifies 102 independent variants and highlights the importance of the prefrontal brain regions. Nat Neurosci 22, 343–352 (2019).

86. Trubetskoy, V. et al. Mapping genomic loci implicates genes and synaptic biology in schizophrenia. Nature 604, 502–508 (2022).

87. van Rheenen, W. et al. Common and rare variant association analyses in amyotrophic lateral sclerosis identify 15 risk loci with distinct genetic architectures and neuron-specific biology. Nat Genet 53, 1636–1648 (2021).

88. Eadb et al. Consensus meta-analysis of genome-wide association studies for Alzheimer’s disease and related dementias. Nat Genet 58, 1214–1225 (2026).

89. Nalls, M.A. et al. Identification of novel risk loci, causal insights, and heritable risk for Parkinson’s disease: a meta-analysis of genome-wide association studies. Lancet Neurol 18, 1091–1102 (2019).

90. Fromer, M. et al. De novo mutations in schizophrenia implicate synaptic networks. Nature 506, 179–84 (2014).

91. Purcell, S.M. et al. A polygenic burden of rare disruptive mutations in schizophrenia. Nature 506, 185–90 (2014).

92. Kosoy, R. et al. Genetics of the human microglia regulome refines Alzheimer’s disease risk loci. Nat Genet 54, 1145–1154 (2022).

93. Brown, G.C., St George-Hyslop, P., Paolicelli, R.C. & Lemke, G. Microglial phagocytosis in Alzheimer disease. Nat Rev Neurol 22, 54–69 (2026).

94. Xu, Z. et al. Microglia-specific regulation of lipid metabolism in Alzheimer’s disease revealed by microglial depletion in 5xFAD Mice. Nat Commun 16, 9156 (2025).

95. Bano, D., Ehninger, D. & Bagetta, G. Decoding metabolic signatures in Alzheimer’s disease: a mitochondrial perspective. Cell Death Discov 9, 432 (2023).

96. DeTomaso, D. et al. Functional interpretation of single cell similarity maps. Nat Commun 10, 4376 (2019).

97. Dudbridge, F. Power and predictive accuracy of polygenic risk scores. PLoS Genet 9, e1003348 (2013).

98. Weeks, E.M. et al. Leveraging polygenic enrichments of gene features to predict genes underlying complex traits and diseases. Nat Genet 55, 1267–1276 (2023).

99. Gerlach, P.A., Milind, N., Spence, J.P. & Pritchard, J.K. High false sign rates in transcriptome-wide association studies. bioRxiv, 2025.12.19.695550 (2025).

100. Hou, K., Pazokitoroudi, A., Strober, B., Jiang, X. & Price, A.L. Functionally informed cis and trans proteome-wide association studies prioritize disease-critical genes. Res Sq (2026).

101. Wainberg, M. et al. Opportunities and challenges for transcriptome-wide association studies. Nat Genet 51, 592–599 (2019).

102. Abduljaleel, Z. et al. Evidence of trem2 variant associated with triple risk of Alzheimer’s disease. PLoS One 9, e92648 (2014).

103. Jonsson, T. et al. Variant of TREM2 associated with the risk of Alzheimer’s disease. N Engl J Med 368, 107–16 (2013).

104. Li, R., Wang, X. & He, P. The most prevalent rare coding variants of TREM2 conferring risk of Alzheimer’s disease: A systematic review and meta-analysis. Exp Ther Med 21, 347 (2021).

105. Wang, G., Sarkar, A., Carbonetto, P. & Stephens, M. A simple new approach to variable selection in regression, with application to genetic fine mapping. J R Stat Soc Series B Stat Methodol 82, 1273–1300 (2020).

106. Das, S. et al. Next-generation genotype imputation service and methods. Nat Genet 48, 1284–1287 (2016).

107. Taliun, D. et al. Sequencing of 53,831 diverse genomes from the NHLBI TOPMed Program. Nature 590, 290–299 (2021).

108. Replogle, J.M. et al. Mapping information-rich genotype-phenotype landscapes with genome-scale Perturb-seq. Cell 185, 2559–2575 e28 (2022).

109. Martin-Rufino, J.D. et al. Transcription factor networks disproportionately enrich for heritability of blood cell phenotypes. bioRxiv (2024).

110. Spence, J.P. et al. Specificity, length and luck drive gene rankings in association studies. Nature 649, 918–925 (2026).

111. Pullin, J.M., Choi, J., McCarthy, D.J. & Wallace, C. Flexible and efficient count-distribution and mixed-model methods for eQTL mapping with quasar. medRxiv, 2025.07.17.25331702 (2025).

112. Hu, K. Become Competent in Generating RNA-Seq Heat Maps in One Day for Novices Without Prior R Experience. Methods Mol Biol 2239, 269–303 (2021).

