## Supplementary figures and notes for "Single-Cell Gene Expression Heritability Informed by Genotype × Cell-Context Interactions"

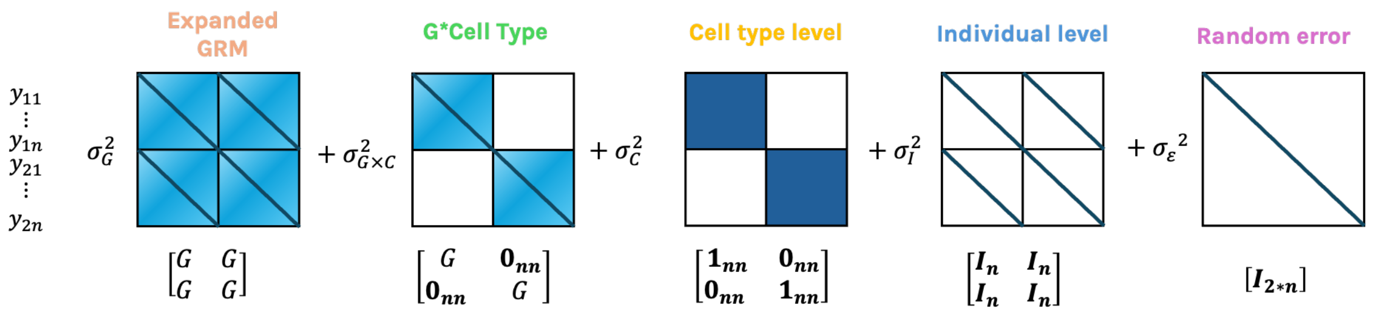


**Supplementary Fig. 1 | Variance-component structure of the G×Cell-Type model.** Covariance kernels used by scHINT-G×Cell-Type to model pseudo-bulk expression in two cell types. The 2 × 2 block layout separates pairs of observations from the same cell type (diagonal blocks) from pairs in different cell types (off-diagonal blocks). From left to right: the genetic relatedness matrix (GRM) expanded to all observations (main genetic variance shared across cell types, $\sigma_G^2$); the interaction GRM (G×Cell-Type variance, $\sigma_{G\times CT}^2$); a block-diagonal indicator of the same cell type (cell-type-level variance, $\sigma_{CT}^2$); an indicator of the same individual (individual-level variance, $\sigma_I^2$); and the identity matrix (residual variance, $\sigma_\varepsilon^2$).

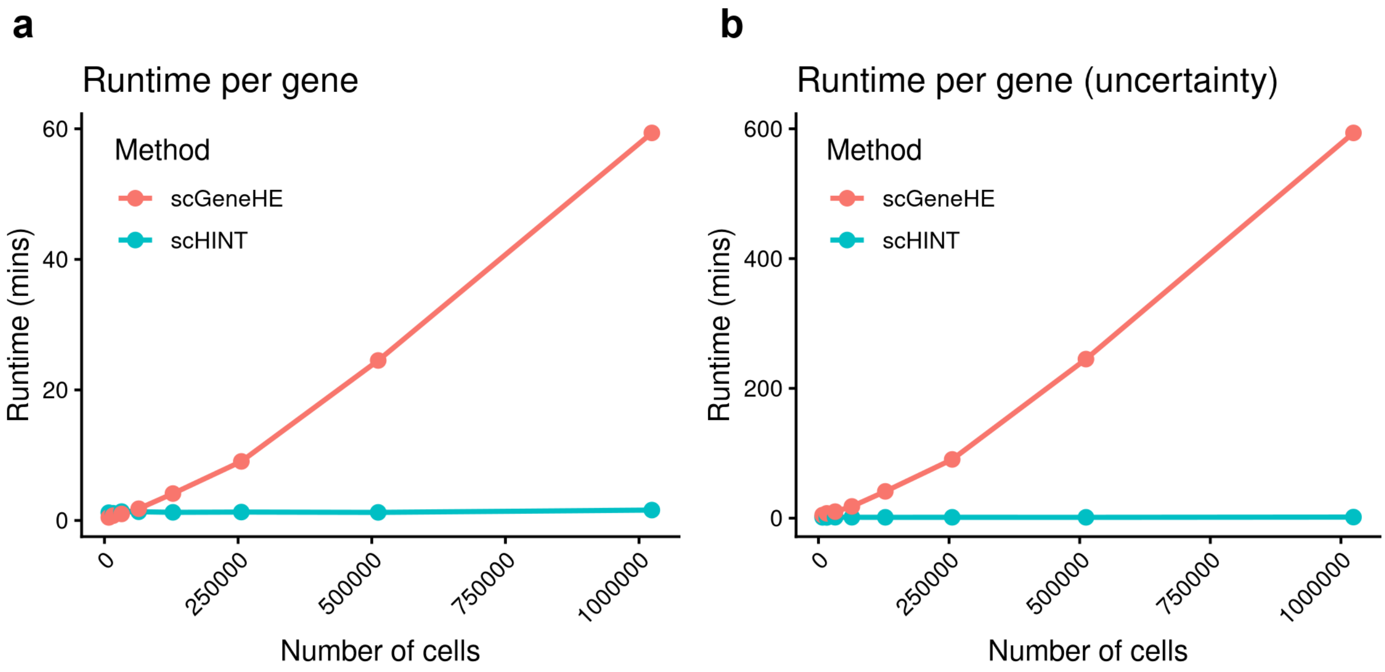


**Supplementary Fig. 2 | Computational efficiency of scHINT compared with scGeneHE. a,** Runtime per gene (minutes) as a function of the number of cells (up to 1 million) for scGeneHE (red) and scHINT (teal), for heritability estimation. b, Runtime when uncertainty is also estimated (donor-level jackknife for scHINT; 10-round bootstrap for scGeneHE). scGeneHE runtime grows with the number of cells, whereas scHINT runtime is nearly constant because it operates on donor-level sufficient statistics. At 1 million cells scHINT is about 36-fold faster for estimation and about 370-fold faster when uncertainty is included.

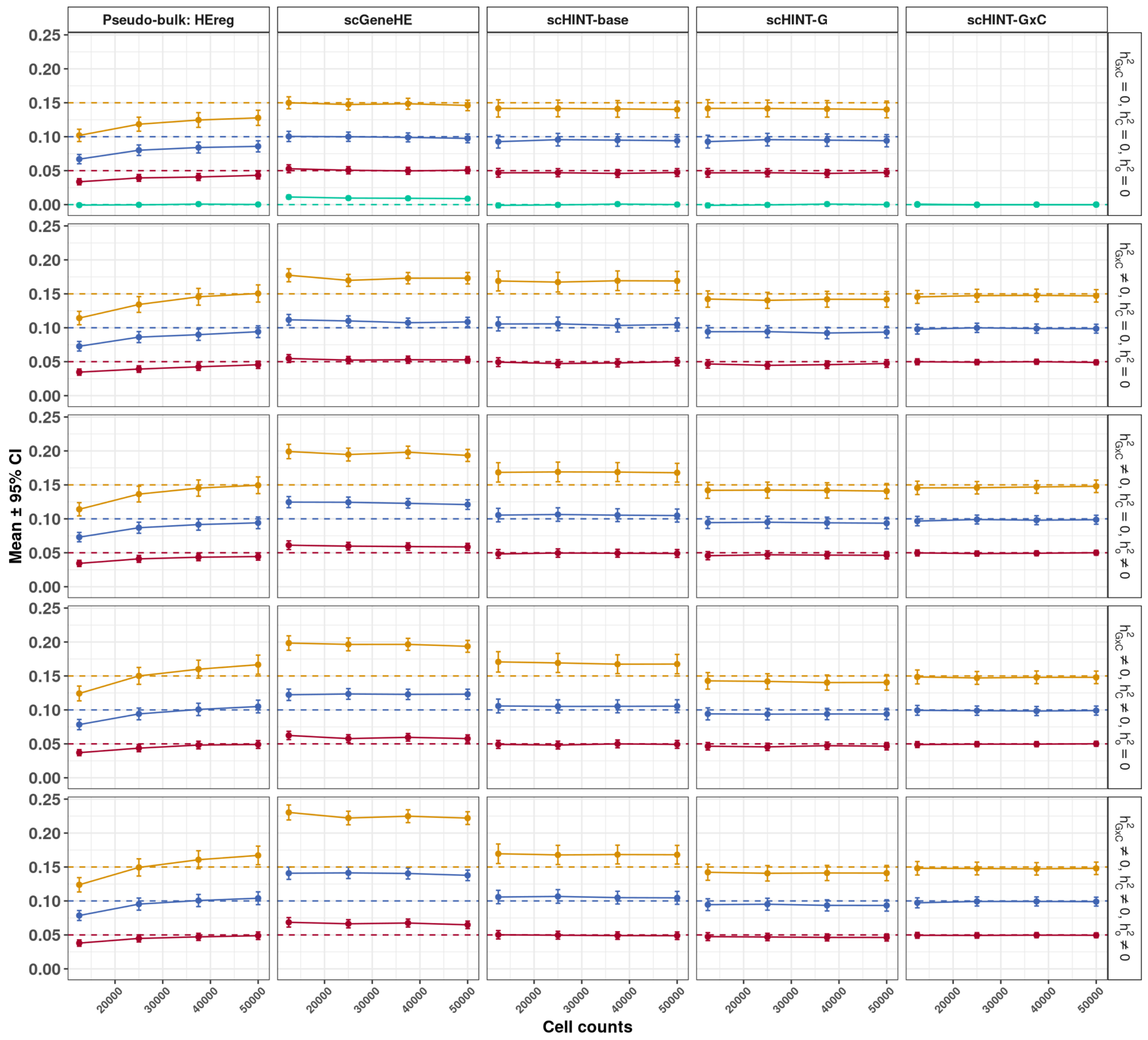


**Supplementary Fig. 3 | Simulation benchmark of heritability estimators at low read depth ($\mu_c = 10$).** Mean estimates and 95% confidence intervals (CI) across 500 simulation replicates for HE regression applied to pseudo-bulk data (Pseudo-bulk: HEreg) and for scGeneHE, scHINT-base (no interaction term), scHINT-G and scHINT-G×C applied to cell-level data. Each simulated dataset comprised 500 individuals with 25–100 cells per individual (12,500–50,000 cells in total; x axis) and a mean read count per cell of $\mu_c = 10$. Columns show methods; the y axis shows the estimated main genetic heritability ($h^2_G$) in every column except scHINT-G×C, which shows the estimated G×Cell-State heritability ($h^2_{G\times C}$). Rows show five variance scenarios: $h^2_{G\times C} = 0$ with cell-state variance ($h^2_C$) and other-covariate variance ($h^2_o$) both zero (null for G×C); $h^2_{G\times C} \neq 0$ with $h^2_C = h^2_o = 0$; $h^2_{G\times C} \neq 0$ with $h^2_o \neq 0$; $h^2_{G\times C} \neq 0$ with $h^2_C \neq 0$; and $h^2_{G\times C} \neq 0$ with both $h^2_C \neq 0$ and $h^2_o \neq 0$. Colours denote the simulated heritability level (gold, 0.15; blue, 0.10; red, 0.05; green, 0) and dashed lines of the same colour mark the true values; where $h^2_{G\times C} \neq 0$, $h^2_G$ and $h^2_{G\times C}$ were set equal. Heritability is the normalized estimate that excludes cell-level residual variance from the denominator (Methods). HEreg on pseudo-bulk data is biased downwards when few cells are available per individual; scGeneHE and scHINT-base are inflated when $h^2_{G\times C} \neq 0$; scGeneHE is also more inflated when $h^2_o \neq 0$; scHINT-G and scHINT-G×C are unbiased across scenarios.

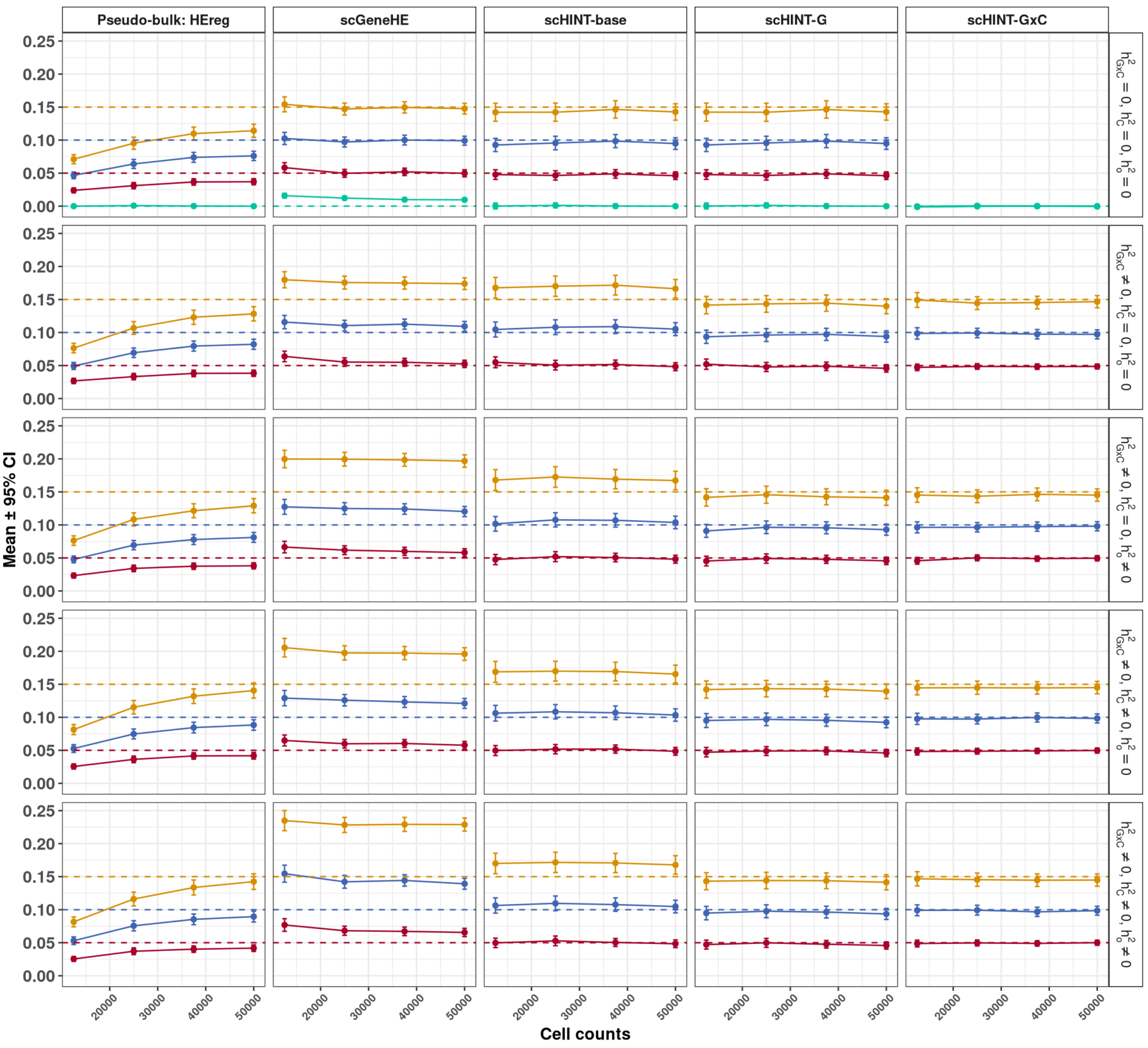


**Supplementary Fig. 4 | Simulation benchmark of heritability estimators at high read depth ($\mu_c = 25$).** Mean estimates and 95% confidence intervals (CI) across 500 simulation replicates for HE regression applied to pseudo-bulk data (Pseudo-bulk: HEreg) and for scGeneHE, scHINT-base (no interaction term), scHINT-G and scHINT-G×C applied to cell-level data. Each simulated dataset comprised 500 individuals with 25–100 cells per individual (12,500–50,000 cells in total; x axis) and a mean read count per cell of $\mu_c = 25$. Columns show methods; the y axis shows the estimated main genetic heritability ($h^2_G$) in every column except scHINT-G×C, which shows the estimated G×Cell-State heritability ($h^2_{G\times C}$). Rows show five variance scenarios: $h^2_{G\times C} = 0$ with cell-state variance ($h^2_C$) and other-covariate variance ($h^2_o$) both zero (null for G×C); $h^2_{G\times C} \neq 0$ with $h^2_C = h^2_o = 0$; $h^2_{G\times C} \neq 0$ with $h^2_o \neq 0$; $h^2_{G\times C} \neq 0$ with $h^2_C \neq 0$; and $h^2_{G\times C} \neq 0$ with both $h^2_C \neq 0$ and $h^2_o \neq 0$. Colours denote the simulated heritability level (gold, 0.15; blue, 0.10; red, 0.05; green, 0) and dashed lines of the same colour mark the true values; where $h^2_{G\times C} \neq 0$, $h^2_G$ and $h^2_{G\times C}$ were set equal. Heritability is the normalized estimate that excludes cell-level residual variance from the denominator (Methods). Compared with Supplementary Fig. 3, the higher read count increases cell-level variation that pseudo-bulk analysis cannot model, so the downward bias of pseudo-bulk HEreg is more pronounced; the other methods behave as in Supplementary Fig. 3.

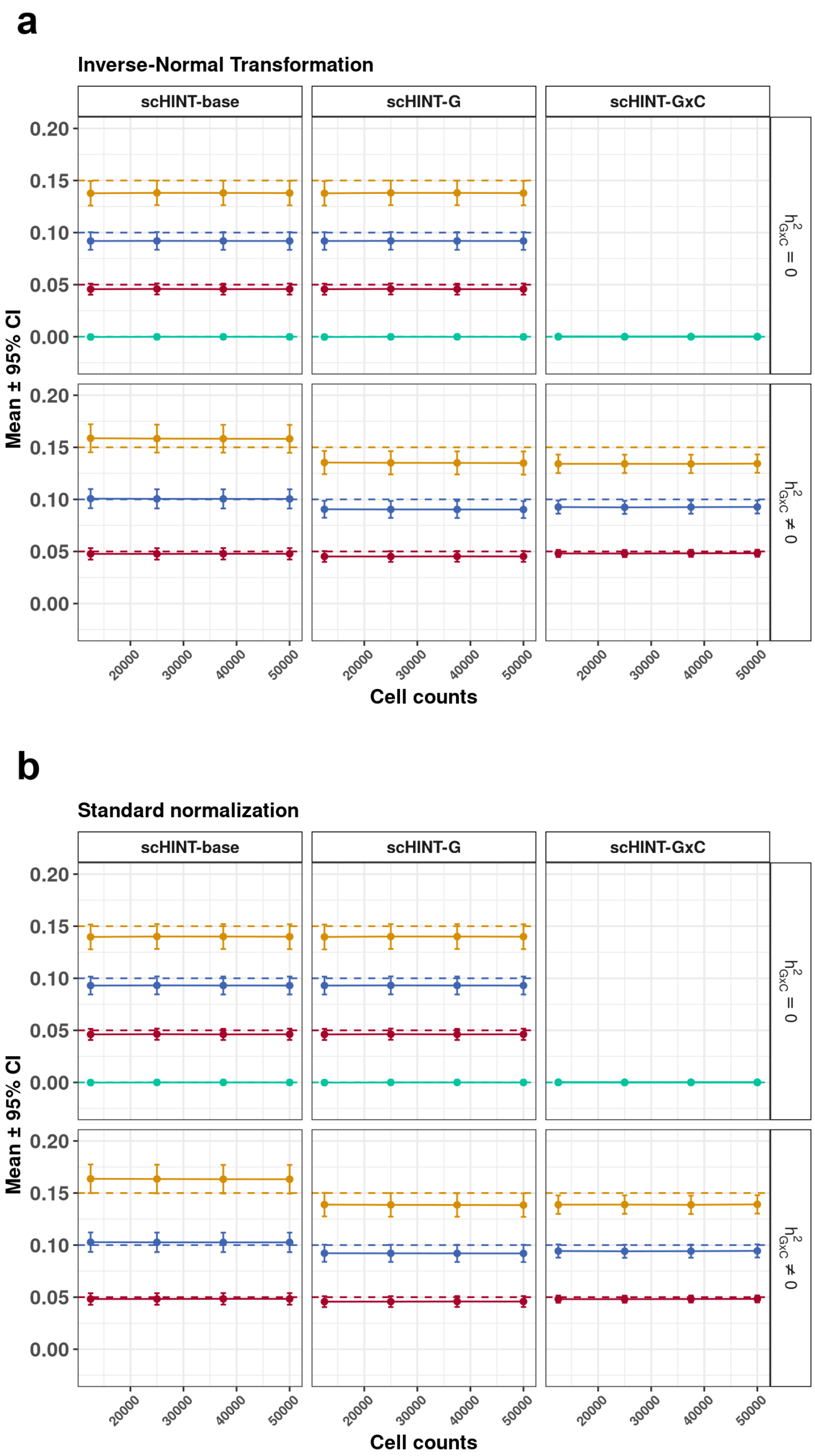


**Supplementary Fig. 5 | Effect of expression standardization on scHINT estimates.** Simulations comparing inverse-normal transformation (**a**) with standard normalization (**b**) of cell-level expression before fitting scHINT. In each panel, columns show scHINT-base, scHINT-G and scHINT-G×C estimates, rows show simulations without ($h^2_{G\times C} = 0$) or with ($h^2_{G\times C} \neq 0$) G×Cell-State effects, and the x axis shows the total number of cells. Points and error bars show mean ± 95% CI across simulation replicates; colours and dashed lines indicate simulated true values as in Supplementary Fig. 3 (gold, 0.15; blue, 0.10; red, 0.05; green, 0). All estimators are centred on zero under the null. Standard normalization gives estimates closer to the simulated values; inverse-normal transformation introduces a modest downward bias, particularly for the G×C component. scHINT-base overestimates $h^2_G$ when G×C effects are present.

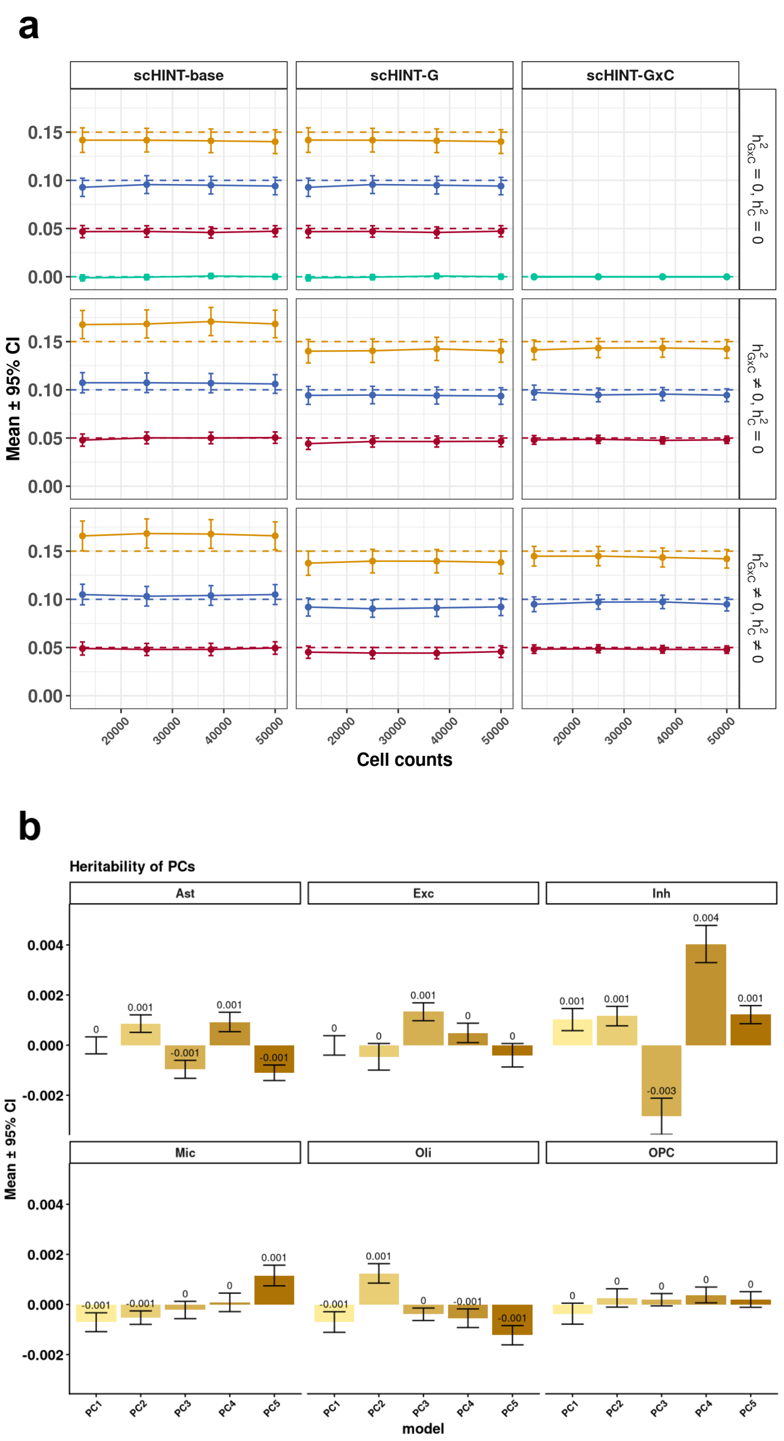


**Supplementary Fig. 6 | scHINT is robust to heritable cell-state axes. a**, Simulations in which the cell-states were themselves partly genetically determined (10% of PC variance explained by genotype). Columns show scHINT-base, scHINT-G and scHINT-G×C estimates as a function of the total number of cells; rows show $h^2_{G\times C} = 0$ with $h^2_C = 0$, $h^2_{G\times C} \neq 0$ with $h^2_C = 0$, and $h^2_{G\times C} \neq 0$ with $h^2_C \neq 0$ ($h^2_C$, variance of the marginal cell-state effect). Colors and dashed lines indicate simulated true values (gold, 0.15; blue, 0.10; red, 0.05; green, 0). Mean ± 95% CI across replicates. **b**, Estimated heritability of the top five within-cell-type cell-state PCs (PC1–PC5) in ROS/MAP snRNA-seq data for each major brain cell type. Bars show mean estimates (printed above bars) with 95% CI. Estimates are close to zero (largest below 0.5%), so heritability of the PCs is unlikely to bias G or G×C estimates.

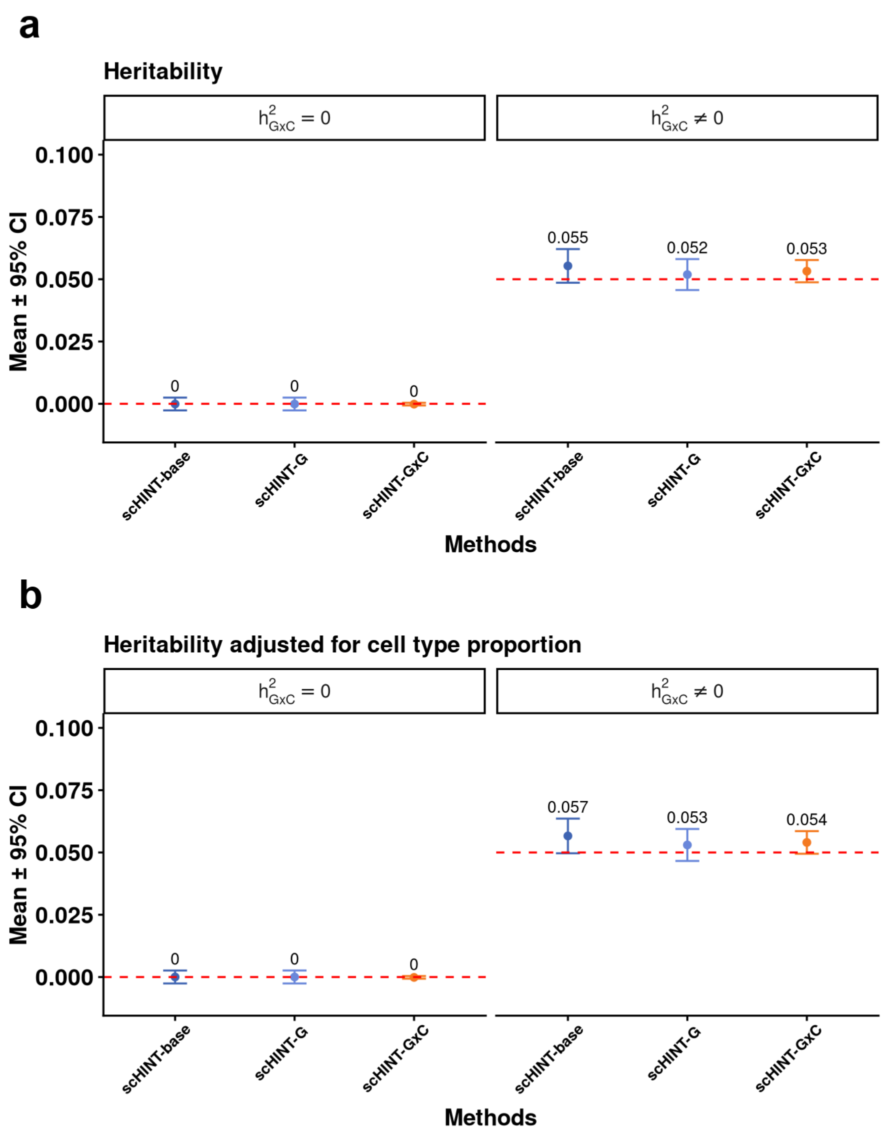


**Supplementary Fig. 7 | scHINT is robust to genetically driven variation in cell number.** Simulations in which the number of cells (cell type proportions) contributed by each individual depended on genotype. **a**, Heritability estimated without adjustment. **b**, Heritability estimated after including cell-type abundance (cells per individual) as a covariate. In both panels, points show mean ± 95% CI of the heritability estimated by scHINT-base, scHINT-G and scHINT-G×C when $h^2_{G\times C} = 0$ (left) or $h^2_{G\times C} \neq 0$ (right); numbers are mean estimates. Dashed red lines mark the simulated values (0 and 0.05). Estimates are close to the simulated values with or without the covariate, indicating that genetically driven differences in cell number do not bias within-cell-type estimation.

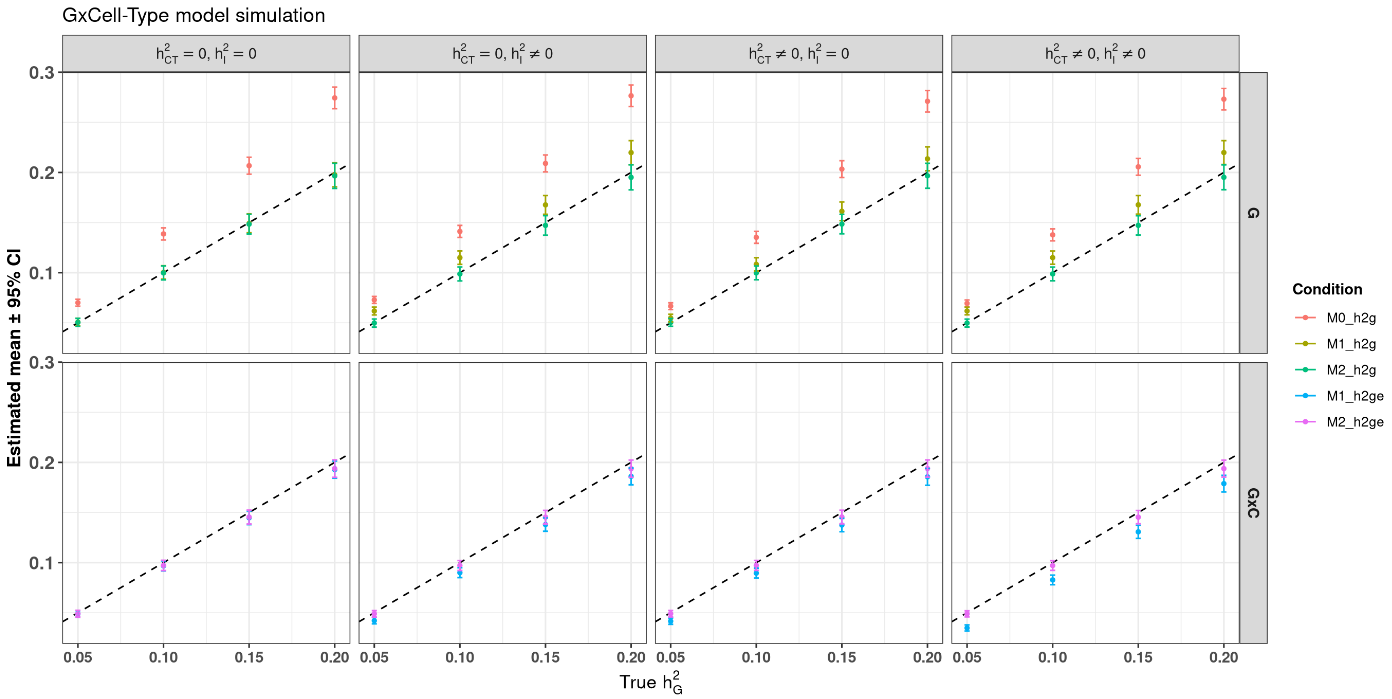


**Supplementary Fig. 8 | Simulation of the G×Cell-Type model in two cell types.** Estimated (y axis) versus true (x axis) heritability in simulated pseudo-bulk expression of two cell types, with $h^2_G = h^2_{G\times CT}$ varied from 0.05 to 0.20. Top row, estimated main genetic heritability (G); bottom row, estimated G×Cell-Type heritability (G×C). Columns show scenarios in which the cell-type-level variance ($h^2_{CT}$) and the individual-level variance ($h^2_I$) are absent (0) or present (≠ 0; simulated value 0.1). Colours denote the fitted model: Model 0 (M0), main genetic component only; Model 1 (M1), main genetic and G×Cell-Type components; Model 2 (M2), additionally including marginal cell-type and individual-level variance components (the full scHINT model). Suffixes h2g and h2ge denote main genetic and G×Cell-Type heritability estimates, respectively. Points show mean ± 95% CI across replicates; dashed line, identity. M0 overestimates $h^2_G$ when G×Cell-Type effects are present, M1 is biased when either additional component is present, and M2 is unbiased in all scenarios.

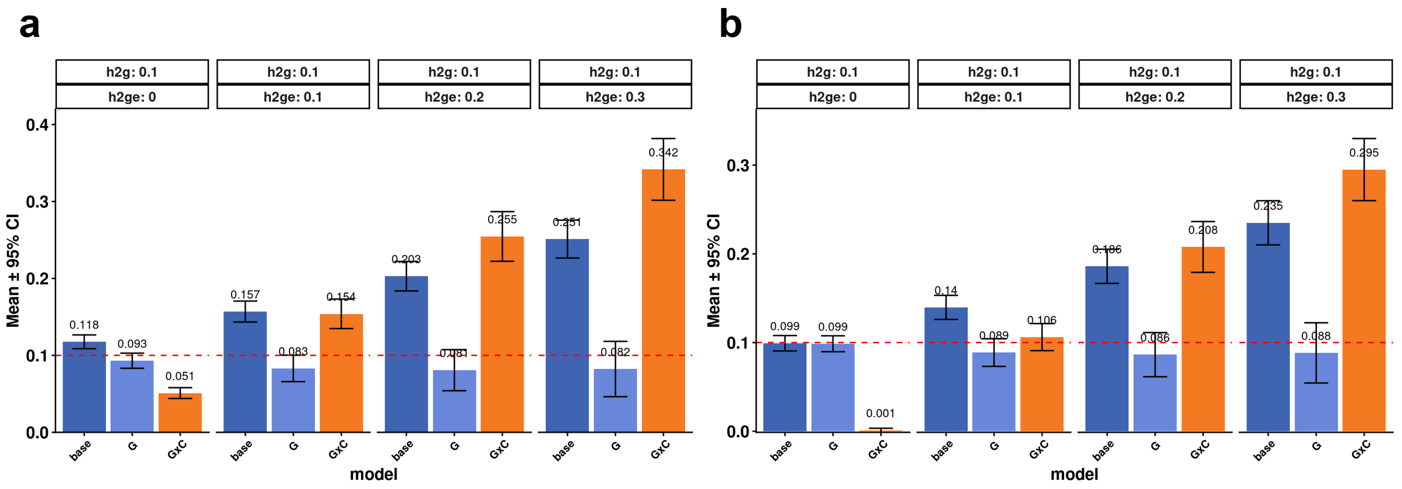


**Supplementary Fig. 9 | Simulation of genetically driven cell-type abundance in the G×Cell-Type model.** Simulations of two cell types in which genotype increases the abundance of one cell type relative to the other. Bars show the mean ± 95% CI of the heritability estimated by the baseline model (base; main genetic component only), and by the main genetic (G) and G×Cell-Type (G×C) components of the full model, for $h^2_G = 0.1$ (h2g) and $h^2_{G\times CT} = 0$, 0.1, 0.2 or 0.3 (h2ge) (panel headers); numbers are mean estimates. **a**, Without adjustment for cell-type proportion. **b**, With cell-type proportion included as a covariate. The dashed red line marks the simulated $h^2_G$. Without adjustment, composition effects are partly absorbed into the G×Cell-Type component (underestimating G and overestimating G×C); adjustment for cell-type proportion brings both estimates close to their simulated values.

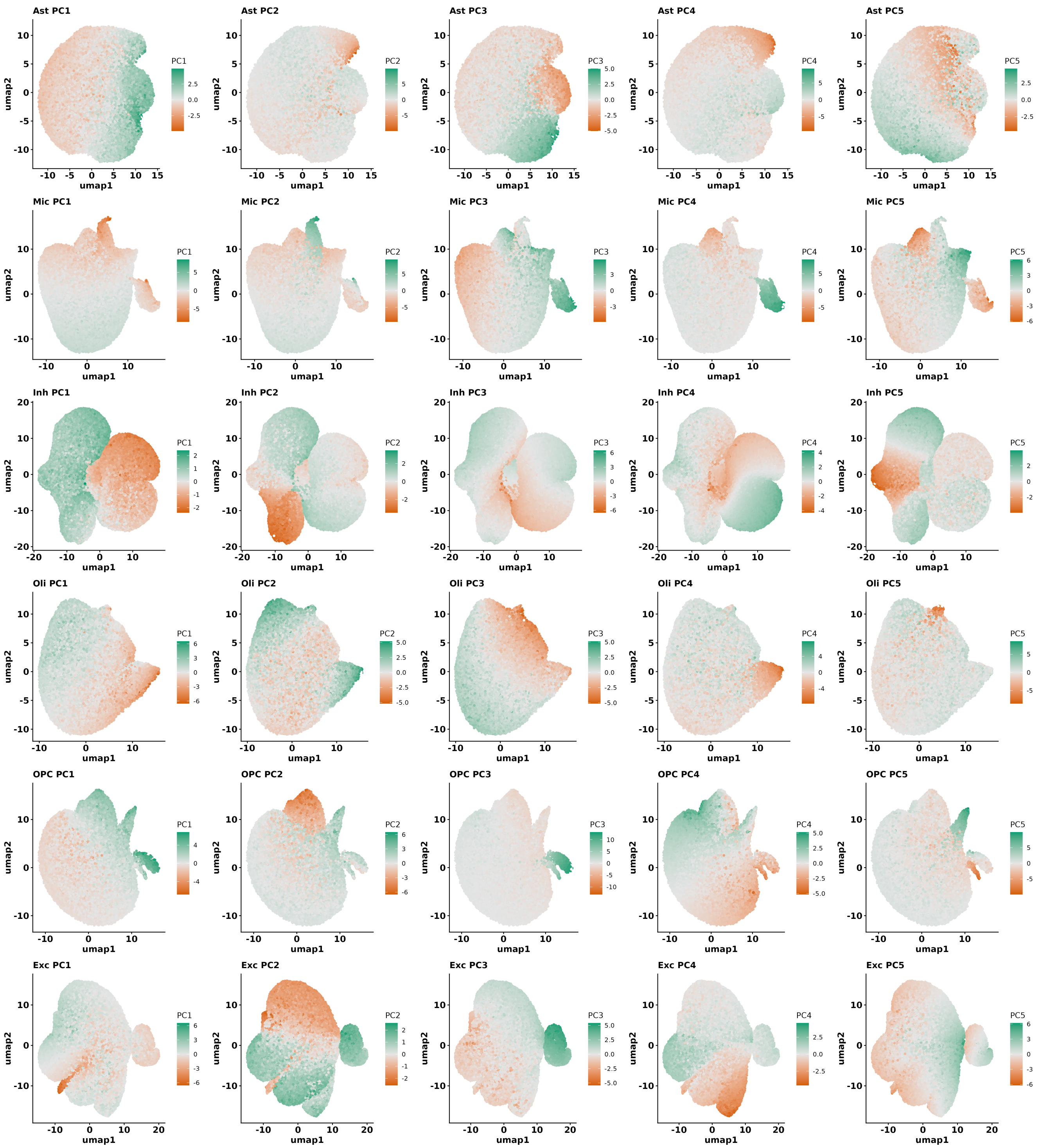

**Supplementary Fig. 10 | Cell-state axes inferred by within-cell-type principal component analysis.** UMAP embeddings of cells from each of the six major brain cell types in ROS/MAP snRNA-seq data (rows; Exc, excitatory neurons; Inh, inhibitory neurons; Ast, astrocytes; Oli, oligodendrocytes; OPC, oligodendrocyte precursor cells; Mic, microglia), coloured by each cell's score on the top five within-cell-type principal components (PC1–PC5; columns). These PCs define the continuous cell-state axes used in the G×Cell-State models. Green and orange indicate high and low PC scores, respectively; colour scales are centred on zero and differ among PCs. Each UMAP is computed separately within a cell type. Each point represents a cell.

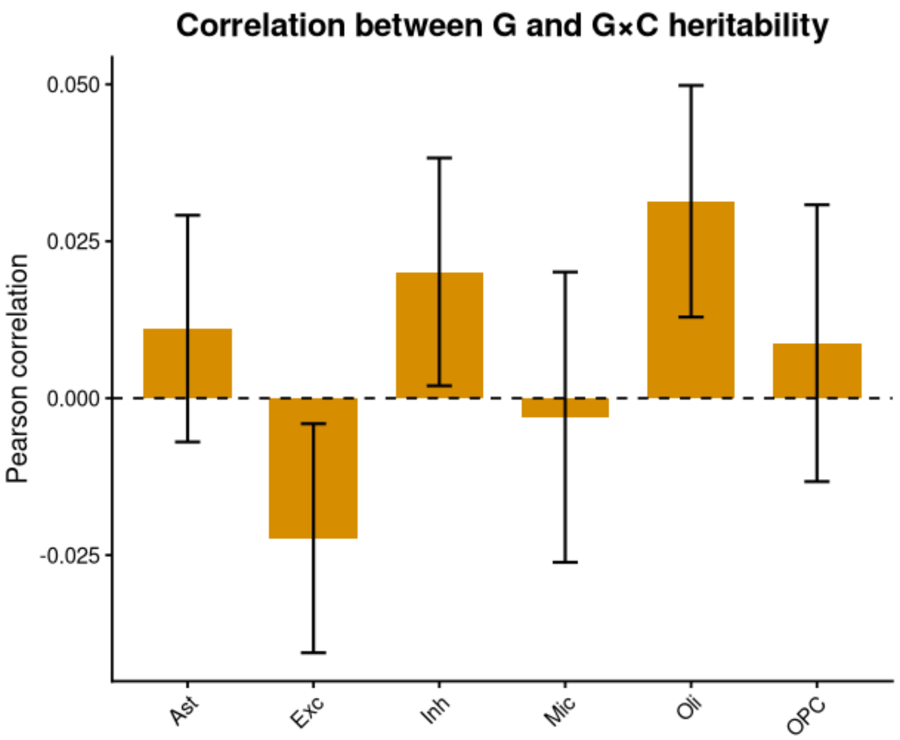


**Supplementary Fig. 11 | Main genetic and G×Cell-State heritability are largely uncorrelated.** Pearson correlation across genes between scHINT-G and scHINT-G×C heritability estimates in each cell type. Bars show correlation coefficients (range −0.022 to 0.031) and error bars show 95% CI; the dashed line marks zero. Genes with weak main genetic effects can therefore carry substantial G×Cell-State heritability, and vice versa.

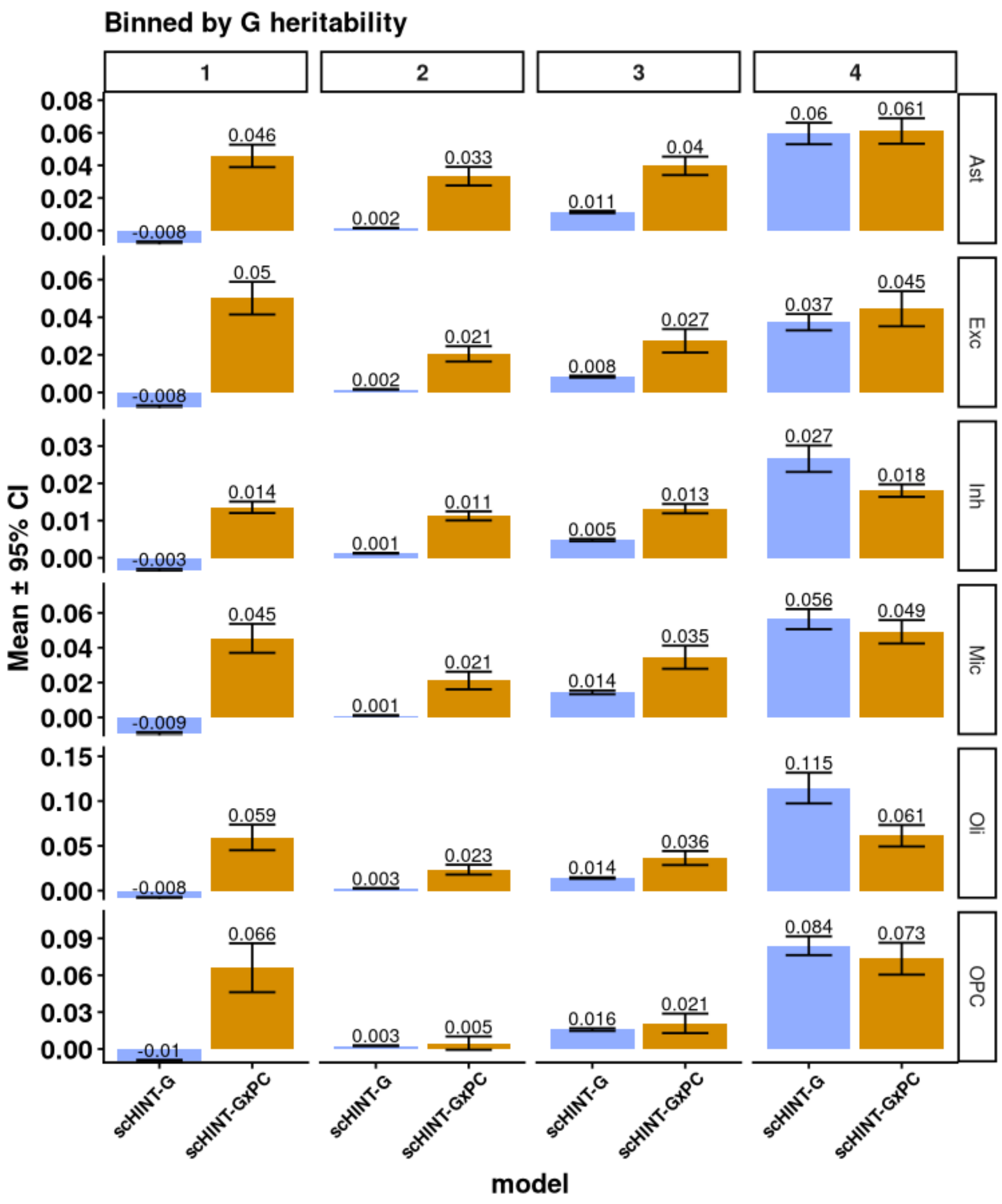


**Supplementary Fig. 12 | G×Cell-State heritability across levels of main genetic heritability.** Mean heritability estimated by scHINT-G (blue) and scHINT-G×PC (G×PC1–PC5 combined; gold) for genes grouped into four bins (1–4; columns) of increasing scHINT-G heritability, for each cell type. Numbers are mean estimates and error bars show 95% CI. G×Cell-State heritability is appreciable in all bins, including genes with negligible main genetic heritability (bin 1).

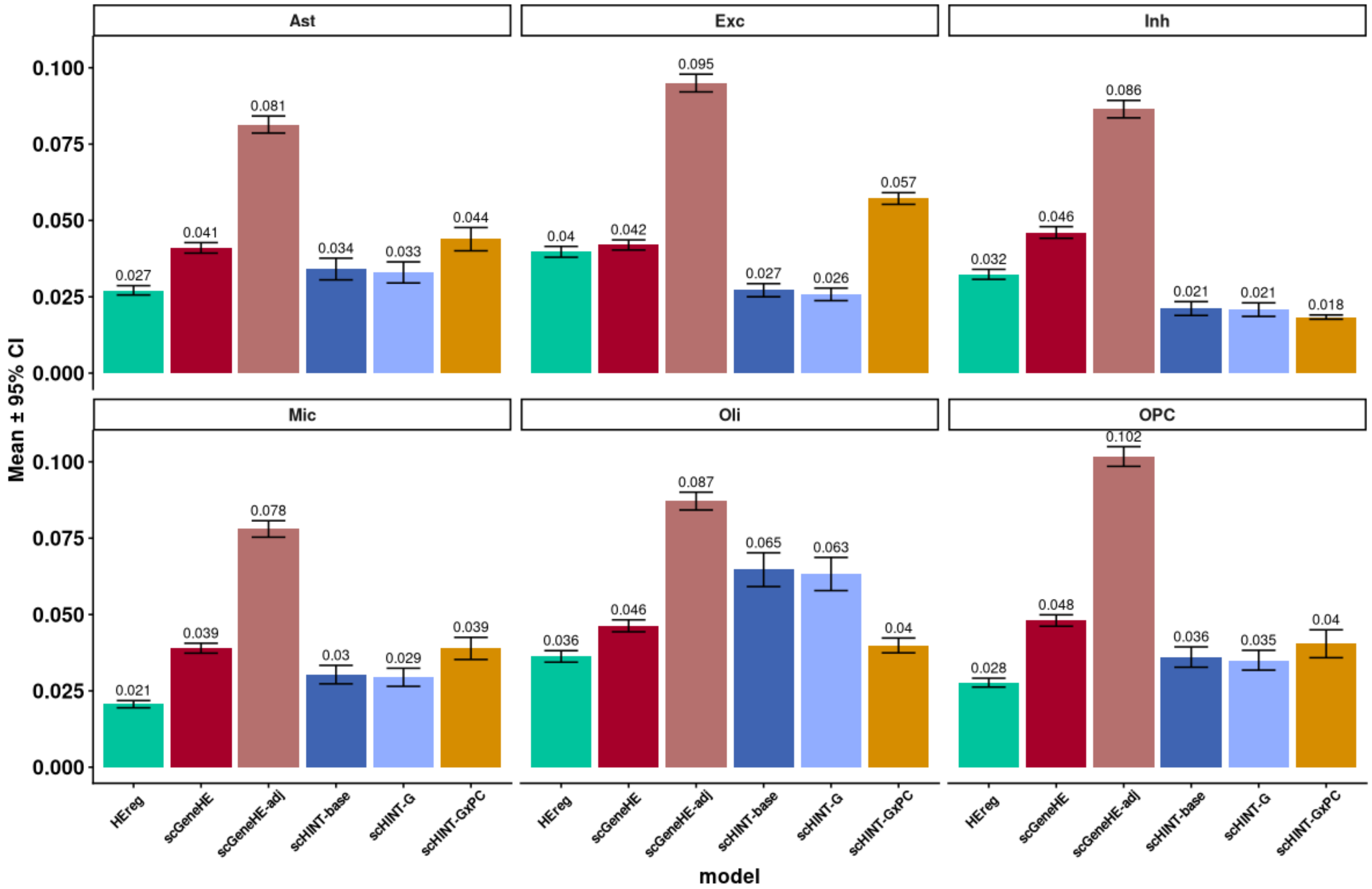


**Supplementary Fig. 13 | Comparison of scHINT with scGeneHE on meta-cell data.** Heritability estimated for each cell type from ~50,000 meta-cells per cell type, constructed by micro-clustering cells within donors. Bars show mean ± 95% CI across genes (numbers are means) for pseudo-bulk HE regression (HEreg), scGeneHE, scGeneHE-adj (scGeneHE adjusted for 25 pseudo-bulk expression PCs and five cell-level PCs), scHINT-base, scHINT-G and scHINT-G×PC. Covariate adjustment roughly doubles scGeneHE estimates, whereas unadjusted scGeneHE is closer to scHINT-G (Supplementary Note 5).

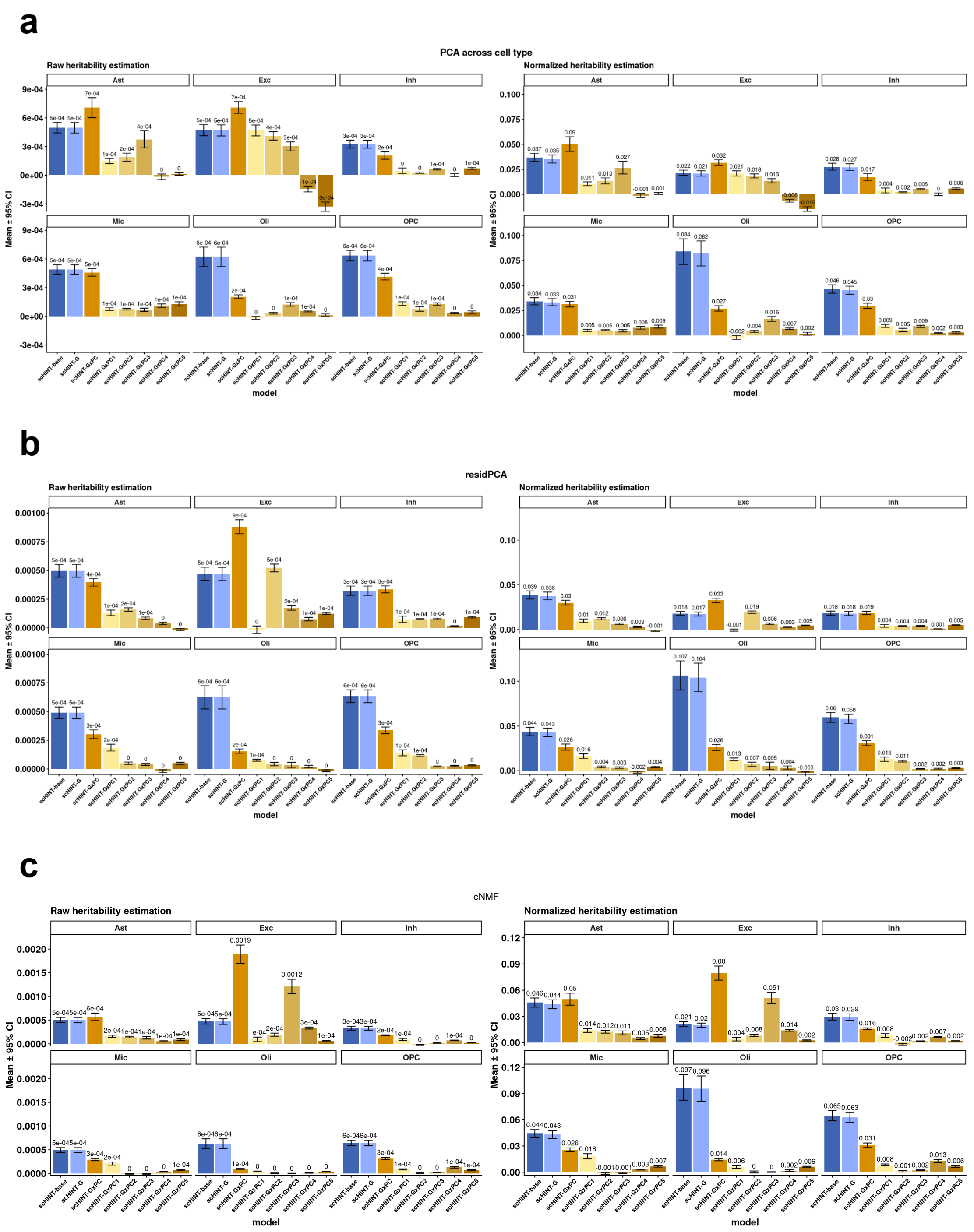


**Supplementary Fig. 14 | Robustness of G×Cell-State estimates to the definition of cell state.** Heritability estimated by scHINT when cell states were defined by across-cell-type PCA (**a**), residPCA (**b**) or consensus non-negative matrix factorization (cNMF; **c**). In each panel, the left half shows raw heritability (fraction of total cell-level variance) and the right half shows normalized heritability (cell-level residual variance excluded from the denominator) for each cell type. Bars show scHINT-base, scHINT-G, scHINT-G×PC (sum over components) and the individual components G×PC1–G×PC5 (for cNMF, the first five expression programs). Mean ± 95% CI; numbers are means. Main genetic heritability is consistent across definitions, whereas G×C estimates vary; across-cell-type PCA yields negative G×C estimates for several PCs in excitatory neurons, consistent with these PCs capturing cell-type identity.

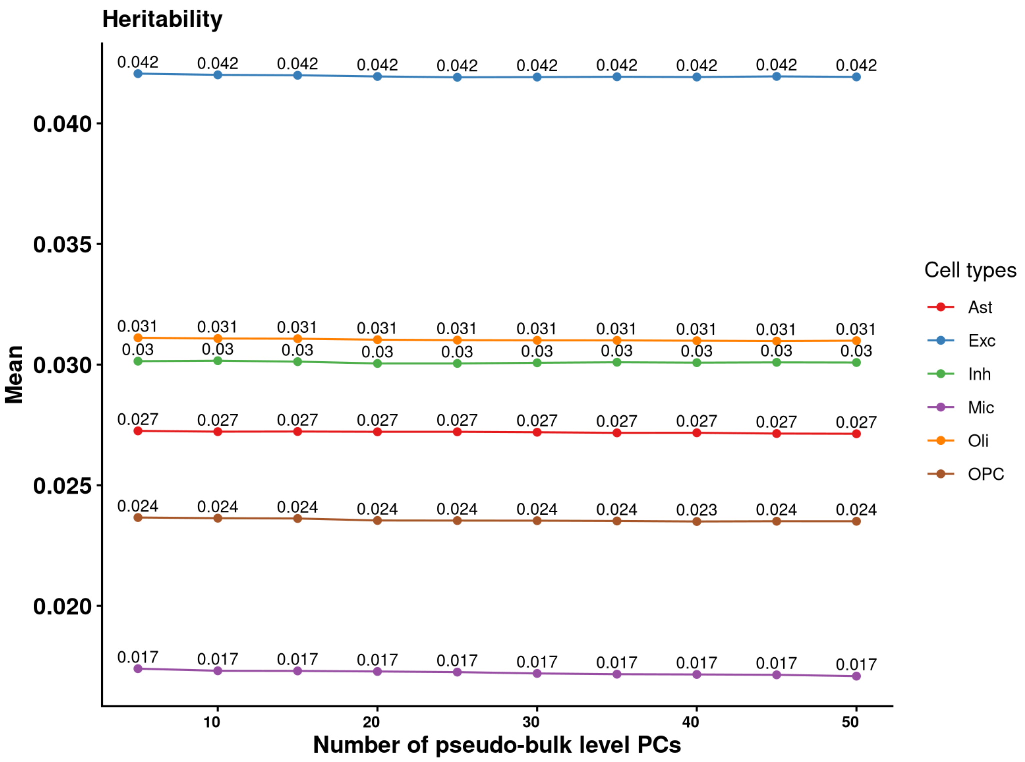


**Supplementary Fig. 15 | Stability of heritability estimates to the number of pseudo-bulk PCs.** Mean heritability estimated with 5 to 50 pseudo-bulk expression PCs (x axis) included as covariates, shown separately for each cell type (colours). Numbers are mean estimates. Estimates are essentially unchanged across the range, so 25 pseudo-bulk PCs were used in all analyses (Supplementary Note 6).

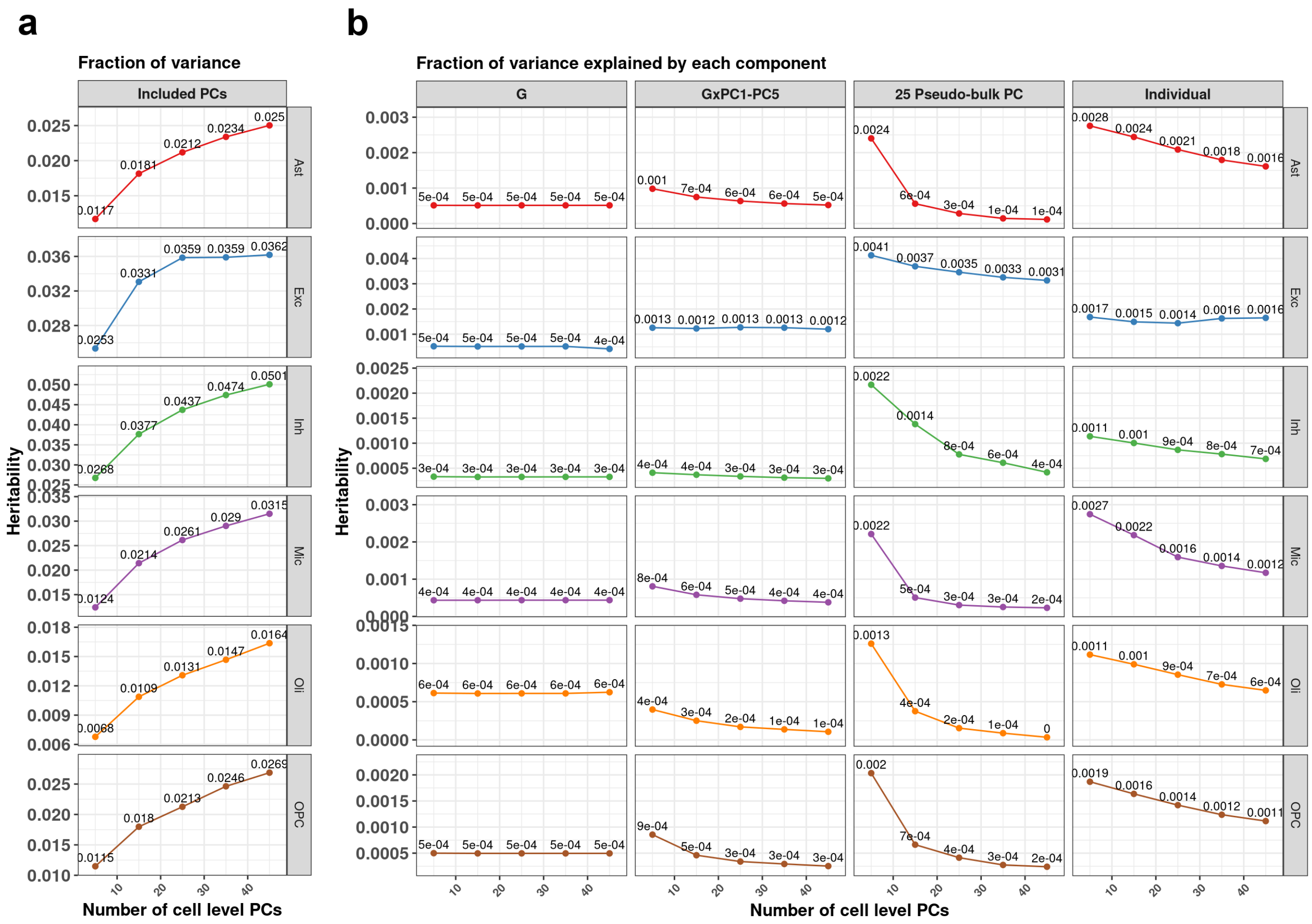


**Supplementary Fig. 16 | Variance components as a function of the number of cell-level PCs.** Models were fitted with 5 to 45 cell-level expression PCs (x axis) as covariates for each cell type. **a**, Fraction of variance attributed to the included cell-level PCs. **b**, Fraction of variance attributed to the main genetic effect (G), the summed G×PC1–PC5 interaction effects, the 25 pseudo-bulk PCs and the individual-level component. Numbers are mean estimates. The variance attributed to cell-level PCs increases with the number of PCs, whereas the variance attributed to pseudo-bulk PCs and the individual-level component decreases; the main genetic variance is stable and G×PC1–PC5 declines gradually in most cell types. Because cell-state variance enters the denominator of the cell-level heritability, raw heritability fractions change with the number of PCs (see Supplementary Fig. 17).

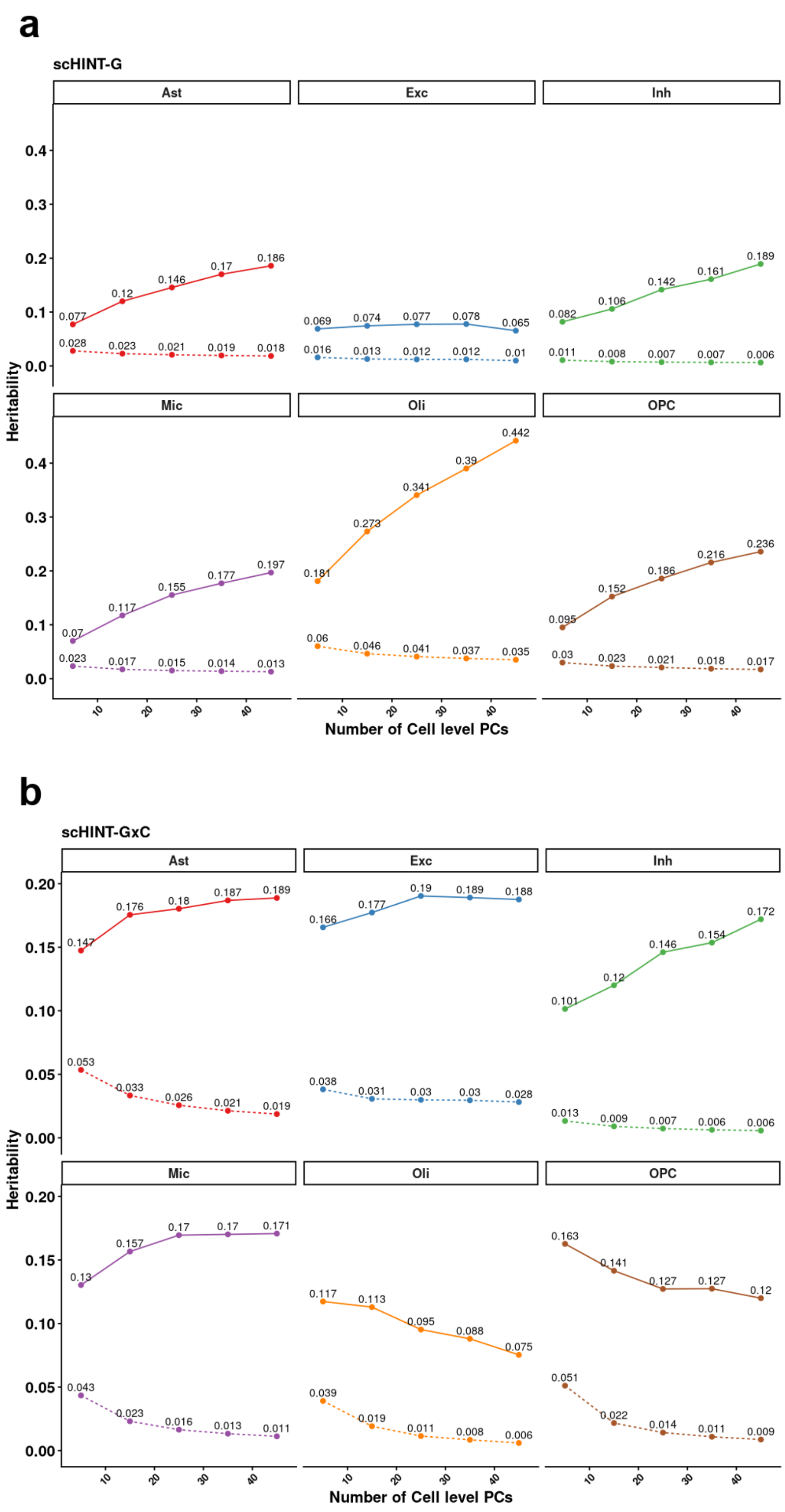


**Supplementary Fig. 17 | Normalized heritability bounds as a function of the number of cell-level PCs.** Normalized cell-level heritability of scHINT-G (**a**) and scHINT-G×C (**b**) estimated with 5 to 45 cell-level PCs (x axis), for each cell type. Dotted lines show the lower bound, in which only cell-level residual variance is excluded from the denominator; solid lines show the upper bound, in which both residual and cell-state variance are excluded. Pseudo-bulk heritability is expected to lie between the two bounds (Supplementary Note 2). Numbers are mean estimates.

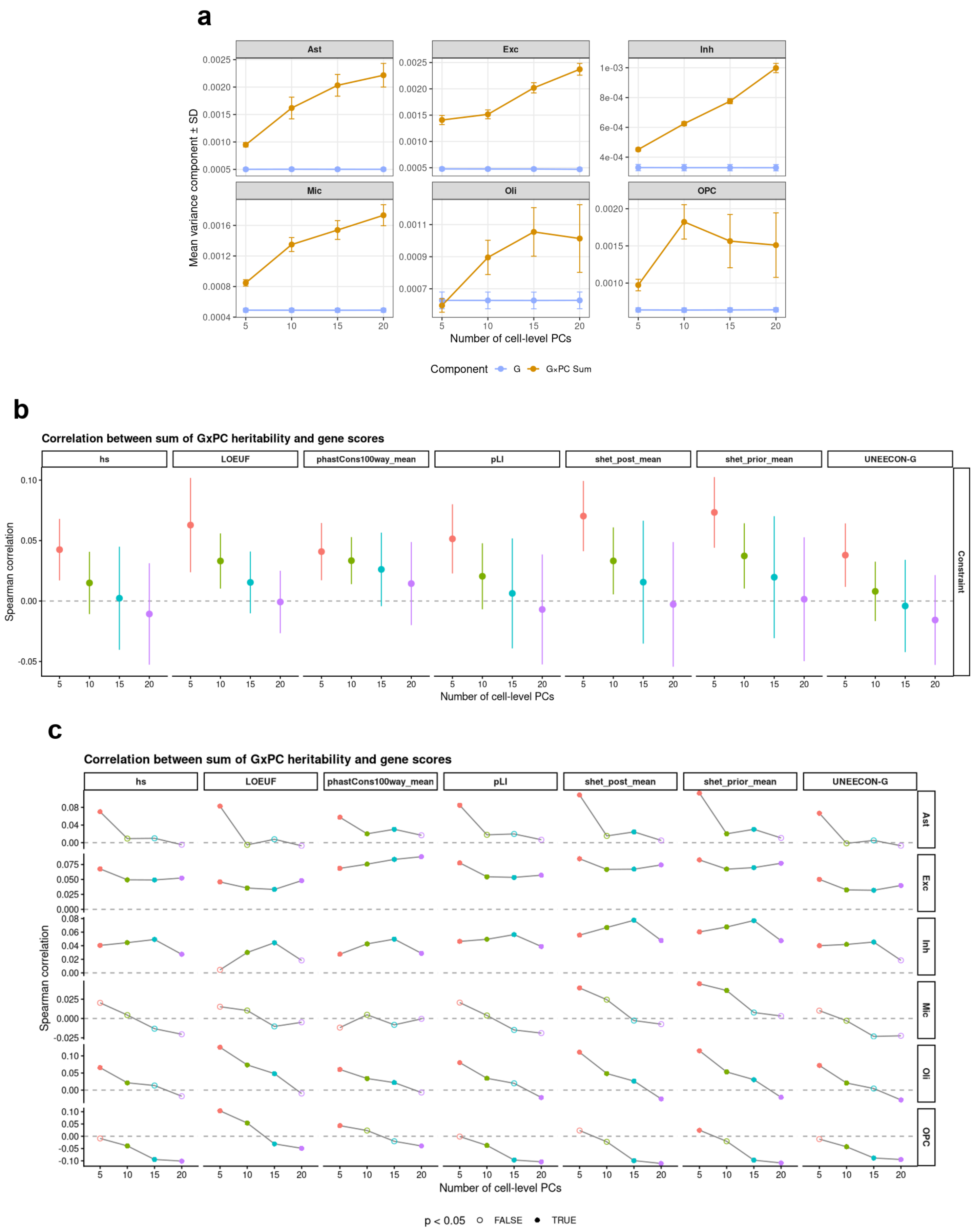

a
Ast
Exc
Inh
Mic
Oli
OPC
Mean variance component ± SD
Number of cell-level PCs
Component
G
G×PC Sum
b
Correlation between sum of GxPC heritability and gene scores
hs
LOEUF
phastCons100way_mean
pLI
shet_post_mean
shet_prior_mean
UNEECON-G
Constraint
Spearman correlation
Number of cell-level PCs
c
Correlation between sum of GxPC heritability and gene scores
hs
LOEUF
phastCons100way_mean
pLI
shet_post_mean
shet_prior_mean
UNEECON-G
Ast
Exc
Inh
Mic
Oli
OPC
Spearman correlation
Number of cell-level PCs
p < 0.05
FALSE
TRUE

**d**

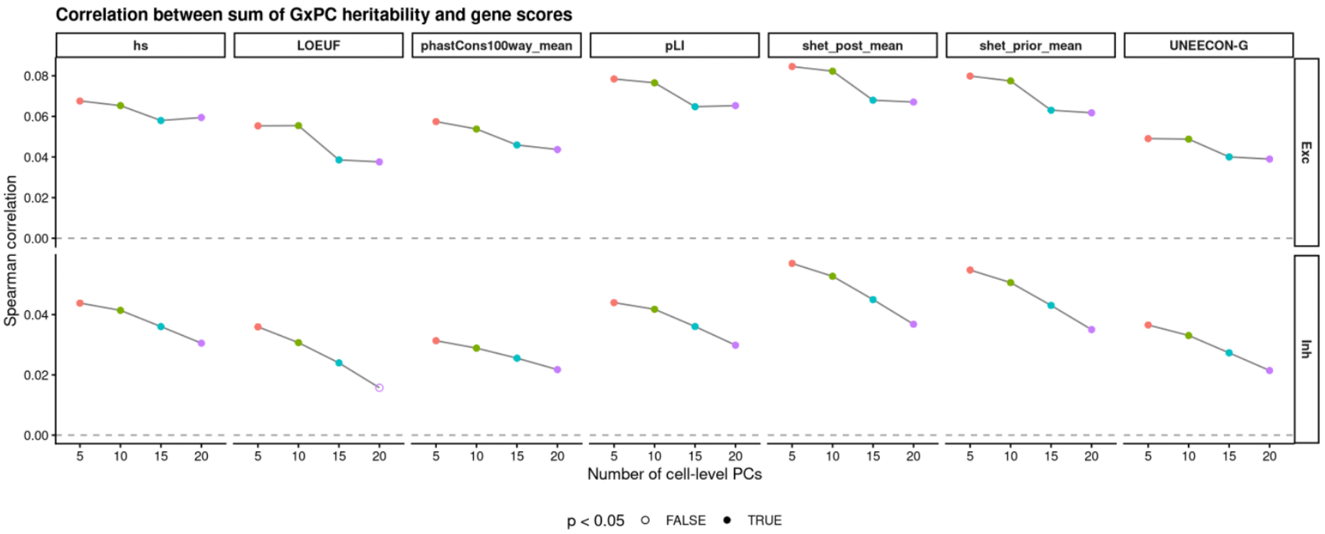


**Supplementary Fig. 18 | Choice of the number of G×PC interaction terms.** a. Mean variance component ± s.d. of the main genetic effect (G; blue) and of the summed G×PC interaction effects (G×PC sum; orange), estimated with 5, 10, 15 or 20 cell-state PCs included in the interaction component, for each cell type (Ast, astrocytes; Exc, excitatory neurons; Inh, inhibitory neurons; Mic, microglia; Oli, oligodendrocytes; OPC, oligodendrocyte precursor cells). b, Spearman correlation between the summed G×PC heritability and gene-level constraint scores (cell type mean and cell type specific), computed for 5, 10, 15 and 20 PCs (colours). c, Spearman correlation between the summed G×PC heritability and each constraint score in each cell type, computed with 5, 10, 15 and 20 PCs (colours). d, As in c, in excitatory and inhibitory neurons using a 10% subsample of cells.

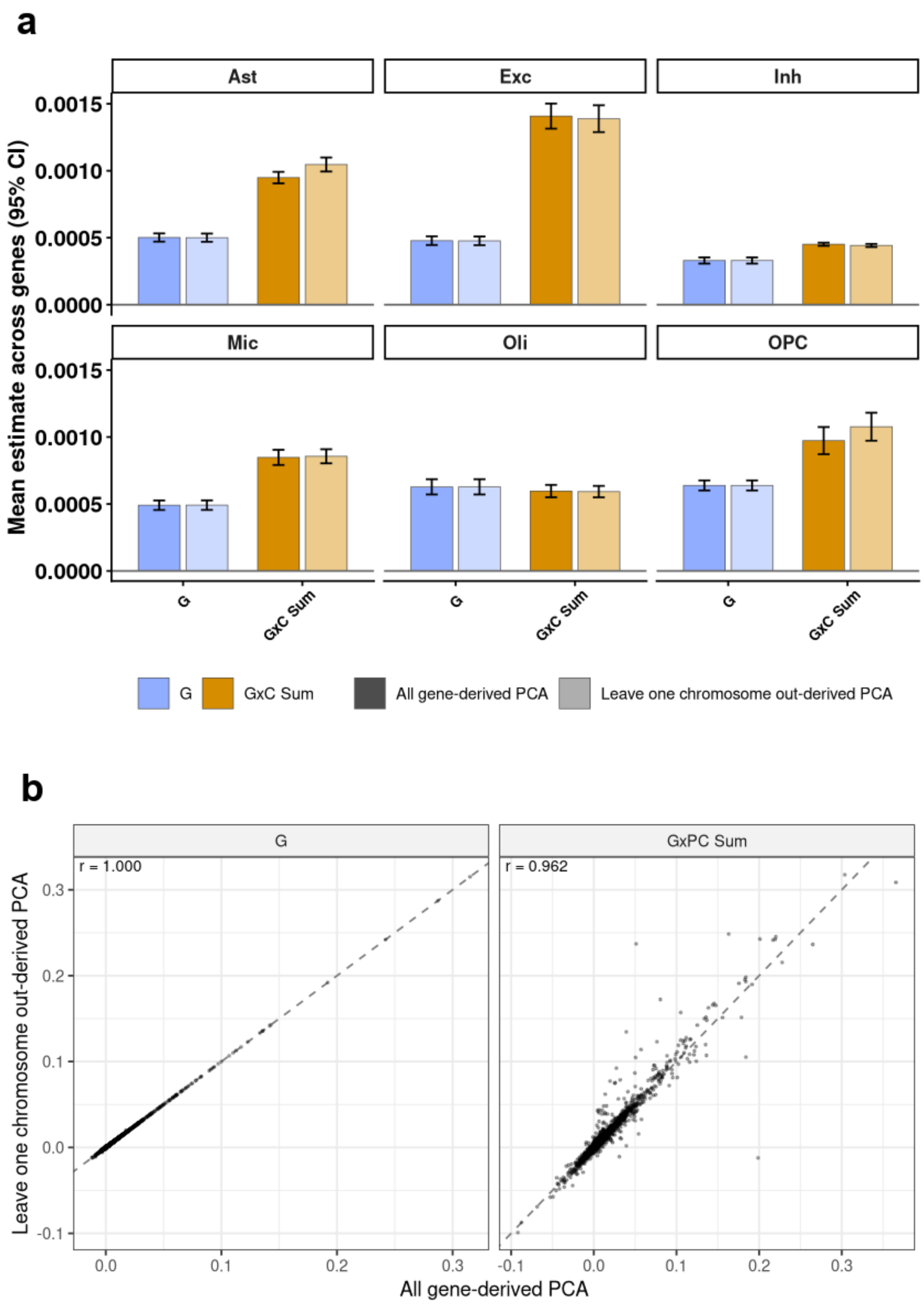


**Supplementary Fig. 19 | G×PC heritability estimated with PCs derived from all genes or with leave-one-chromosome-out PCs. a**, Mean estimate across genes (± 95% CI) of the main genetic variance component (G) and the summed G×PC variance component (G×C sum) in each cell type, using cell-state PCs derived from all genes (dark bars) or from all genes outside the chromosome of the focal gene (leave-one-chromosome-out PCA; light bars). b. Gene-level estimates of G (left) and G×PC sum (right) from all-gene-derived PCs (x axis) versus leave-one-chromosome-out PCs (y axis). Each point is a gene, dashed lines show y = x, and r is the Pearson correlation.

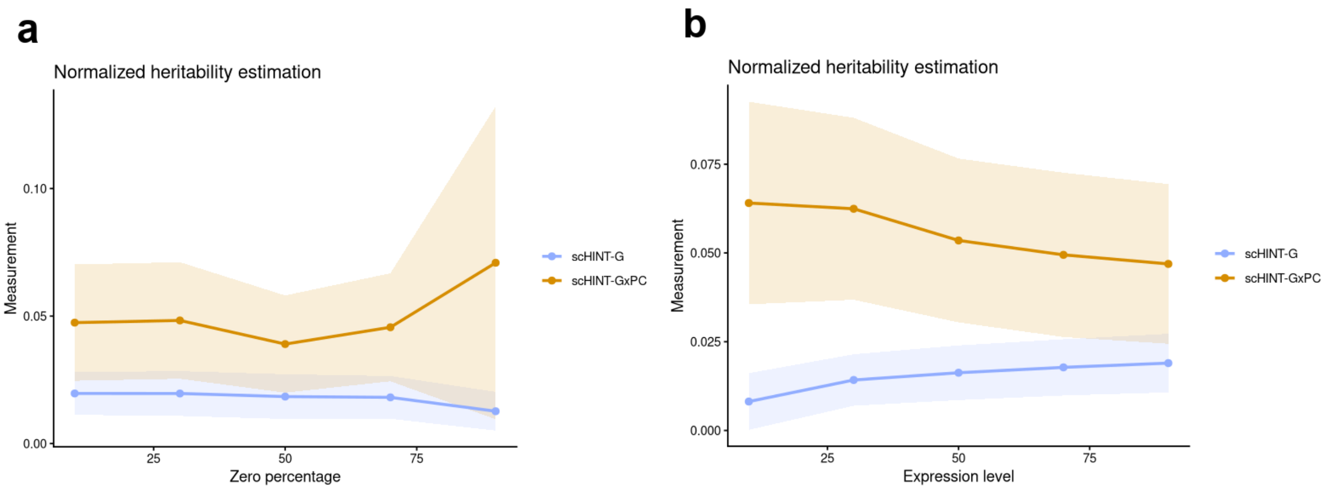


**Supplementary Fig. 20 | Robustness of scHINT to artificially increased sparsity and sequencing depth. a.** Normalized heritability of scHINT-G (blue) and scHINT-G×PC (gold) after randomly setting 10–90% of cell-level expression measurements to zero (x axis, zero percentage); cell-state PCs were recomputed from the perturbed expression matrix. Lines show mean estimates and shaded bands show 95% interval across genes in Microglia. Estimates are stable up to moderate sparsity; at very high sparsity the G×Cell-State component is inflated and the main genetic component is modestly reduced. **b.** Normalized heritability of scHINT-G (blue) and scHINT-G×PC (gold) estimated separately in cells stratified into library-size quantile bins (x axis, library-size percentile; low to high sequencing depth). Lines show mean estimates and shaded bands show 95% interval across genes in Microglia. Cells with lower library size show a modestly smaller main genetic component and a slightly larger G×Cell-State component; estimates from higher-depth cells are similar to the overall results.

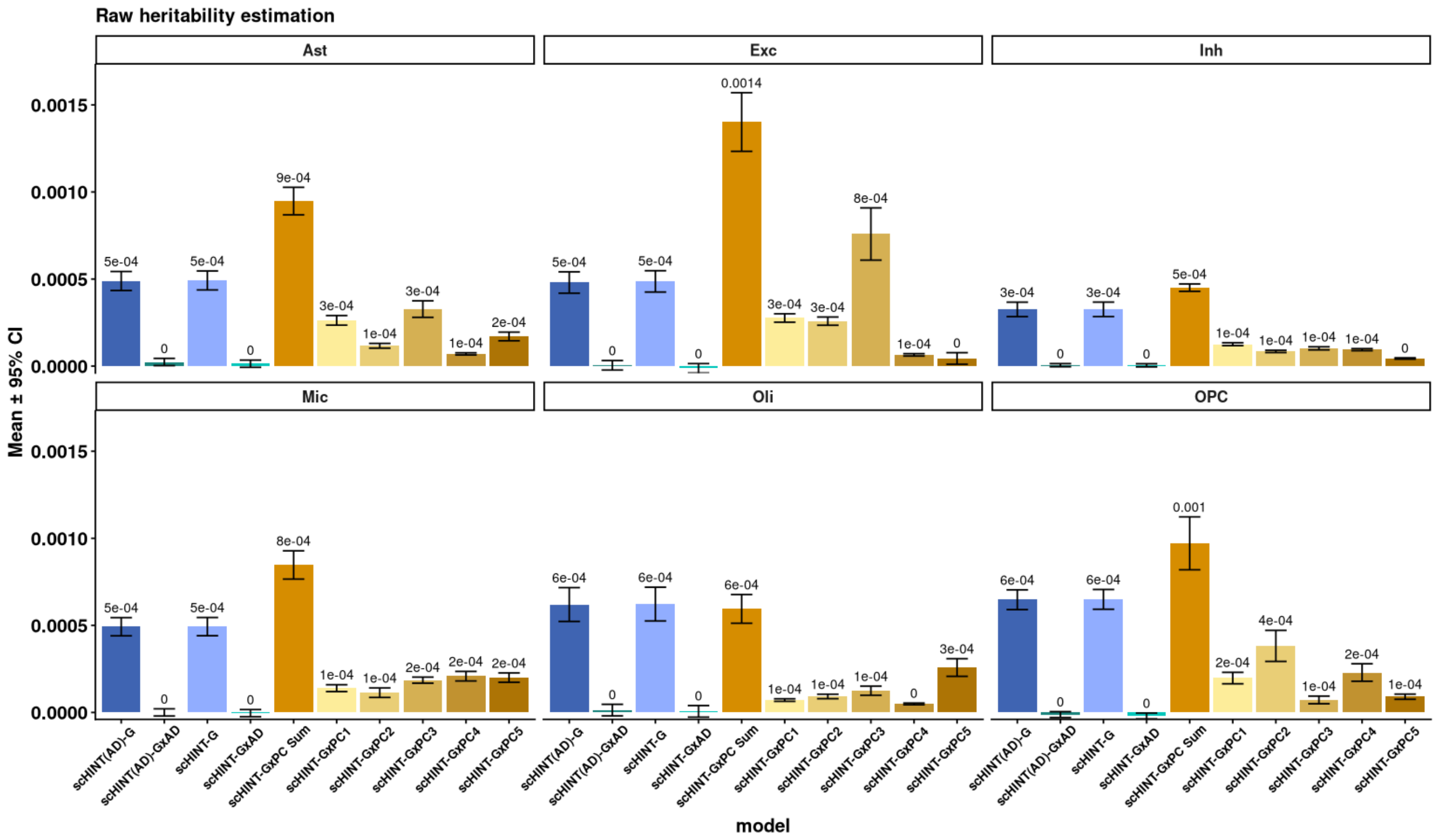


**Supplementary Fig. 21 | Sensitivity of heritability estimates to Alzheimer's disease status.** Raw heritability (variance component; mean ± 95% CI across genes) estimated in each cell type for scHINT models that include AD status as an additional context variable. Bars show the main genetic component (G) and the G×AD interaction component in the baseline model with AD status and in the full model with AD status, together with the summed G×PC component (G×PC sum) and the individual components G×PC1–G×PC5 of the full model. Numbers above bars are mean estimates.

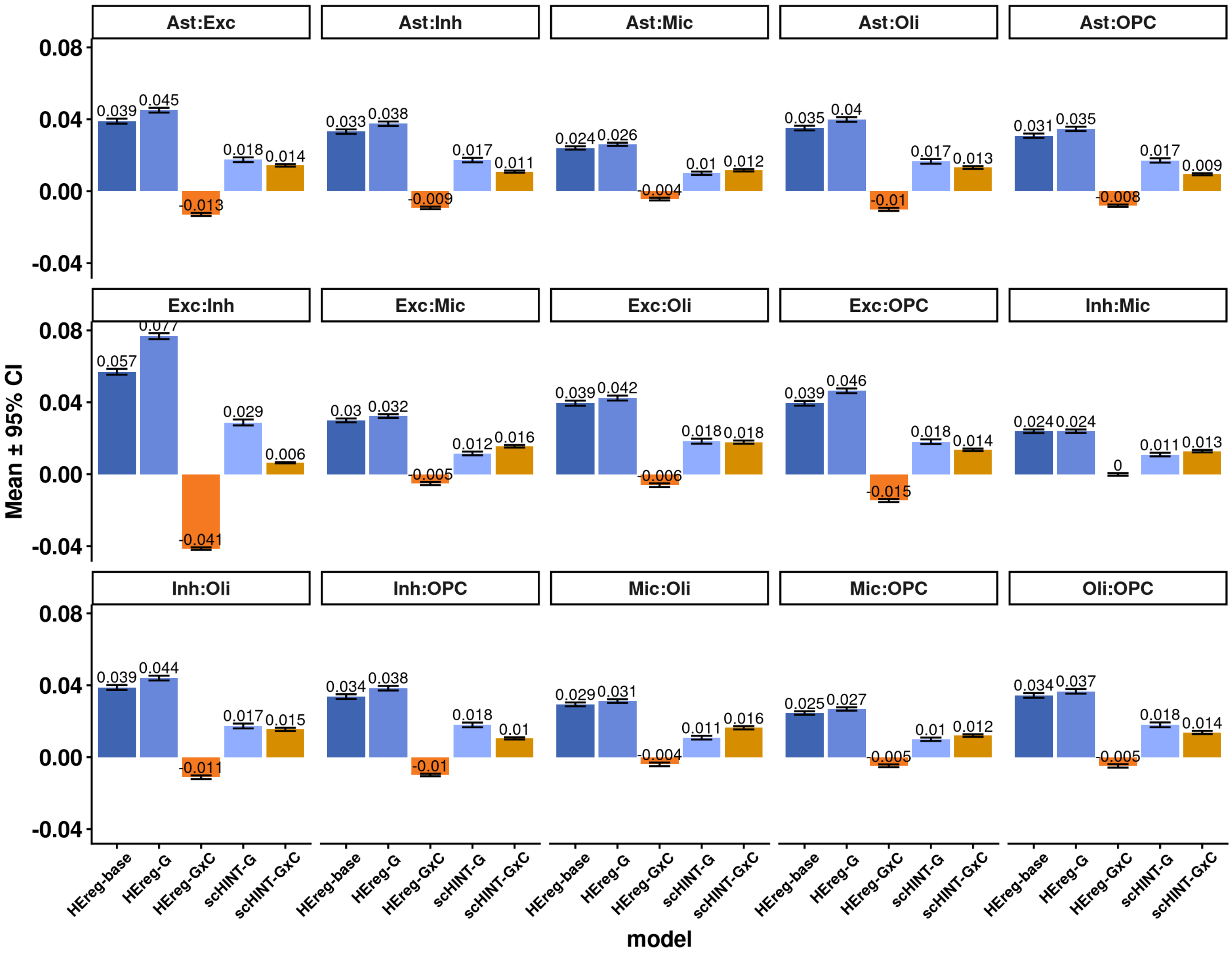


**Supplementary Fig. 22 | Pairwise G×Cell-Type analysis across brain cell types.** Heritability estimates for each of the 15 pairs of the six major brain cell types (panels). Bars show mean ± 95% CI (numbers are means) for HEreg-base (main genetic component only), HEreg-G and HEreg-G×C (main genetic and G×Cell-Type components from a joint model without cell-type and individual-level variance components) and scHINT-G and scHINT-G×C (full model that additionally includes marginal cell-type and individual-level variance components). Omitting these components inflates G and deflates, sometimes below zero, the G×Cell-Type estimate, whereas scHINT gives positive, stable G×Cell-Type estimates.

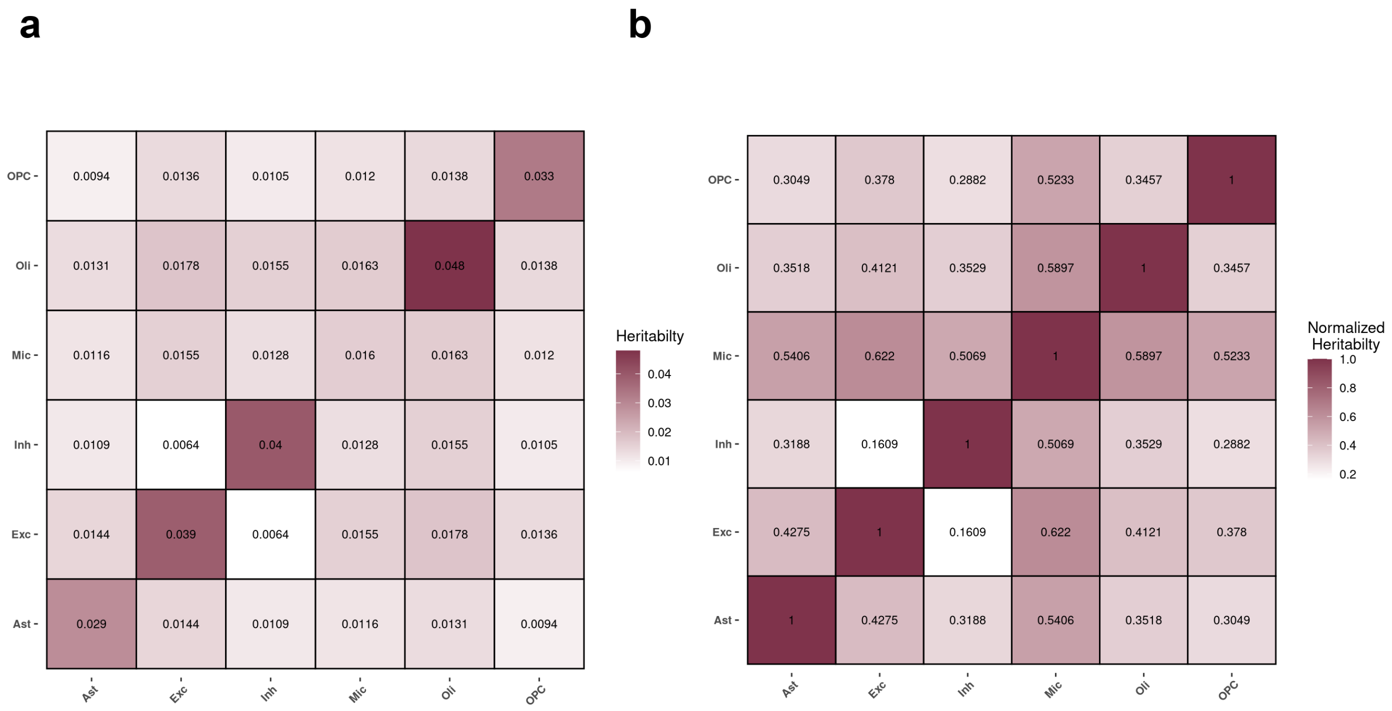


**Supplementary Fig. 23 | G×Cell-Type heritability across cell-type pairs. a**, Mean scHINT G×Cell-Type heritability for each pair of cell types (off-diagonal); the diagonal shows main genetic heritability within each cell type. **b**, G×Cell-Type heritability normalized by the mean main genetic heritability of the two cell types in each pair (diagonal = 1). Darker colours indicate higher values. Excitatory–inhibitory neurons show the lowest G×Cell-Type heritability, and pairs of more distinct cell types (for example, neurons and microglia) show higher values.

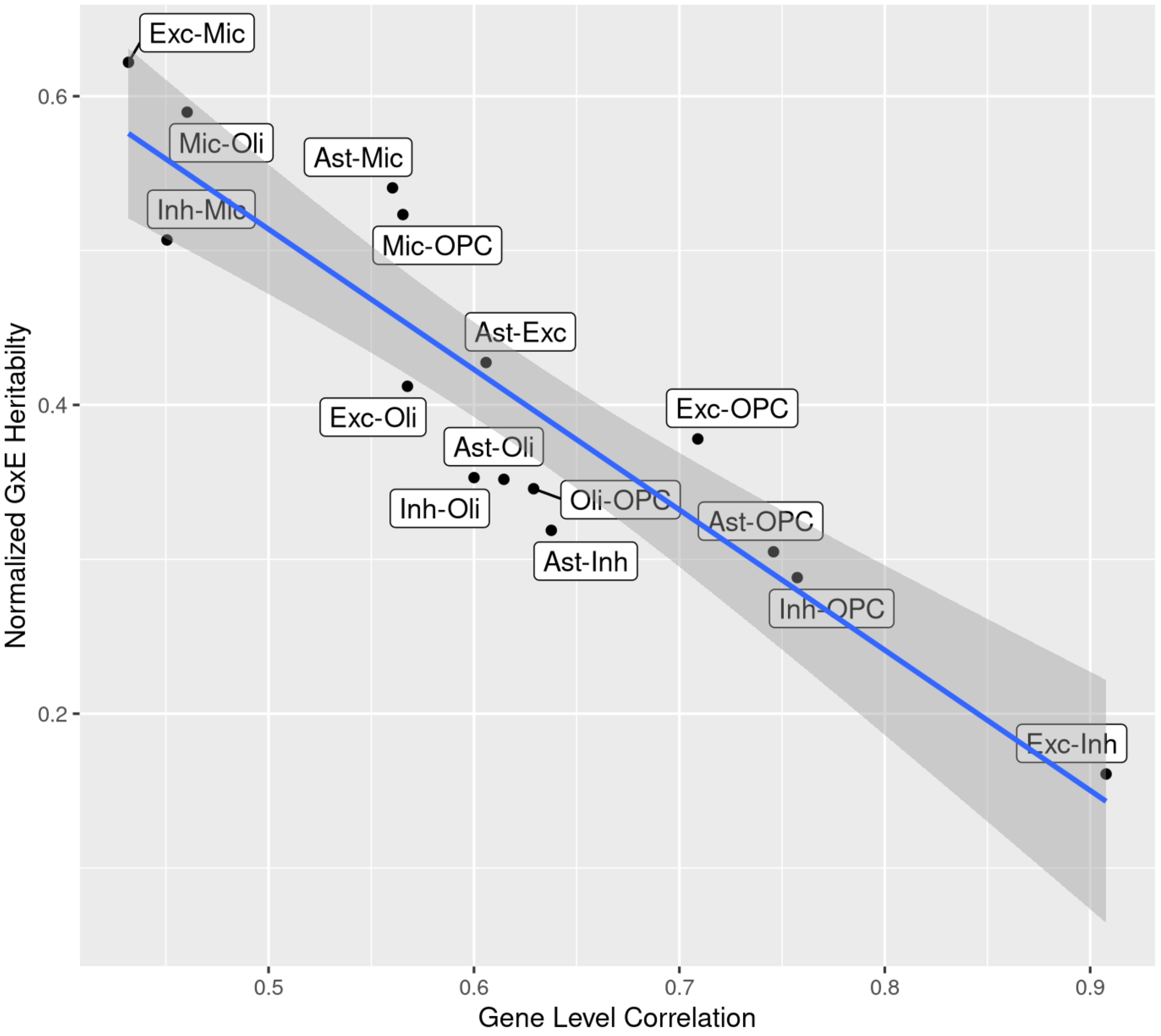


**Supplementary Fig. 24 | Normalized G×Cell-Type heritability decreases with transcriptional similarity of cell types.** Each point represents a pair of cell types. The x axis shows the gene-level expression correlation between the two cell types and the y axis shows G×Cell-Type heritability normalized by the mean main genetic heritability of the pair. The blue line is a linear fit and the grey band its 95% confidence interval. Transcriptionally similar pairs (such as excitatory and inhibitory neurons) have weaker relative G×Cell-Type effects than divergent pairs.

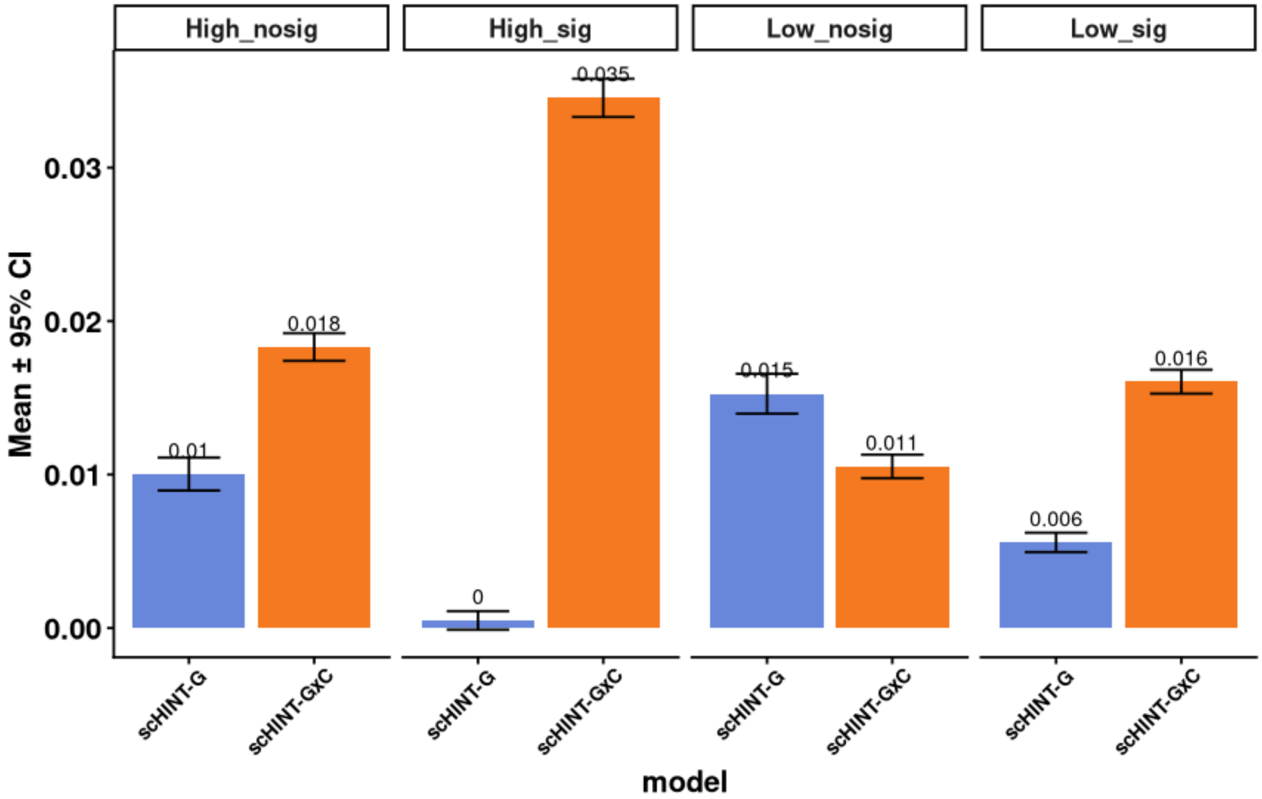


**Supplementary Fig. 25 | G×Cell-Type heritability by expression level and differential expression between cell types.** Mean scHINT-G and scHINT-G×C heritability (± 95% CI; numbers are means) for genes stratified by expression level (High or Low compared to mean) and by whether their expression differs significantly between the two cell types (sig) or not (nosig). G×Cell-Type heritability is highest for highly expressed genes with significant expression differences, linking cell-type-dependent genetic regulation to transcriptional divergence.

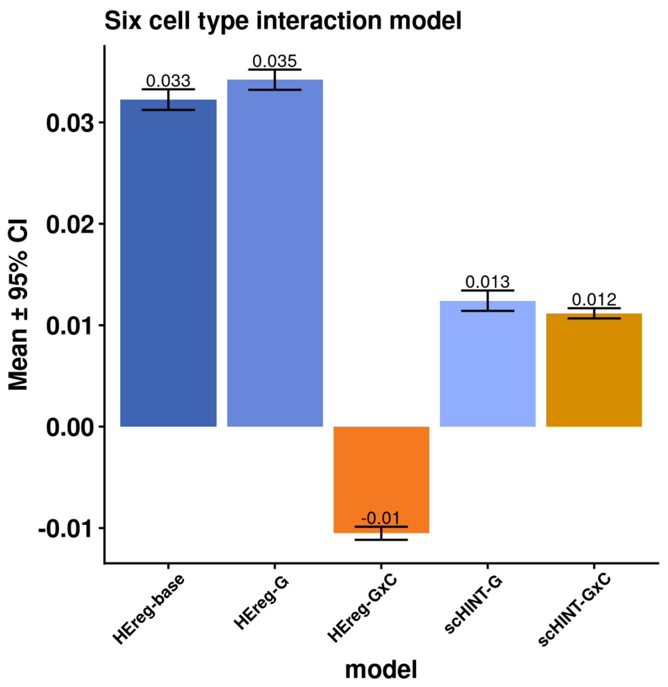


**Supplementary Fig. 26 | Joint G×Cell-Type analysis across six cell types.** Heritability estimated by modelling genetic heterogeneity jointly across all six major brain cell types (Exc, Inh, Ast, Oli, OPC, Mic). Bars show mean ± 95% CI (numbers are means) for HEreg-base (main genetic component only), HEreg-G and HEreg-G×C (joint model without cell-type and individual-level variance components) and scHINT-G and scHINT-G×C (full model). Accounting for cell-type and individual-level variance reduces the main genetic estimate and yields a positive G×Cell-Type estimate.

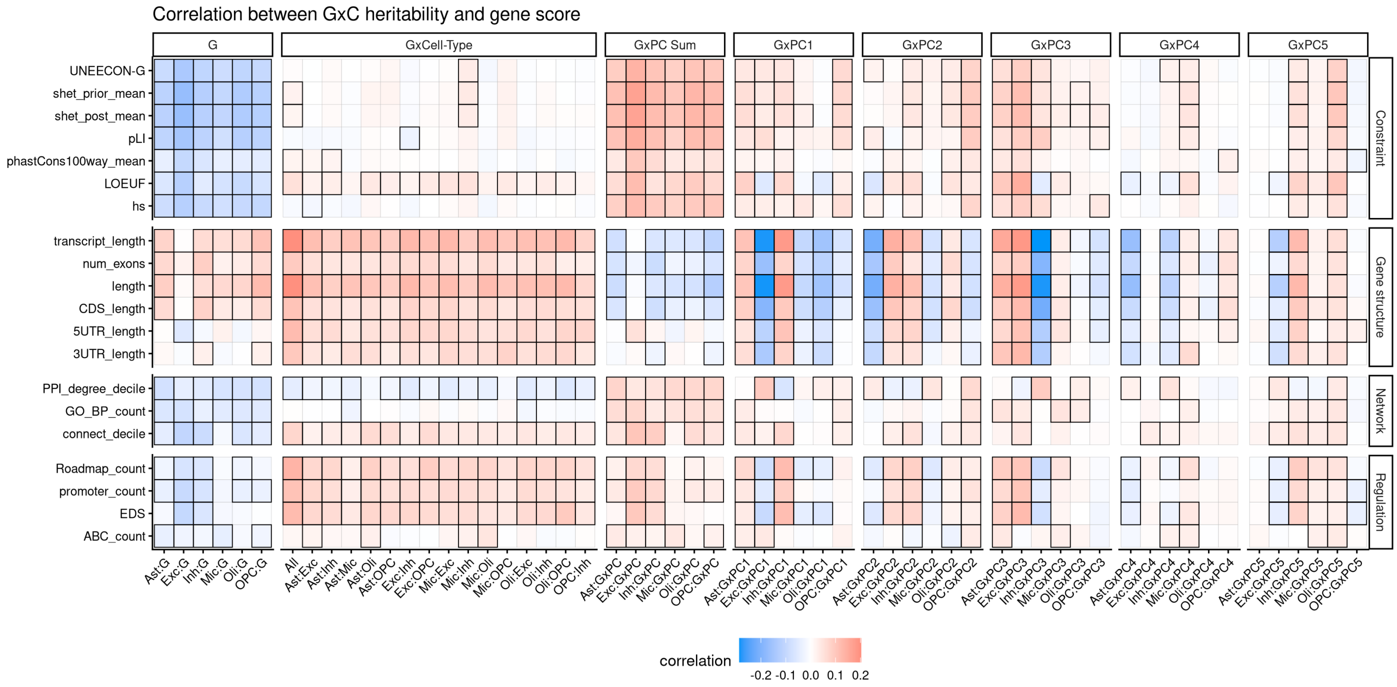


**Supplementary Fig. 27 | Associations between the proportion of heritability attributable to G, G×Cell-Type and G×Cell-State and gene-level annotations.** Correlation (standardized regression coefficient from rank-transformed variables) between the proportion of heritability attributed to each component and gene-level scores in ROS/MAP brain cell types. Rows are gene scores grouped into constraint (UNEECON-G, $s_{het}$ prior and posterior mean, pLI, phastCons, LOEUF, hs), gene structure, network and regulation annotations; columns are heritability components: main genetic (G) for each cell type, G×Cell-Type for all cell types combined and each cell-type pair, summed G×PC (GxPC Sum) and each of G×PC1–G×PC5 for each cell type (Exc, excitatory neurons; Inh, inhibitory neurons; Ast, astrocytes; Oli, oligodendrocytes; OPC, oligodendrocyte precursor cells; Mic, microglia). Colour indicates the correlation (red, positive; blue, negative) and black borders mark significant correlations (FDR < 0.05). Interaction components were adjusted for the corresponding main genetic heritability. G is negatively and G×Cell-State positively associated with constraint.

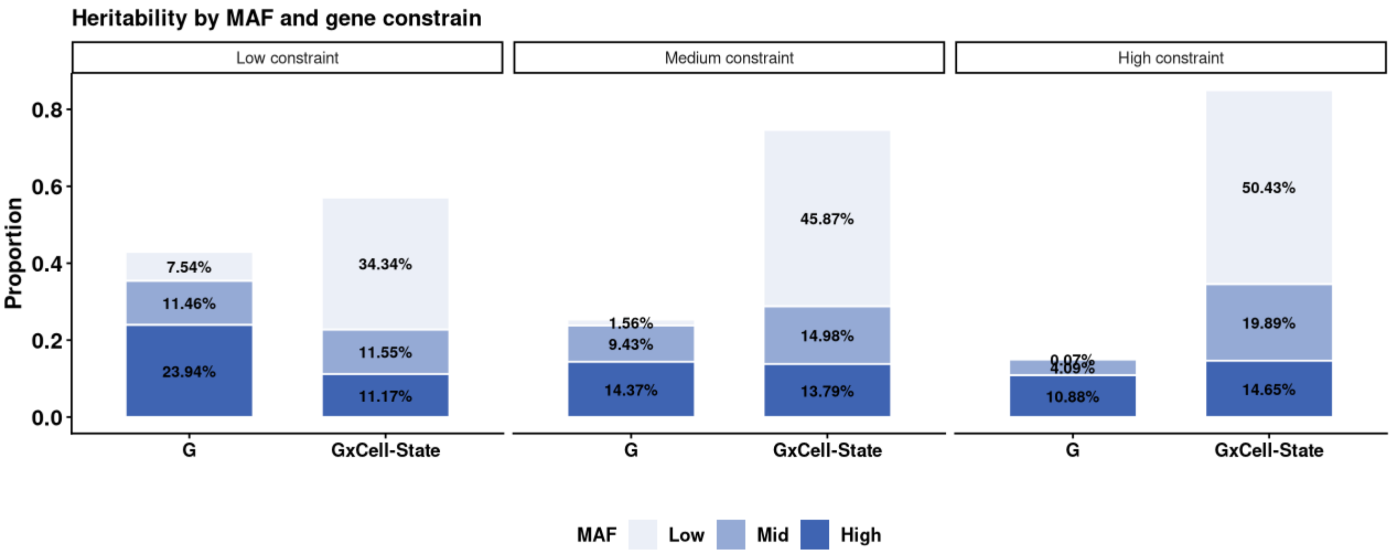


**Supplementary Fig. 28 | Allele-frequency architecture of G and G×Cell-State heritability by gene constraint.** Proportion of total heritability (G + G×Cell-State) contributed by variants in each minor allele frequency (MAF) bin (low, 1–5%; mid, 5–20%; high, > 20%) for the main genetic (G) and G×Cell-State components in genes with low, medium and high evolutionary constraint (pLI; low and high constraint, bottom and top 20% of genes). Percentages are shown within bars. G×Cell-State heritability is dominated by low-frequency variants, increasingly so in constrained genes, whereas main genetic heritability is dominated by common variants.

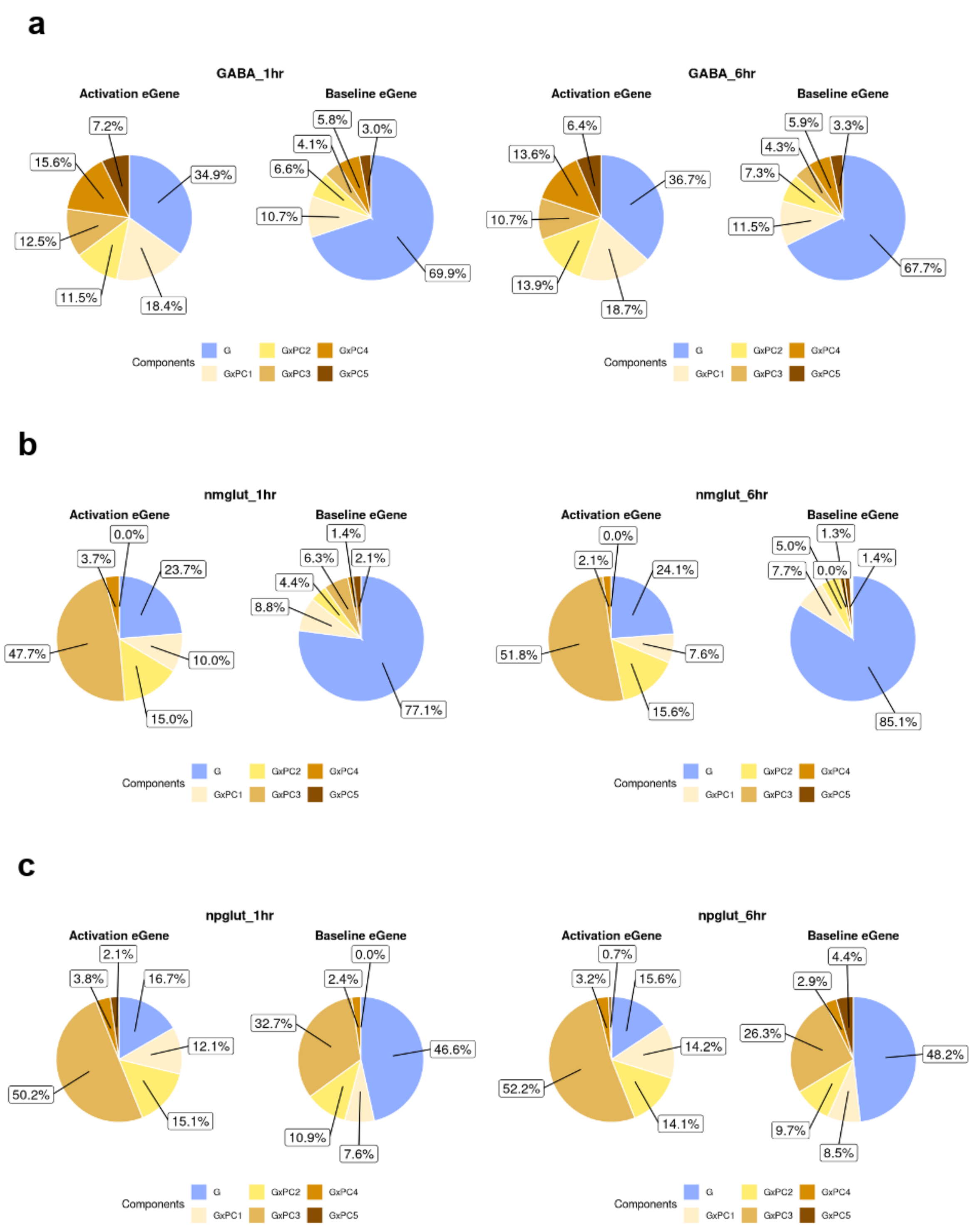


**Supplementary Fig. 29 | Activation-dependent eGenes show a larger G×Cell-State fraction in neurons.** Relative contribution (%) of heritability components (main genetic, G; G×PC1–G×PC5) estimated by scHINT in ROS/MAP neurons, for eGenes specific to neuronal activation (Activation eGene) and for baseline eGenes (Baseline eGene) identified in iPSC-derived neurons of Liang et al. after 1 h or 6 h of potassium chloride stimulation. **a**, GABA+ inhibitory neurons (compared with ROS/MAP inhibitory neurons). **b**, nmGlut, excitatory neurons characterized by a low-neurofilament program. **c**, npGlut, the other excitatory neuron population (b and c compared with ROS/MAP excitatory neurons). G×C components account for a larger share of heritability among activation-specific eGenes.

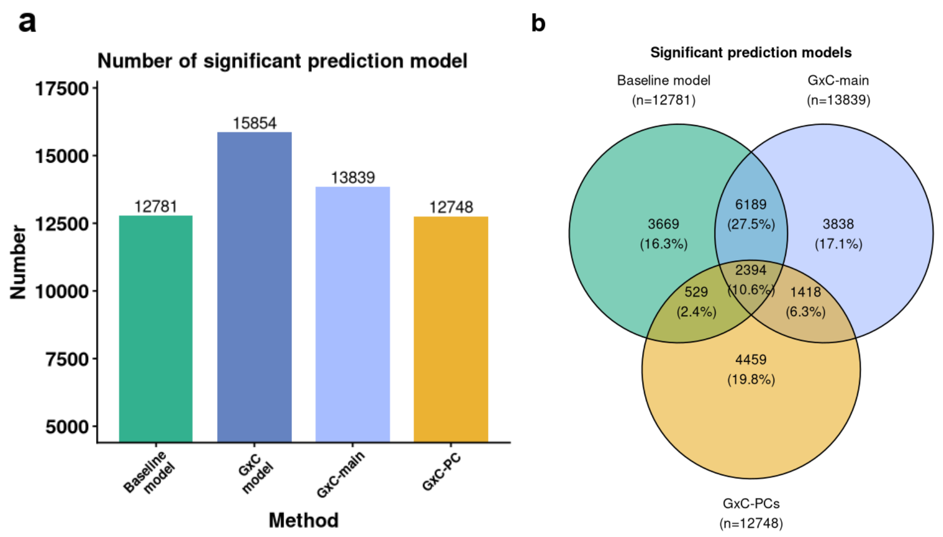


**Supplementary Fig. 30 | G×Cell-State models improve cell-level expression prediction.** Elastic-net prediction models of cell-level expression (metacells) were trained for each gene and cell type and evaluated in held-out donors with a mixed model including a donor random intercept (a model is significant if the combined coefficient is positive and $P < 0.05$). **a**, Number of significant prediction models across the six cell types for the baseline model (genotype only), the combined G×Cell-State model (G×C model), its main-effect weights (G×C-main) and its interaction weights (G×C-PC). **b**, Overlap of significant prediction models among the baseline model, G×C-main and G×C-PCs; numbers are model counts (% of the union).

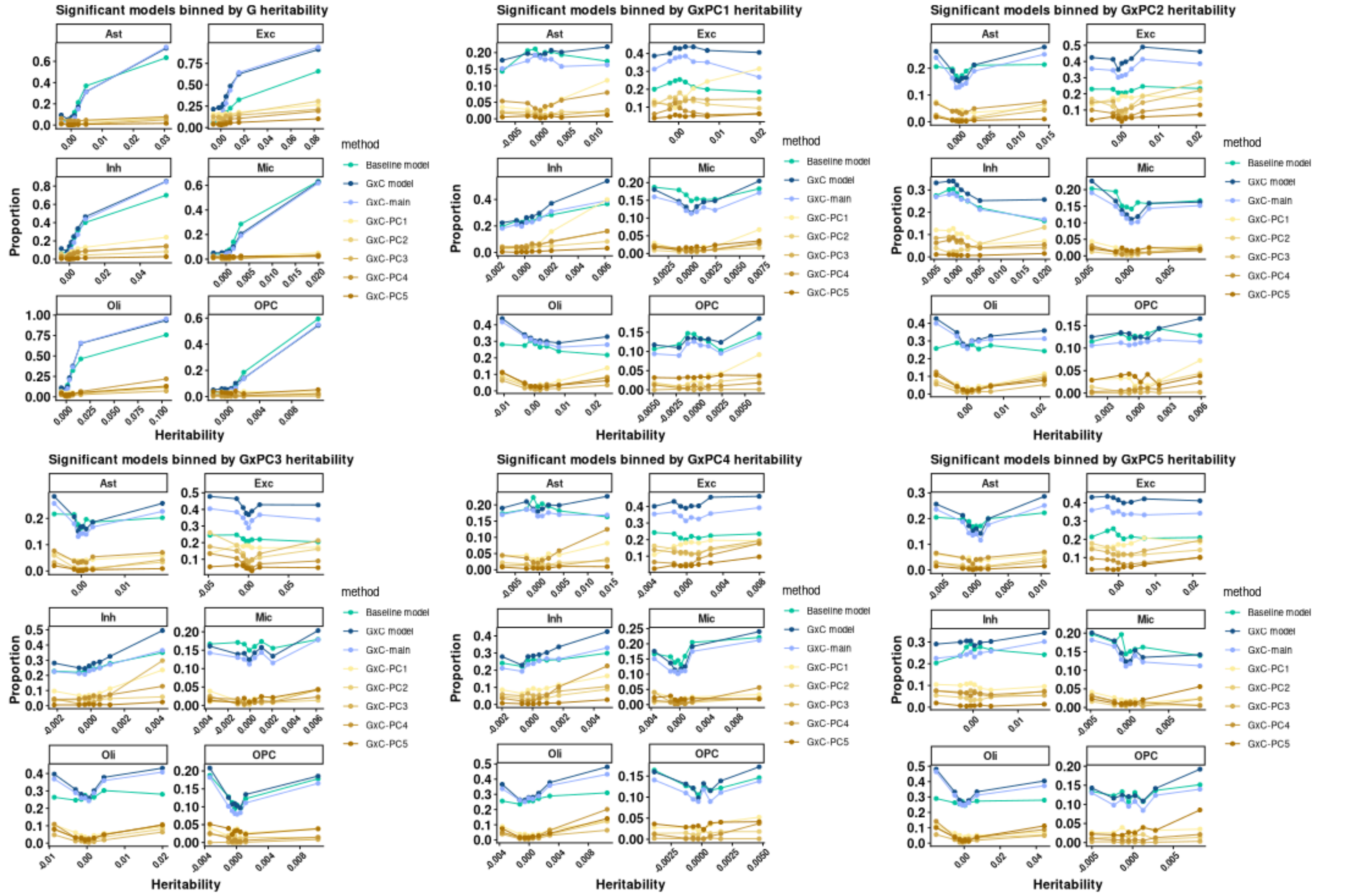


**Supplementary Fig. 31 | Proportion of significant prediction models by heritability.** Proportion of gene–cell-type models with significant out-of-sample prediction (y axis) for genes binned by scHINT heritability (x axis; bin mean heritability) of the main genetic component (top left) or of G×PC1 to G×PC5 (other panels), shown for each cell type (Ast, Exc, Inh, Mic, Oli, OPC). Lines show the baseline model, the combined G×Cell-State model (G×C model), its main-effect weights (G×C-main) and the interaction weights for each PC (G×C-PC1–PC5). Genes with higher G×PC1 heritability have more significant G×PC1 prediction weights, but this does not hold for main-effect weights or other PCs, indicating that interaction signal is concentrated in specific cell-state axes.

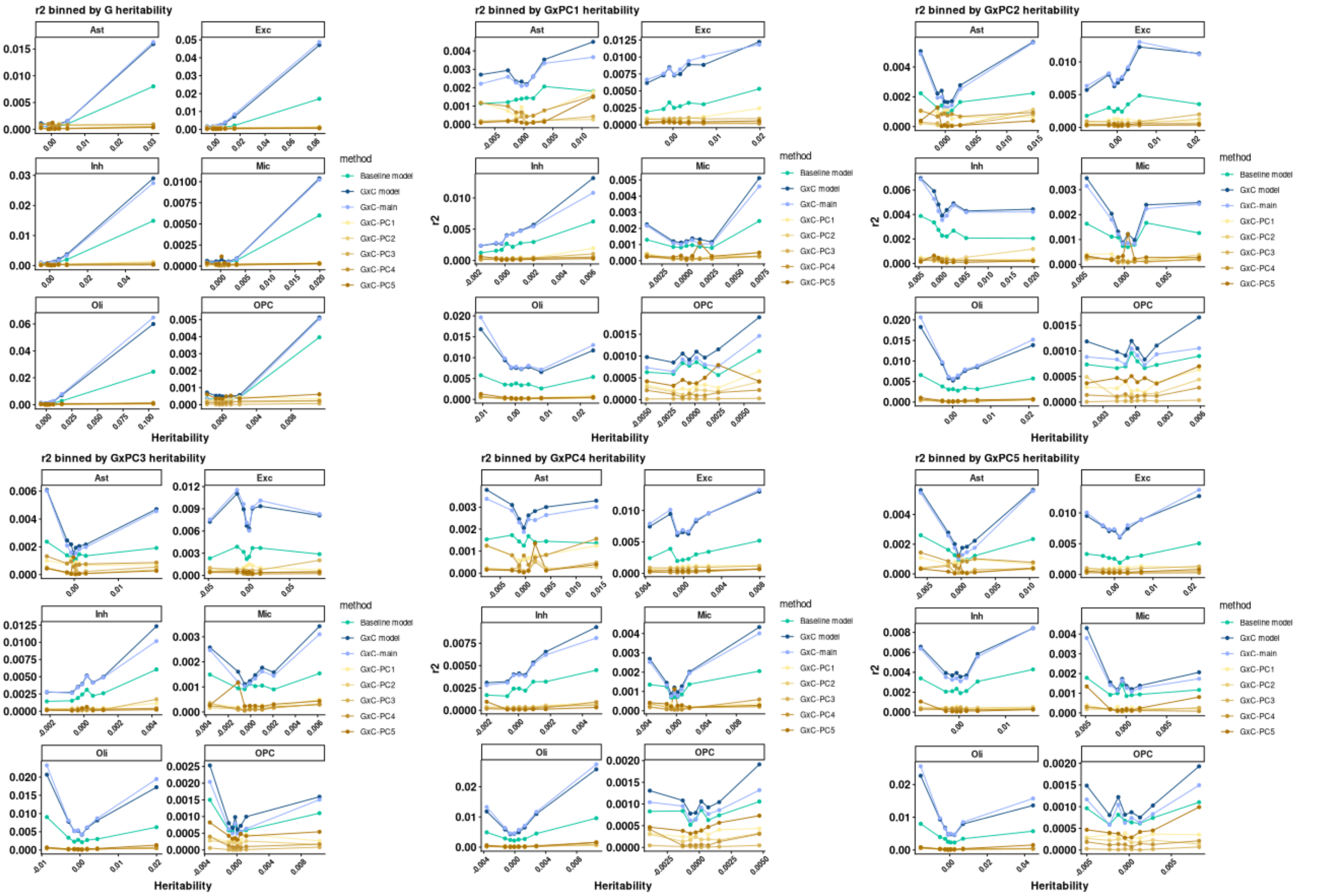


**Supplementary Fig. 32 | Cross-validated prediction accuracy by heritability.** Mean cross-validated prediction accuracy ($r^2$; y axis) for genes binned by scHINT heritability (x axis; bin mean heritability) of the main genetic component (top left) or of G×PC1 to G×PC5 (other panels), shown for each cell type. Lines show the baseline model, the combined G×Cell-State model (G×C model), its main-effect weights (G×C-main) and the interaction weights for each PC (G×C-PC1–PC5). Prediction accuracy of the corresponding component increases with its heritability.

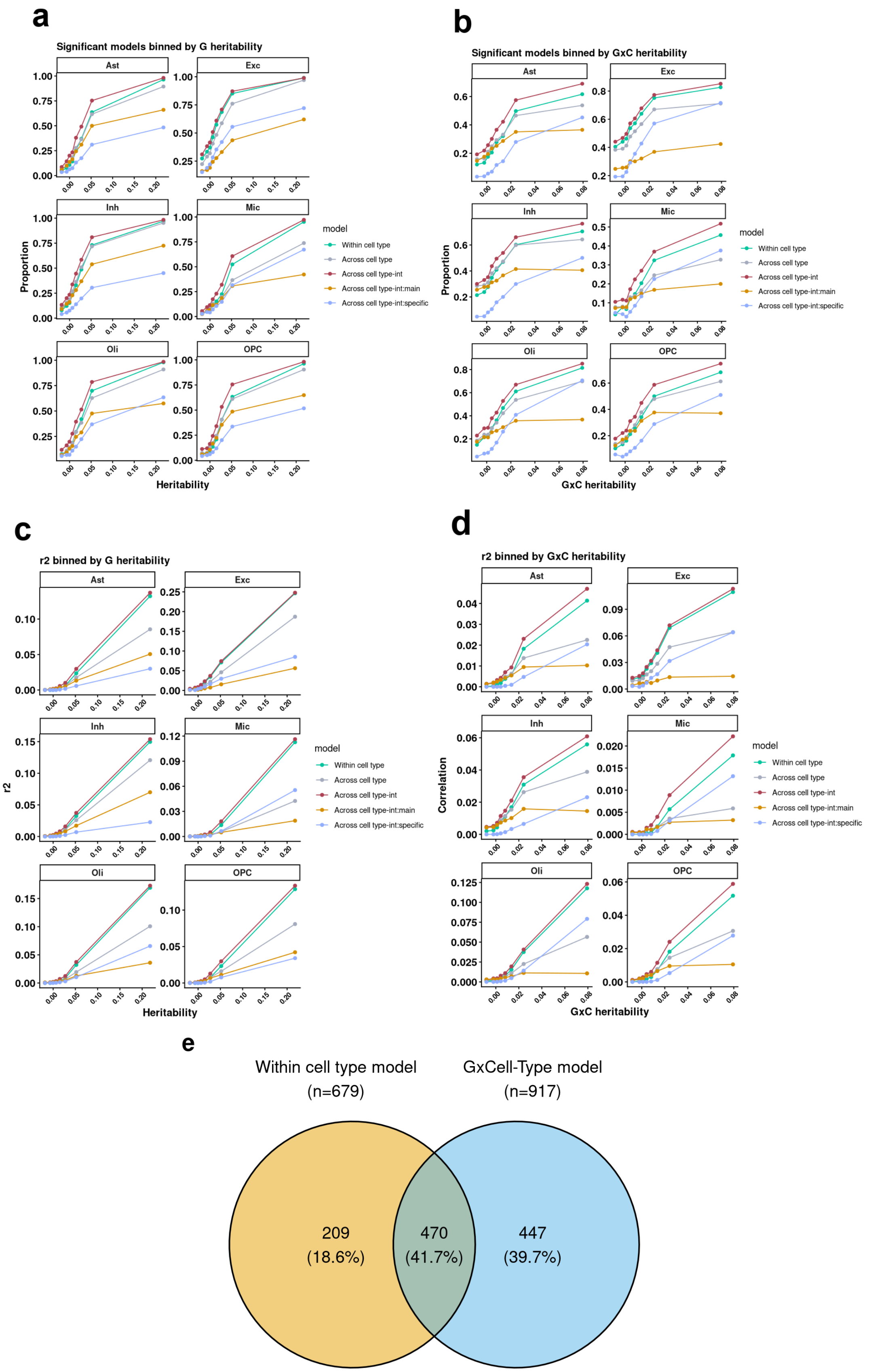


**Supplementary Fig. 33 | Prediction performance and gene–trait associations of G×Cell-Type TWAS models. a**,**b**, Proportion of gene–cell-type models with significant out-of-sample prediction for genes binned by scHINT main genetic (G) heritability (**a**) or G×Cell-Type heritability (**b**; x axis, bin mean heritability), shown separately for each cell type (Ast,

Exc, Inh, Mic, Oli, OPC). **c**,**d**, Cross-validated prediction accuracy ($r^2$) for the same heritability bins of G (**c**) and G×Cell-Type (**d**). In **a–d**, lines show the cell-type-specific model trained within each cell type (Within cell type), the pooled main-effect model trained across cell types (Across cell type), the full G×Cell-Type interaction model (Across cell type int), and its shared main genetic component (int-main) and cell-type-specific interaction component (int-specific). Prediction performance increases with heritability for all models; models that use information across cell types generally outperform within-cell-type models, and the gain is largest for the cell-type-specific interaction component in genes with high G×Cell-Type heritability, whereas the shared main component improves little. **e**, Overlap of significant gene–trait associations from transcriptome-wide association analyses of seven brain-related traits (ADHD, bipolar disorder, major depressive disorder, schizophrenia, ALS, Alzheimer's disease and Parkinson's disease) using conventional within-cell-type (pseudo-bulk) prediction models (n = 679) and G×Cell-Type models (n = 917). Numbers are association counts (% of the union); 447 associations (39.7%) are identified only by the G×Cell-Type model.

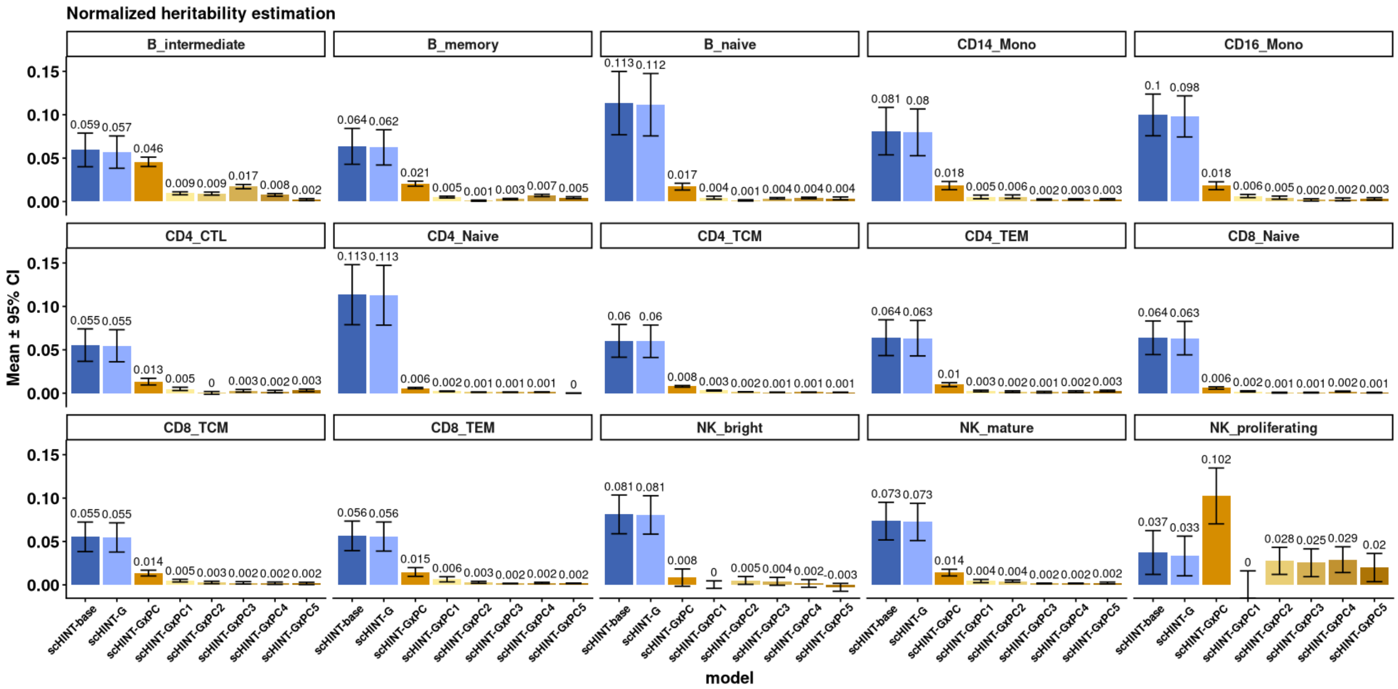


**Supplementary Fig. 34 | G×Cell-State heritability across 15 blood cell types in OneK1K.** Normalized heritability estimated by scHINT in 15 cell types from OneK1K scRNA-seq data (B, CD4 T, CD8 T, NK and monocyte lineages; panels). Bars show scHINT-base, scHINT-G, scHINT-G×PC (sum over components) and individual G×PC1–G×PC5 components (mean ± 95% CI; numbers are means). G×Cell-State heritability is lower than in brain and is relatively larger in differentiated or proliferating populations (for example proliferating NK cells) than in naive populations.

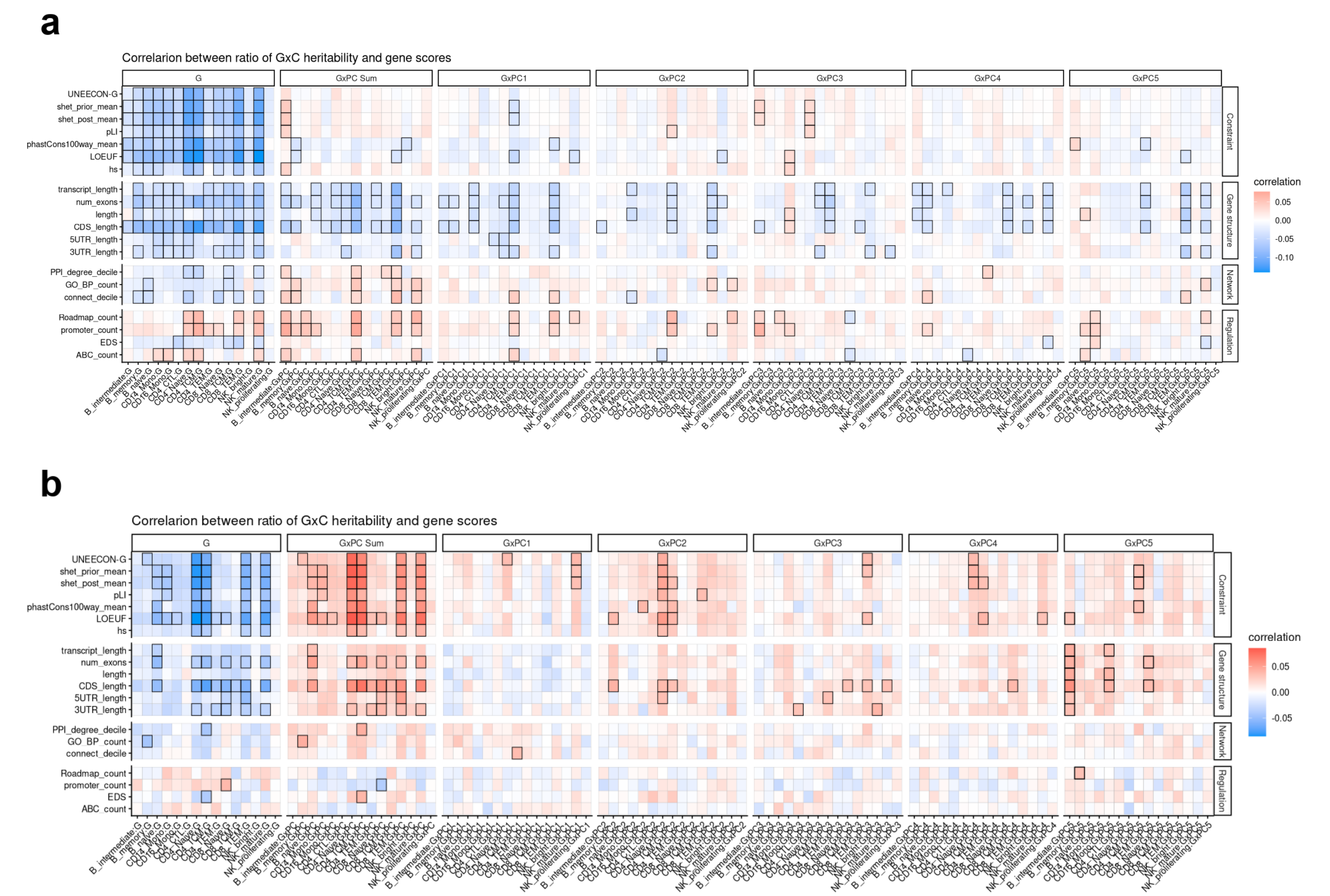


**Supplementary Fig. 35 | Associations of G×Cell-State heritability with gene-level annotations in blood cell types.** Correlation (standardized regression coefficient) between heritability components and gene-level scores across the 15 OneK1K cell types. **a**, Absolute heritability. **b**, Proportion of genetic heritability attributable to each component. Rows are gene scores grouped into constraint, gene structure, network and regulation annotations; columns are the main genetic component (G), summed G×PC (GxPC Sum) and G×PC1–G×PC5 for each cell type. Colour indicates the correlation (red, positive; blue, negative) and black borders mark significant correlations.

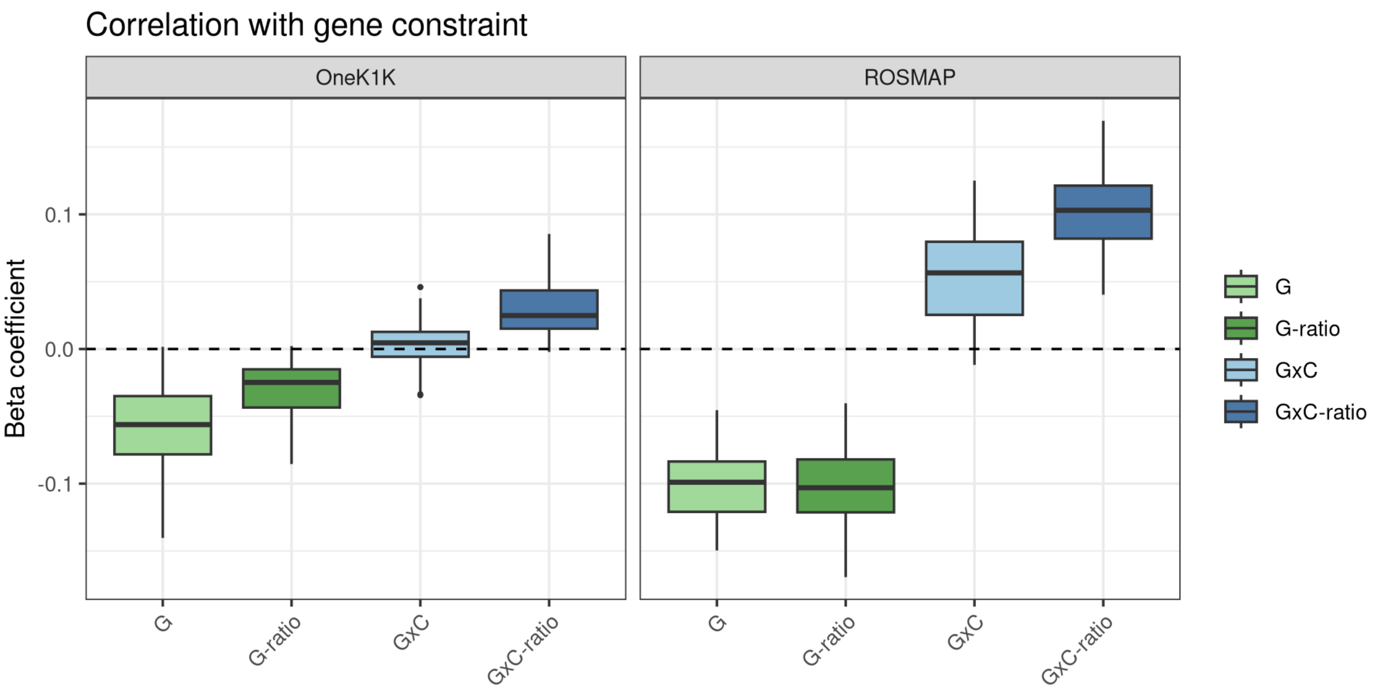


**Supplementary Fig. 36 | Association of heritability components with gene constraint in blood and brain.** Distribution of standardized regression coefficients between gene constraint scores and main genetic heritability (G), the proportion of heritability attributable to G (G-ratio), G×Cell-State heritability (G×C) and the proportion attributable to G×Cell-State (G×C-ratio) in OneK1K (left; 15 cell types) and ROS/MAP (right; 6 cell types). Each box summarizes coefficients across cell types and the seven constraint scores (centre line, median; box, interquartile range; whiskers, 1.5 × interquartile range; points, outliers). The dashed line marks zero. The proportion attributable to G×Cell-State is positively associated with constraint in both tissues, but absolute G×Cell-State heritability is only weakly associated in blood.

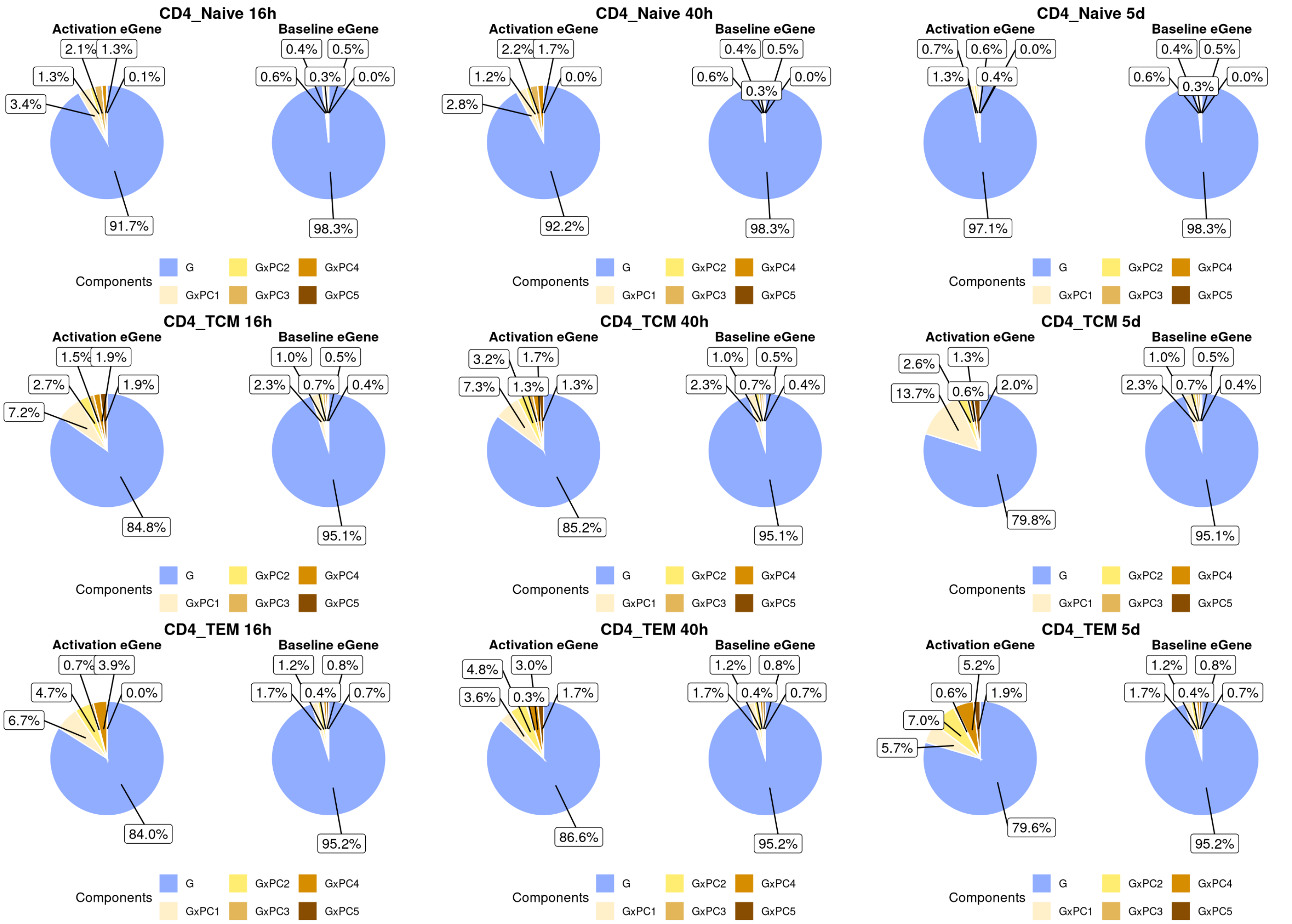


**Supplementary Fig. 37 | G×Cell-State fraction in CD4 T-cell activation-dependent eGenes.** Relative contribution (%) of heritability components (main genetic, G; G×PC1–G×PC5) estimated by scHINT in OneK1K for eGenes specific to CD4 T-cell activation (Activation eGene) and for baseline eGenes (Baseline eGene) from Soskic et al., in naive, central memory (TCM) and effector memory (TEM) CD4 T cells (rows) at 16 h, 40 h and 5 days after activation (columns). The G×Cell-State fraction is larger for activation-dependent eGenes, particularly in memory cells.

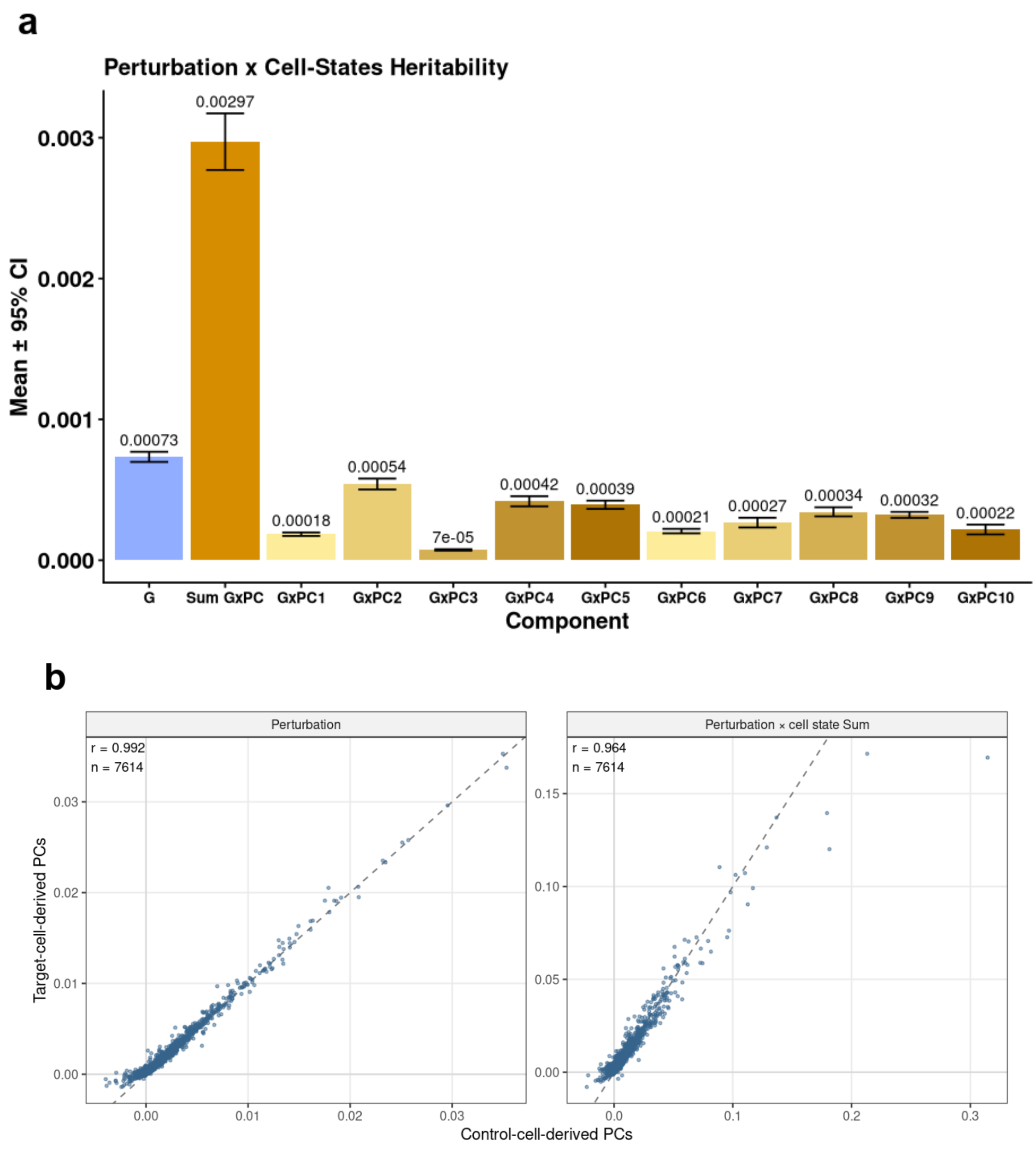


**Supplementary Fig. 38 | Robustness of perturbation heritability estimates to the cell-state PC definition. a.** Perturbation × cell-state heritability in K562 Perturb-seq, using PCs derived from control cells. Mean variance in response-gene expression explained by the main perturbation effect (G), the summed perturbation × cell-state component (Sum GxPC) and each of the first ten perturbation × cell-state PC components (GxPC1–10). PCs were derived from non-targeted control cells and projected onto targeted cells. Bars show the mean across response genes, and error bars show 95% confidence intervals. b. Each point is one response gene. The x axis shows the estimate using PCs derived from control (non-targeted) cells and projected onto targeted cells (as in the main analysis). The y axis shows the estimate using PCs derived from the targeted cells. Left, main perturbation component. Right, summed perturbation × cell-state component. Dashed lines indicate y = x, and r is the Pearson correlation.

**Supplementary notes**

1. **G×Cell-Type heritability model**

**1.1 Generative model**

For each gene, let $y_{it}$ denote the normalized pseudo-bulk expression level for individual i = 1,…,n in cell type t = 1,…,T. Let $g_i \in \mathbb{R}^p$ denote the vector of standardized cis-genotypes for individual i, defined using variants within ±500 kb of the gene. We model pseudo-bulk expression across cell types as:

$$y_{it} = g_i^{\mathsf{T}}\beta + g_i^{\mathsf{T}}\gamma_t + \alpha_t + o_{it}^{\mathsf{T}}\eta + e_i + \varepsilon_{it}$$

Here, $\beta \in \mathbb{R}^p$ represents the genetic effect shared across cell types, $\gamma_t \in \mathbb{R}^p$ represents the cell-type-specific genetic deviation for cell type t, $\alpha_t$ captures the mean effect of cell type, and $o_{it}$ denotes additional sample-level covariates. The individual-level random effect $e_i$ captures expression variation shared across pseudo-bulk samples from the same donor, and $\varepsilon_{it}$ denotes residual sample-level variation. We assume

$$\beta \sim N\left(0, \sigma_G^2 p^{-1} I_p\right), \quad \gamma_t \sim N\left(0, \sigma_{G\times CT}^2, p^{-1} I_p\right)$$

$$\alpha_t \sim N(0, \sigma_T^2), e_i \sim N(0, \sigma_I^2), \quad \varepsilon_{it} \sim N(0, \sigma_\varepsilon^2)$$

with mutually independent random components. Under this model, $\sigma_G^2$ quantifies the genetic variance shared across cell types, whereas $\sigma_{G\times CT}^2$, quantifies cell-type-specific genetic variance (Supplementary Fig. 1).

**1.2 Haseman–Elston estimation**

scHINT estimates these variance components by extending Haseman–Elston regression to pseudo-bulk expression measured across multiple cell types. Let

$$K_{ij}^G = \frac{g_i^{\mathsf{T}} g_j}{p}$$

denote the local cis-genetic relatedness between individuals i and j in cell type s and t correspondingly. For two pseudo-bulk observations (i,s) and (j,t), the expected expression product is

$$\mathbb{E}\left(y_{is}y_{jt}\right) = \sigma_G^2 K_{ij}^G + \sigma_{G\times CT}^2 K_{ij}^G \mathbf{1}(s = t) + \sigma_{CT}^2 \mathbf{1}(s = t) + \sigma_I^2 \mathbf{1}(i = j)$$

For distinct observations, the residual term is zero and is not fitted as an off-diagonal Haseman–Elston component. The fitted kernels therefore include local genetic relatedness $K_{ij}^G$ cell-type indicator kernels $\mathbf{1}(s = t)$, their genetic interaction kernels $K_{ij}^G \mathbf{1}(s = t)$ and the individual-level kernel $\mathbf{1}(i = j)$. Covariate kernels may also be included when the corresponding covariates are modeled as random effects. Haseman–Elston regression estimates the variance components by regressing cross-products of distinct pseudo-bulk observations on these kernels.

2. **The definition of cell-level heritability.**

We first consider a simple scenario with no G×C effects, in which each sample contains m cells. Given individual i, cell k, the cell level expression follows:

$$q_{ik} = \alpha_1 X_i + \alpha_2 Z_{ik} + \beta\, G_i + I_i + \varepsilon_{ik}$$

where $X_i$ is an standardized sample-level covariate, $Z_{ik}$ is a standardized cell-level covariate or cell-state factor, $G_i$ represents the standardized genotype shared by all cells from individual i, $I_i$ is an additional individual-level random effect with variance equals $\sigma_I^2$, and $\varepsilon_{ik}$ is cell-specific residual variation with variance equals $\sigma_\varepsilon^2$. This model therefore contains both components shared across cells from the same individual and components that vary among cells.

At the cell level, the raw heritability is defined as the proportion of total variation across individual cells attributable to genetic variation:

$$h_{cell}^2 = \frac{var(\beta\, G_i)}{var(q_{ik})} = \frac{\beta^2}{\alpha_1^2 + \alpha_2^2 + \beta^2 + \sigma_I^2 + \sigma_\varepsilon^2}$$

The denominator includes all sources of cell-level variation: individual-level covariates, cell-state variation, genetic effects, individual-specific effects, and cell-specific residual noise. Thus, cell-level heritability measures the relative genetic contribution to variation among cells.

Pseudo-bulk expression is obtained by aggregating the m cells from each individual:

$$Q_i = \sum_{k=1}^{m} (\alpha_1 X_i + \alpha_2 Z_{ik} + \beta\, G_i + I_i + \varepsilon_{ik}) = m\,(\alpha_1 X_i + \beta\, G_i + I_i) + \sum_{k=1}^{m} (\alpha_2 Z_{ik} + \varepsilon_{ik})$$

Equivalently, the individual-level mean expression is

$$\frac{Q_i}{m} = \alpha_1 X_i + \beta\, G_i + I_i + \frac{\sum_{k=1}^{m} (\alpha_2 Z_{ik} + \varepsilon_{ik})}{m}$$

Averaging does not reduce components shared across cells, including the genetic effect and individual-level effect. In contrast, cell-specific variation is reduced. Assuming independence across cells, pseudo-bulk heritability is

$$h_{PB}^2 = \frac{var(\beta\, G_i)}{var(\frac{q_i}{m})} = \frac{\beta^2}{\alpha_1^2 + \beta^2 + \sigma_I^2 + \frac{\alpha_2^2 + \sigma_\varepsilon^2}{m}}$$

Therefore, the two heritability definitions have different denominators. Cell-level heritability includes the full cell-state and residual variances, whereas pseudo-bulk heritability includes only the remaining variation after these cell-specific components have been reduced by averaging. Pseudo-bulk heritability may consequently be larger even when the underlying genetic variance is identical.

The two estimates are therefore difficult to compare directly. We choose two normalization strategies by excluding $\sigma_\varepsilon^2$ or both $\sigma_\varepsilon^2$ and $\alpha_2^2$ from the denominator

$$h_{cell_{lower}}^2 = \frac{\beta^2}{\alpha_1^2 + \alpha_2^2 + \beta^2 + \sigma_I^2}$$

$$h_{cell_{upper}}^2 = \frac{\beta^2}{\alpha_1^2 + \beta^2 + \sigma_I^2}$$

We will have an upper and lower bound of cell level heritability and $h_{PB}^2$ will fall between the intervals.

$$h_{cell_{lower}}^2 < h_{PB}^2 < h_{cell_{upper}}^2$$

We then consider a model with G×C effect

$$q_{ik} = \alpha_1 X_i + \alpha_2 Z_{ik} + \beta\, G_i + \gamma\, G_i Z_{ik} + I_i + \varepsilon_{ik}$$

Similarly, the raw G×C heritability is defined as the proportion of total variation across individual cells attributable to G×C variation:

$$h_{GxC-cell}^2 = \frac{var(\gamma\, G_i Z_{ik})}{var(q_{ik})} = \frac{\gamma^2}{\alpha_1^2 + \alpha_2^2 + \beta^2 + \gamma^2 + \sigma_I^2 + \sigma_\varepsilon^2}$$

We also choose two normalization strategies by excluding $\sigma_\varepsilon^2$ or both $\sigma_\varepsilon^2$ and $\alpha_2^2$ from the denominator

$$h_{G-cell_{lower}}^2 = \frac{\beta^2}{\alpha_1^2 + \alpha_2^2 + \beta^2 + \gamma^2 + \sigma_I^2}$$

$$h_{G-cell_{upper}}^2 = \frac{\beta^2}{\alpha_1^2 + \beta^2 + \gamma^2 + \sigma_I^2}$$

$$h_{GxC-cell_{lower}}^2 = \frac{\gamma^2}{\alpha_1^2 + \alpha_2^2 + \beta^2 + \gamma^2 + \sigma_I^2}$$

$$h^2_{GxC-cell_{upper}} = \frac{\gamma^2}{\alpha_1^2 + \beta^2 + \gamma^2 + \sigma_I^2}$$

### 3. Extended simulation of G×Cell-State model

In supplementary notes, we further discuss how other parameters influence the heritability estimation and perform additional simulations to examine how cell type abundance, heritable PCs and standardization procedures influence the estimation.

#### 3.1 Standardization methods

We compared inverse-normal transformation and standard normalization under simulations with and without G×C effects (**Supplementary Fig. 5**). Under the null setting, where both genetic and G×C variance components were absent, baseline mode, scHINT-G and scHINT-G×C were well calibrated, with estimates remaining close to zero across sample sizes. For scHINT, standard normalization generally provided more accurate estimation than inverse-normal transformation. Estimates of the main genetic component were closer to the simulated values regardless of whether the G×C component was absent or present. Standard normalization also improved estimation of the G×C variance component, producing estimates that were consistently closer to the truth across cell counts. In contrast, inverse-normal transformation introduced modest downward bias, particularly for the G×C component. We therefore used standard normalization in real data. Like previous result, the baseline method consistently overestimated the main genetic variance when G×C effects were present.

#### 3.2 Heritable PCs

We next evaluated scHINT under a setting in which the cell-state PCs were themselves partially genetically determined. Specifically, we simulated PCs with 10% of their variance explained by genotype and considered gene-expression models with either a nonzero G×C component or a nonzero marginal PC component. Across increasing cell counts, scHINT produced results consistent with those observed in the main simulations (**Supplementary Fig. 6 a**). The estimated main genetic, G×C variance components remained close to their simulated values, with no substantial bias introduced by the heritability of the PCs. These results indicate that scHINT is robust when the inferred cell-state features are themselves partially heritable.

We then assessed the heritability of the first five PCs in the real single-cell data (**Supplementary Fig. 6 b**). Across all major cell types, the estimated PCs' heritability was generally close to zero, with only a small number of PCs showing modest positive or negative estimates. Even the largest estimates were below approximately 0.5%, indicating that SNPs explain only a very small proportion of variation in these cell-state axes. Together with the simulation results, this suggests that the modest heritability of the observed PCs is unlikely to substantially bias scHINT estimates of main genetic or G×C effects.

#### 3.3 Cell type abundance

We further considered a setting in which cell-type abundance was genetically influenced. Because the cell-level simulations analyze each cell type separately and estimate heritability within cell type, we modeled this scenario by allowing the number of cells contributed by each individual to depend on genotype. Specifically, number of cell counts were simulated as a function of four causal variants. scHINT produced unbiased heritability estimates in both the absence and presence of G×C effects (**Supplementary Fig. 7**). Estimates remained close to their simulated values whether or not cell counts were included as covariates, indicating that genetically driven variation in the number of sampled cells does not substantially bias within-cell-type estimation. We revisit the related issue of heritable cell-type proportions in the G×Cell-Type simulations, where variation in relative cell-type abundance is modeled directly.

### 4. Simulation of G×Cell-Type model

#### 4.1 Simulate pseudo-bulk level gene expression data in two cell types

We simulated pseudo-bulk level gene expression under a G×Cell-Type genetic architecture using real genotype data to preserve the local allele-frequency and linkage-disequilibrium structure. Similar to simulate cell-level gene expression, simulations were based on real genotype data to preserve local allele-frequency and linkage-disequilibrium structure. For each simulated gene, we selected cis-variants located within 500 kb of the gene body and standardized the resulting genotype matrix. For each simulation setting, parameters were taken from a predefined grid, including

- the main genetic variance component $h_G^2 = (0, 0.05, 0.1)$,
- the G×Cell-Type variance component $h_{G\times CT}^2 = (0, 0.05, 0.1)$,
- the individual-level variance component $h_I^2 = (0, 0.1)$,
- the cell-type variance component $h_{CT}^2$, = (0,0.1),
- the number of causal variants (n_causal = 4).

We first simulated a main genetic effect shared across cell types. Specifically, if X denotes the standardized N × P genotype matrix, the shared genetic effect was simulated as Xβ, where nonzero elements of βwere sampled from N(0, $h_G^2$/n_causal). This genetic effect was the same across the two simulated cell types so that the same main cis-genetic effect contributed to expression in both cell types. We then generated a cell-type-dependent genetic component. For the same causal variants, we sampled two independent sets of cell-type-specific effects, $\beta_{CT1}$ and $\beta_{CT2}$, with nonzero elements sampled from N(0, $h_{G\times C}^2$/n_causal). The G×Cell-Type genetic effect was defined as X$\beta_{CT1}$ for the first cell type and X$\beta_{CT2}$ for the second cell type. This component was rescaled so that its empirical variance matched the target ratio between the G×Cell-Type and main genetic components. The total genetic value was therefore the sum of the shared genetic effect and the cell-type-dependent genetic effect.

Formally, for individual i in cell type t, expression was simulated as

$$y_{it} = X_i\beta + X_i\beta_{CT_t} + u_i + a_t + e_{it}$$

where $X_i\beta$ is the main genetic effect shared across cell types, $X_i\beta_{CT_t}$ is the G×cell-type effect, $u_i$ is an individual-level random effect shared by the same individual across cell types, $a_t$ is a cell-type-specific intercept, and $e_{it}$is residual noise. After constructing the genetic value, the remaining non-genetic variance was computed from the empirical genetic variance so that the simulated expression had the desired genetic-to-non-genetic variance ratio. Individual-level random effects, cell-type effects, and residual errors were generated from normal distributions and scaled accordingly.

#### 4.2 Simulation results of G×Cell-Type interaction model

We compared three models with increasing levels of variance-component adjustment. Model 0 included only the main genetic component; Model 1 included both main genetic and G×Cell-Type components; and Model 2 additionally included marginal cell-type and individual-level variance components. We considered a range of main genetic and G×Cell-Type heritability, with $h_G^2 = h_{G\times C}^2$ varying from 0.05 to 0.20, under settings in which the cell-type and individual-level components were either absent or nonzero (0.1).

The top panel compares estimates of the main genetic heritability. Model 0 increasingly overestimated $h_G^2$ as the magnitude of the unmodeled G×Cell-Type component increased, indicating that interaction effects were partially absorbed into the main genetic component. Model 1 produced accurate estimates when both the marginal cell-type and individual-level components were absent, but became inflated when either component was present. In contrast, Model 2 consistently recovered the simulated main genetic heritability across all settings.

The bottom panel compares estimates of G×Cell-Type heritability from Models 1 and 2. Model 1 was unbiased when the marginal cell-type and individual-level variance components were both zero, but systematically underestimated $h_{G\times C}^2$when these additional sources of variation were present. Model 2 remained well calibrated across all simulation settings. Together, these results demonstrate that jointly modelling main genetic, G×Cell-Type, marginal cell-type and individual-level effects, is necessary for unbiased estimation when multiple sources of cell-level variation are present.

#### 4.3 Cell type abundance

Similar to the preceding cell-level simulations, we further considered a setting in which cell-type abundance was genetically influenced. We simulated two cell types and allowed genotype to increase the abundance of one cell type relative to the other, thereby inducing genetically driven variation in cell-type proportions. We varied both the main genetic heritability and the G×Cell-Type heritability across simulation settings.

Without adjustment for cell-type proportions, the baseline model consistently overestimated the main genetic heritability when a nonzero G×Cell-Type component was present. Under the same settings, scHINT tended to underestimate the main genetic component and overestimate the G×Cell-Type component, indicating that genetically driven differences in cell-type composition can be partially absorbed into the estimated interaction effect. After including cell-type proportions as covariates, estimates of both the scHINT-G and scHINT-G×Cell-Type became substantially closer to the true values. These results indicate that scHINT remains robust to genetically influenced cell-type composition when cell-type proportions are appropriately modeled, and they highlight the importance of adjusting for cell-type abundance in G×Cell-Type analyses.

### 5. Comparison with scGeneHE in real data

Direct application of scGeneHE to the full ROS/MAP data was computationally prohibitive: in our benchmarking, analyzing a single gene with approximately one million cells required about one hour. Thus, genome-wide analysis across all genes and six cell types was not feasible. The original scGeneHE preprint analyzed OneK1K, which contains substantially fewer cells per cell type, and focused on several hundred highly expressed genes. To enable a comparable analysis in ROS/MAP, we therefore performed meta-cell aggregation, generating approximately 50,000 meta-cells within each cell type.

Using these meta-cell expression profiles, we recomputed within-cell-type states using PCA and applied both scGeneHE and scHINT to the same meta-cell data. We observed that scGeneHE estimates were highly sensitive to covariate adjustment. Specifically, we evaluated an unadjusted model and an adjusted model that additionally included 25 pseudo-bulk PCs and five cell-level PCs as covariates. These two specifications produced substantially different heritability estimates across cell types. The unadjusted scGeneHE estimates were generally closer to the main genetic component estimated by scHINT-G, whereas covariate-adjusted scGeneHE produced markedly different estimates. It was consistent with our simulation results that scGeneHE is inflated when there existed significant non-zero variance from those covariates.

### 6. The robustness of G×Cell-State estimation across different settings

#### 6.1 PC methods

The G×Cell-State heritability estimates were robust to the approach used for state inference and cell sampling. First, we used other three alternative approaches to infer cell states: across-cell-type PCA, residual PCA[1] and consensus non-negative matrix factorization (cNMF[2]) (**Methods**). We compared G×Cell-State heritability estimates obtained using other definitions of cell state methods. Main genetic heritability was broadly consistent across the four cell-state representations included in the model. In contrast, G×Cell-State estimates varied by cell-state definition. Overall, within-cell-type PCA yielded the higher G×Cell-State heritability across cell types than the other three methods. Across-cell-type PCA produced unusually negative G×Cell-State estimates for several PCs in excitatory neurons. Because across-cell-type PCA is performed jointly across all cell types, its leading PCs may capture cell-type differences rather than within-cell-type state variation. This can induce correlations between inferred cell states and cell-type identity, potentially leading to spurious or negative G×Cell-State signals. We also examined correlations among cell-state components within each cell type. By construction, within-cell-type PCs are orthogonal to one another, whereas cell-state components derived from the other three methods showed significant cross-component correlations, suggesting that they may capture partially redundant biological variation. We therefore focused analyses on cell states inferred using within-cell-type PCA (**Supplementary Fig. 14**).

#### 6.2 The number of pseudo-bulk level PCs included

To determine the number of pseudo-bulk principal components (PCs) to include as covariates, we evaluated the sensitivity of pseudo-bulk heritability estimates to this choice. We varied the number of included pseudo-bulk PCs from 5 to 50 and re-estimated heritability separately for each of the six major brain cell types. The estimates were highly stable across the full range of PC numbers, with essentially no systematic change as additional PCs were included (**Supplementary Fig. 15**). This indicates that the estimated genetic variance is robust to reasonable variation in the number of pseudo-bulk PCs. Based on this stability, we included 25 pseudo-bulk PCs in all subsequent analyses which consistent with previous studies[3,4].

#### 6.3 The number of cell level PCs included

We evaluated the sensitivity of variance-component estimates to the number of cell-level PCs included as covariates (interaction terms fixed at 5, **Supplementary Fig. 16**). We varied the number of included PCs from 5 to 45 and quantified variance attributable to the main genetic effect, the summed G×PC1–PC5 interaction effects, 25 pseudo-bulk PCs, the individual-level component, and the included cell-level PCs. As expected, the total variance attributed to cell-level PCs increased with the number of PCs included. In parallel, variance attributed to pseudo-bulk PCs and the individual-level component generally decreased, indicating that additional cell-level PCs absorbed residual expression heterogeneity that would otherwise be captured by these nuisance components. In contrast, the absolute variance explained by the main genetic effect remained stable across the number of included PCs in all six cell types, whereas that explained by G×PC1–PC5 declined gradually in most cell types (excitatory neurons being the exception) but changed far less than the pseudo-bulk PC and individual-level components.

We also examined two normalized versions of cell-level heritability, which exclude cell-specific residual variance (lower bound) or both residual and cell-state variance (upper bound) from the denominator, as the number of cell-level PCs increased from 5 to 45 (**Supplementary Fig. 17**). The lower bound decreased steadily with more PCs for both scHINT-G and scHINT-G×C in every cell type; for example, scHINT-G in astrocytes fell from 0.028 to 0.018 and scHINT-G×C in oligodendrocytes from 0.039 to 0.006, because the cell-state variance carried by the added PCs enlarges the denominator. In contrast, the upper bound for scHINT-G increased with the number of PCs in most cell types (for example, from 0.077 to 0.186 in astrocytes and from 0.181 to 0.442 in oligodendrocytes), and was stable in excitatory neurons. The upper bound for scHINT-G×C rose and then plateaued in astrocytes, excitatory neurons, inhibitory neurons and microglia, and declined in oligodendrocytes and OPCs.

#### 6.4 The number of interaction terms included

To choose the number of G×PC interaction terms, we varied the number of cell-state PCs in the interaction component (5, 10, 15 and 20) and compared two outputs across cell types (**Supplementary Fig. 18a**). The main genetic variance component (G) was stable across all four settings. The summed G×PC variance component increased with the number of PCs in astrocytes, excitatory neurons, inhibitory neurons and microglia, whereas in oligodendrocytes and OPCs it increased up to about 10–15 PCs and then plateaued. To test whether the additional interaction terms carry biologically meaningful signal, we correlated the summed G×PC heritability with seven gene constraint scores (**Supplementary Fig. 18b,c**). Averaged across cell types, the correlation was strongest with five PCs (0.04–0.07 across scores), remained positive for all scores with 10 PCs (0.01–0.04), and approached zero with 15 and 20 PCs. The cell-type-specific results followed the same overall trend with some heterogeneity. In astrocytes, oligodendrocytes and microglia the correlation declined as more PCs were added, and in OPCs it turned negative with 15–20 PCs, whereas in excitatory and inhibitory neurons it remained positive up to 20 PCs. The association with constraint is therefore robust to moderate changes in the number of PCs, whereas higher-order PCs mainly add noise in most cell types. When we down-sampled excitatory and inhibitory neurons to 10% of cells, the correlation stayed positive but declined steadily as more PCs were included (**Supplementary Fig. 18d**). With all cells, additional PCs captured more non-technical signal, whereas with fewer cells the first five PCs maximized the signal, suggesting that the optimal number of PCs depends on cell number. We therefore used the first five PCs for all cell types in all downstream G×PC analyses, noting that the optimal number may vary by cell type and read depth.

#### 6.5 The genes used to derive the cell-state PCs

The cell-state PCs are derived from the same expression data in which heritability is estimated, so a gene could contribute to the PCs that define its own interaction term, which could inflate G×PC heritability. To test this, we re-derived the within-cell-type PCs for each chromosome using only genes located outside that chromosome (leave-one-chromosome-out, LOCO PCA), and used these PCs to estimate heritability for genes on the held-out chromosome. We compared the resulting estimates of main genetic (G) and summed G×PC heritability with those from PCs derived from all genes (**Supplementary Fig. 19**). Mean estimates across genes were highly similar between the two approaches in every cell type for both G and G×PC sum. At the gene level, G estimates were essentially identical ($r = 1.000$) and summed G×PC estimates were highly concordant ($r = 0.962$). These results indicate that our G×PC heritability estimates are not driven by the focal gene's contribution to the cell-state axes.

#### 6.6 Sparsity of gene expression

We next evaluated the robustness of scHINT to sparsity in single-cell expression profiles using two complementary strategies. First, we artificially increased sparsity by randomly setting an increasing proportion of cell-level expression measurements to zero. For each sparsity level, we recalculated the cell-state PCs from the perturbed expression matrix and re-estimated the main genetic (G) and G×Cell-State components. Across a broad range of zero proportions, both components remained generally stable, indicating that scHINT is robust to moderate levels of randomly introduced sparsity. Only under very high sparsity did we observe noticeable deviation, characterized primarily by inflation of the G×Cell-State component and a modest reduction in the main genetic component (**Supplementary Fig. 20**).

Second, we assessed robustness to variation in expression depth by stratifying cells according to library-size quantiles and repeating the analysis using cells from different expression-depth ranges. The estimated variance components again remained broadly consistent across strata. Cells with lower library size showed a modestly smaller main genetic component and a slightly larger G×Cell-State component, whereas estimates from higher-depth cells were similar to the overall results (**Supplementary Fig. 20**). Together, these two analyses suggest that scHINT is generally robust to both experimentally induced sparsity and natural variation in cell-level sequencing depth.

#### 6.7 Interaction with disease status

To test whether disease status affects our estimates, we classified ROS/MAP donors as AD if they had a clinical diagnosis of AD at death (cogdx 4 or 5) and pathological AD (Braak stage $\geqslant 4$ and CERAD score $\leqslant 2$), and as non-AD otherwise. We added AD status as a categorical context variable and fitted a pooled G×AD interaction component, which captures extra genetic covariance between donors in the same AD group, with AD status also included as a main effect. We fitted two models in each cell type: a model with the G×AD term only (no G×PC terms), and the full model with G×PC1–5 and G×AD (**Supplementary Fig. 21**). The G×AD component was close to zero in both models and in all cell types, and including AD status did not change the main genetic (G) or summed G×PC heritability relative to our original model. We therefore found no evidence that AD status affects our heritability estimates and did not observe significant G×AD interaction heritability.

### 7. G×Cell-Type heritability results

#### 7.1 Two cell types of interaction model

To further evaluate cell-type-dependent genetic effects, we performed pairwise analyses across the six major cell types. For each pair, we fitted three models with increasing adjustment for non-genetic variation: HEreg-base, including only the main genetic component (G); HEreg-G×CT, jointly modeling G and G×CT; and scHINT, additionally accounting for marginal cell-type (CT) and individual-level effects. HEreg-base generally produced higher genetic heritability estimates than scHINT, which models G×CT together with cell-type and individual-level effects, indicating that unmodeled cell-type-dependent genetic effects can be absorbed into the main genetic component (**Supplementary Fig. 22**). However, simply adding G×CT was insufficient: HEreg-G×CT frequently yielded inflated G estimates together with deflated, and sometimes negative, G×CT estimates. In contrast, after accounting for CT and individual-level variance, scHINT produced consistently positive and more stable G×CT estimates, highlighting the importance of modeling these additional variance components for appropriately partitioning shared and cell-type-dependent genetic effects.

G×CT heritability varied substantially across cell-type pairs and appeared to reflect their biological similarity (**Supplementary Fig. 23**). The excitatory–inhibitory neuron pair showed the lowest G×CT heritability (~0.006), whereas comparisons between more distinct cell types, such as neurons and microglia, oligodendrocytes, or OPCs, generally showed larger G×CT components (~0.016–0.018). To account for differences in the overall magnitude of genetic regulation across cell types, we normalized G×CT heritability by the main genetic heritability of the corresponding cell-type pair and compared it with gene-expression similarity. Normalized G×CT heritability showed a clear inverse relationship with gene-level expression correlation: transcriptionally similar cell-type pairs had weaker relative G×CT effects, whereas more divergent cell types showed stronger effects (**Supplementary Fig. 24**). Consistently, the fitted regression line decreased with increasing expression correlation, with excitatory and inhibitory neurons representing the high-similarity, low-G×CT extreme. Together, these results suggest that cell-type-dependent genetic regulation becomes progressively stronger as cell types become more transcriptionally distinct.

We also examined whether G×CT heritability was related to differences in gene expression between cell types. Genes were stratified according to whether their expression differed significantly between the two cell types and whether they had high or low overall expression levels (**Supplementary Fig. 25**). Among highly expressed genes, genes with significant expression differences showed substantially higher G×CT heritability than genes without significant differences (0.035 vs. 0.018). In contrast, this pattern was weaker among lowly expressed genes (0.016 vs. 0.011). These results suggest that G×CT effects are particularly pronounced for highly expressed genes whose expression levels differ between cell types, further supporting a relationship between transcriptional divergence and cell-type-dependent genetic regulation.

#### 7.2 Six cell types of interaction model

Alternatively, we extended the pairwise analysis to jointly model genetic heterogeneity across all six major cell types (**Supplementary Fig. 26**). For each gene, we included expression measurements from all available cell types (up to six). Here, the G×CT component captures the extent to which genetic regulation varies across multiple cell types rather than between a specific pair. Consistent with the pairwise analysis, HEreg-base produced an inflated G estimate (~0.033), while adding G×CT without accounting for other variance components resulted in further inflation of G (~0.035) and a negative G×CT estimate (~−0.010). After adjusting for cell-type and individual-level effects, scHINT estimated substantially lower main genetic heritability (~0.013) and a positive G×CT heritability (~0.012). These results further demonstrate that a substantial component of genetic regulation varies across cell types and that explicitly accounting for cell-type and individual-level variation is necessary to appropriately separate shared from cell-type-dependent genetic effects.

8. **G×Cell-Type TWAS**

### 8.1 Model training

To evaluate whether modeling cell-type-dependent genetic effects improves gene-expression prediction, we trained elastic-net prediction models using cis-genotypes and cell-type-specific gene expression. Only genes with expression measurements in at least two cell types with non-zero expression variance were analyzed.

We compared three prediction strategies. First, a single-cell-type model was trained separately within each cell type using cis-genotypes to predict expression in that cell type. Second, a multi-cell-type main-effect model pooled expression measurements across all available cell types for a gene and used the shared cis-genotype effects only. Third, a multi-cell-type interaction model included both shared cis-genotype effects and cell-type-specific genotype interaction terms. Specifically, for each SNP, the interaction design matrix contained the main genotype dosage together with genotype terms multiplied by cell-type indicators, allowing the effect of the same cis-variant to vary across cell types.

All prediction models were fitted using elastic-net regression with glmnet, using an equal mixture of L1 and L2 penalties ($\alpha = 0.5$). The penalty parameter $\lambda$ was selected using internal five-fold cross-validation and the value minimizing cross-validation error (lambda.min). Non-zero coefficients from the fitted elastic-net models were retained as SNP weights.

For the interaction model, the predicted expression in a given cell type was decomposed into a shared genetic component, calculated from the main SNP weights, and a cell-type-specific genetic component, calculated from the corresponding SNP-by-cell-type interaction weights. The total interaction-model prediction was defined as the sum of these two components. Prediction scores and observed expression values were standardized within the held-out cell type before evaluation. Prediction performance was assessed by Pearson correlation between observed expression and predicted score. For the interaction model, we additionally fitted a joint regression containing both the shared and cell-type-specific prediction components to quantify their independent contributions.

Results were combined across the five held-out folds for each gene and cell type. After cross-validation, final prediction weights were re-estimated using all available donors for each gene using the same SNP filtering, model specification, elastic-net penalty, and internal five-fold selection of $\lambda$. These full-sample weights were subsequently used for downstream genetically predicted expression analyses.

### 8.2 Prediction performance

To evaluate how the estimated heritability components relate to genetic prediction performance, we stratified genes into bins according to either their main genetic (G) heritability or G×CT heritability. Within each heritability bin and cell type, we evaluated prediction performance using two complementary metrics: the proportion of gene–cell-type models with significant out-of-sample prediction and the cross-validated prediction ($R^2$). We compared the cell-type-specific model, the pooled model without interactions, and the G×CT model, with the latter further decomposed into its shared main genetic and cell-type-specific interaction components. This analysis allowed us to test whether genes with larger estimated G or G×CT heritability showed correspondingly greater out-of-sample predictability and, importantly, whether G×CT heritability was specifically associated with improved prediction from cell-type-dependent genetic effects (**Supplementary Fig. 33**).

Prediction performance increased strongly with both main genetic (G) and G×CT heritability. When genes were binned by G heritability, the proportion of significant prediction models and cross-validated ($R^2$) increased progressively with increasing heritability across all cell types. Models incorporating information across cell types generally outperformed cell-type-specific models, with the full G×CT model showing among the highest prediction rates and ($R^2$). For example, in excitatory neurons, the proportion of significant models for the full G×CT model increased from <20% in the lowest G-heritability bin to nearly 100% in the highest bin, while ($R^2$) increased from near zero to >0.20. This confirms that the estimated main genetic heritability is strongly predictive of the amount of cis-genetic signal that can be recovered by expression prediction models (**Supplementary Fig. 33 a and b**).

A distinct pattern emerged when stratifying by G×CT heritability. Both the proportion of significant models and prediction ($R^2$) increased with G×CT heritability, but this improvement was particularly pronounced for models incorporating cell-type-specific genetic effects. In excitatory neurons, for example, the full G×CT model increased from a relatively low prediction rate in the lowest G×CT-heritability bin to >80% in the highest bin, accompanied by a substantial increase in ($R^2$). In contrast, the shared main-effect component showed considerably weaker improvement across G×CT-heritability bins. Similar patterns were observed across the other cell types (**Supplementary Fig. 33 c and d**).

Together, these results provide a direct connection between the variance-component estimates and out-of-sample genetic prediction. High G heritability primarily indicates stronger shared cis-genetic predictability, whereas high G×CT heritability identifies genes for which allowing genetic effects to vary across cell types provides additional predictive information. Thus, the G×CT component estimated by scHINT represents not only statistical heterogeneity in genetic effects but also cell-type-dependent genetic signal that can be recovered in independent samples.

#### 8.3 Gene-trait associations

We next asked whether the improved cell-type-aware prediction models could identify disease-associated genes. As in the G×Cell-State TWAS analysis, we retained gene–cell-type models with significant cross-validated prediction and applied their trained weights to GWAS summary statistics from seven brain-related traits. We compared two sets of weights: conventional models trained within individual cell types, and G×Cell-Type models that combine the shared main genetic weights with the cell-type-specific interaction weights. The G×Cell-Type models identified more significant gene–trait associations (917) than the conventional within-cell-type models (679). Of the 1,126 associations identified by either approach, 470 (41.7%) were detected by both, indicating a substantial core of associations that is captured regardless of modeling strategy. However, many associations were specific to one approach: 447 (39.7%) were identified only by the G×Cell-Type models, and 209 (18.6%) only by the within-cell-type models..

**Reference**


1. Carver, S., Taraszka, K., Groha, S. & Gusev, A. Discovery of disease-associated cellular states using ResidPCA in single-cell RNA and ATAC sequencing data. *HGG Adv* **7**, 100538 (2026).
2. Kotliar, D. *et al.* Identifying gene expression programs of cell-type identity and cellular activity with single-cell RNA-Seq. *Elife* **8**(2019).
3. Consortium, G.T. The GTEx Consortium atlas of genetic regulatory effects across human tissues. *Science* **369**, 1318–1330 (2020).
4. Fujita, M. *et al.* Cell subtype-specific effects of genetic variation in the Alzheimer's disease brain. *Nat Genet* **56**, 605–614 (2024).
